# Whole-Genome Landscape of Breast Cancers from India shows Distinct Clinically Actionable Subtypes

**DOI:** 10.64898/2026.03.27.26349506

**Authors:** Divya Khanna, Arnab Ghosh, Priyanka Bhadwal, Ramakanth Chirravuri-Venkata, Leepakshi Dhingra, Subrata Das, Isha Choubey, Ankita Singh, Geeta Jadaun, Hriday Home Chowdhury, Vibha Mattoo, Pooja Dixit, Suryanshi Tiwari, Safeer Khan, Vinit Gupta, Tuneer R. Mallick, Surya Narayan Mishra, Tulasi Nagabandi, Venkata Krishna Vanamamalai, Malini Nemalikanti, Renu Kumari, Diksha Singhal, Laxmi Yadav, Vaishali Pandey, Shailya Verma, Rashmi Gudur, Senthil Kumar, Susanta Roychoudhury, Rohan Chaubal, Sonam Dhamija, Koushik Mondal, Lipi Thukral, Aruna Korlimarla, Rekha V Kumar, Ashutosh Mishra, Bhawna Sirohi, SVS Deo, Samir Bhattacharya, B S Srinath, Juhi Tayal, Anurag Mehta, IBCGA Consortium, Shilpak Chatterjee, Sanjeev Khosla, Divya Tej Sowpati, Kumardeep Chaudhary, Karthik Bharadwaj Tallapaka, Sherry Bhalla, Nidhan K. Biswas, Sudeep Gupta, Shantanu Chowdhury

## Abstract

Breast cancer remains the leading cause of cancer-related mortality among women globally, yet South Asian populations are critically underrepresented in genomic studies. Here, we present whole-genome and transcriptome analyses of over 500 treatment-naïve, clinically annotated breast tumors from India, representing the first large-scale integrative omics characterization of this population.

In addition to established drivers, we identify novel significantly mutated genes, including *ISM2*, *TERF2*, and *DHRSX*, under positive selection, along with previously unreported potentially pathogenic somatic variants. Notably, the distribution of key hotspot mutations appears shaped by immune escape pressures.

We uncover recurrent copy number alterations driving lipid metabolic reprogramming (e.g., *NCEH1* and *PLD1* amplifications) and complex structural events, including *IKZF3* promoter hijacking leading to *ERBB2* overexpression and *TTC28*-associated genomic instability—findings not previously described in breast cancer.

Mutational processes are dominated by DNA repair deficiency, APOBEC activity, and genome instability. We identify three genomic features significantly associated with poor post-surgical recurrence-free survival. Transcriptomic profiling reveals distinct intrinsic subtypes with divergent immune landscapes, including a high-risk HER2-androgenic cluster with the worst outcomes. Additionally, we find a novel transcriptomic signature that robustly captures a HER2-like transcriptional state within triple-negative breast cancer patients – with major implications for TNBC treatment.

Finally, clinically actionable germline and somatic alterations are revealed with high significance, highlighting opportunities for therapeutic repurposing. Together, the study establishes a comprehensive molecular atlas of breast cancer in an underrepresented population, providing critical insights into tumour biology and advancing precision oncology.

## Introduction

Breast cancer is the most common cancer in women globally, with more than 2.3 million new cases every year and approximately 670,000 deaths reported in 2022 (1). Beyond the classical differentiation based on estrogen receptor (ER), progesterone receptor (PR), and human epidermal growth factor receptor 2 (HER2/ERBB2) expression, breast tumours show extensive genetic heterogeneity. Currently, clinical management decisions are made based on pathology and the receptors mentioned above, classifying tumours as ER+, PR+, HER2+, or triple-negative breast cancers (TNBCs). Over the past decade, large-scale studies have significantly advanced our understanding of breast cancer heterogeneity at genomic and transcriptomic levels (2–7). Understanding this heterogeneity is key to devising personalized treatment strategies.

Seminal studies such as The Cancer Genome Atlas (TCGA) (2), the Molecular Taxonomy of Breast Cancer International Consortium (METABRIC) (4, 5), the multi-omics landscape analysis of Korean cohort (8) and the Fudan University Shanghai Cancer Centre (FUSCC) cohort (6, 7) have identified key driver mutations, defined intrinsic molecular subtypes, and provided insights into the immune landscape of breast tumours. However, these studies predominantly represent patients of European and East Asian descent, leaving substantial gaps in our knowledge regarding the global genetic landscape of breast cancer. Several studies have revealed ethnicity and region-specific differences in the mutation landscape and driver mutations involved in breast tumorigenesis across populations (9, 10), (11–13). Analysis of Nigerian and Tunisian breast cancer cohorts revealed distinct mutation spectra and aggressive tumour characteristics associated with African ancestry (9). Similarly, work on Mexican-Hispanic patients highlighted differences in tumour biology and gene expression profiles compared to non-Hispanic white women (10). The Taiwanese cohort uncovered subtype-specific differences in tumour microenvironments and therapeutic targets among East Asian populations (11). A study from Malaysia reported higher prevalence of HER2+ subtypes, higher prevalence of *TP53* mutations in the ER+ patients, and distinct immune signatures in Asian breast cancer (12). HER2+ subtypes were prevalent in another Asian cohort from Korea, which also displayed distinct enrichment of mutation signatures linked to homologous recombination repair deficiency (12, 13). These efforts collectively emphasize the influence of ethnicity, geography, and ancestry on breast cancer pathogenesis and underscore the need for diverse, regionally representative datasets.

South Asia, and particularly India, harbours immense genetic diversity due to its complex population history across centuries. India’s unique genetic structure calls for a dedicated investigation into the molecular underpinnings of breast cancer among its population. According to GLOBOCAN 2022 statistics, breast cancer is now the leading cancer in India, accounting for more than 192,000 new cases and over 98,000 deaths annually. Despite South Asia harbouring a sizable part of the world population and contributing towards many breast cancer patients, this region remains critically underrepresented in cancer genomics datasets. To address this significant gap, our manuscript presents the first-ever systematic genomic and transcriptomic profiling of breast tumours from an Indian cohort (acronym: IBCGA – Indian Breast Cancer Genome Atlas) (**Table 1**), representative of the broader South Asian population. Using integrative multi-omics approaches, we explore somatic alteration (mutational, copy number and structural alterations) patterns, gene expression landscapes, and immune cell infiltration profiles, and contextualize these findings against international and ethnicity-specific cohorts. Our breast cancer cohort represents a distinct distribution of IHC subtypes, specifically high prevalence of TNBC tumours (14), compared to well characterized Western cohorts (3, 4, 15). This was further confirmed by PAM50 intrinsic subtyping of the gene expression data, revealing a relatively high proportion of basal-like tumours. Our study, which represents the second largest whole-genome sequencing dataset for breast cancer, reveals novel insights into the molecular features of Indian breast cancer and provides a foundation for precision oncology approaches tailored to the Indian population.

**Table 1:**
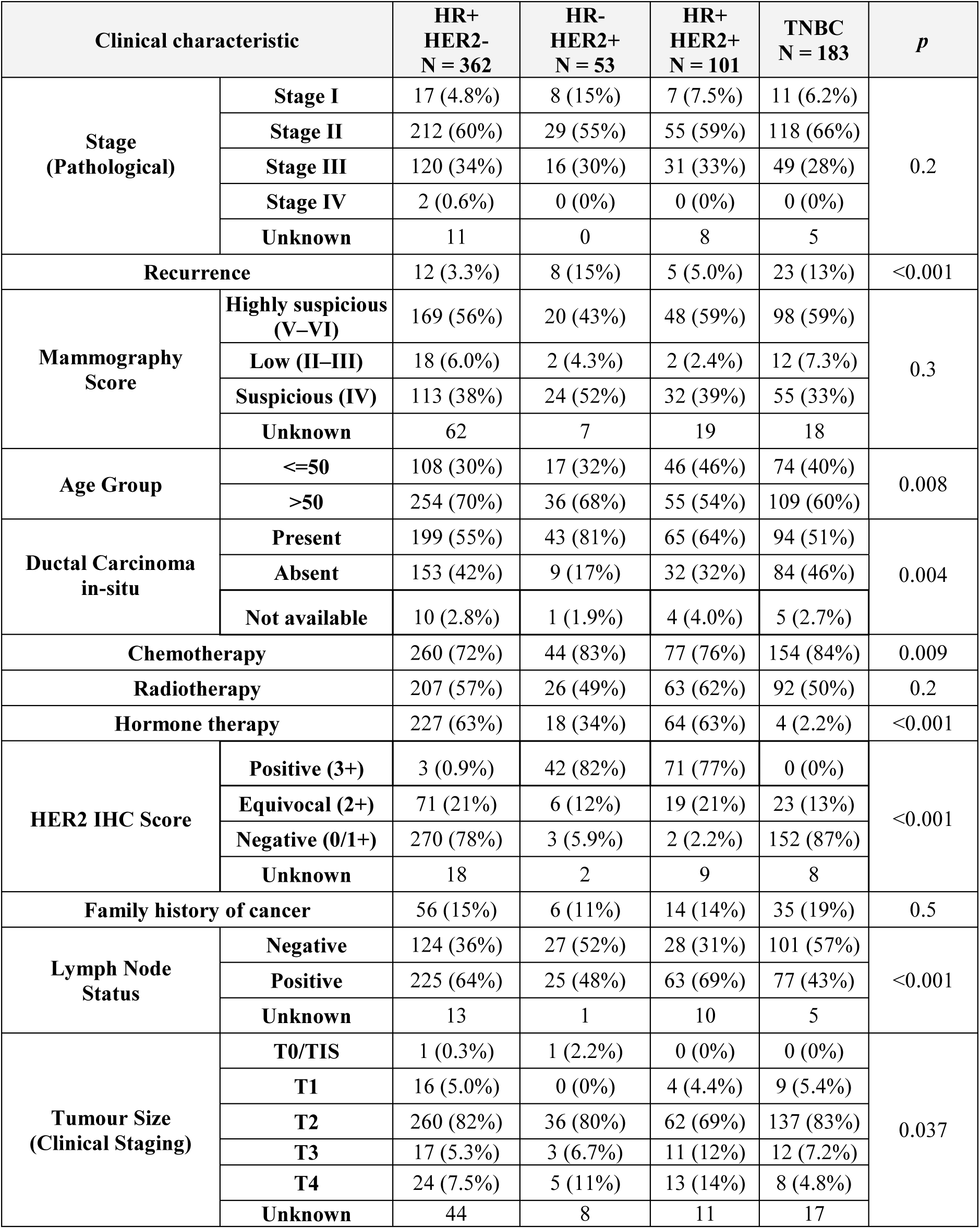
Clinicopathological characteristics of breast cancer patients from Indian cohort.

| Clinical characteristic |  | HR+<br>HER2-<br>N = 362 | HR-<br>HER2+<br>N = 53 | HR+<br>HER2+<br>N = 101 | TNBC<br>N = 183 | *p* |
| --- | --- | --- | --- | --- | --- | --- |
| Stage<br>(Pathological) | Stage I | 17 (4.8%) | 8 (15%) | 7 (7.5%) | 11 (6.2%) | 0.2 |
|  | Stage II | 212 (60%) | 29 (55%) | 55 (59%) | 118 (66%) |  |
|  | Stage III | 120 (34%) | 16 (30%) | 31 (33%) | 49 (28%) |  |
|  | Stage IV | 2 (0.6%) | 0 (0%) | 0 (0%) | 0 (0%) |  |
|  | Unknown | 11 | 0 | 8 | 5 |  |
| Recurrence |  | 12 (3.3%) | 8 (15%) | 5 (5.0%) | 23 (13%) | <0.001 |
| Mammography<br>Score | Highly suspicious<br>(V–VI) | 169 (56%) | 20 (43%) | 48 (59%) | 98 (59%) | 0.3 |
|  | Low (II–III) | 18 (6.0%) | 2 (4.3%) | 2 (2.4%) | 12 (7.3%) |  |
|  | Suspicious (IV) | 113 (38%) | 24 (52%) | 32 (39%) | 55 (33%) |  |
|  | Unknown | 62 | 7 | 19 | 18 |  |
| Age Group | ≤50 | 108 (30%) | 17 (32%) | 46 (46%) | 74 (40%) | 0.008 |
|  | >50 | 254 (70%) | 36 (68%) | 55 (54%) | 109 (60%) |  |
| Ductal Carcinoma<br>in-situ | Present | 199 (55%) | 43 (81%) | 65 (64%) | 94 (51%) | 0.004 |
|  | Absent | 153 (42%) | 9 (17%) | 32 (32%) | 84 (46%) |  |
|  | Not available | 10 (2.8%) | 1 (1.9%) | 4 (4.0%) | 5 (2.7%) |  |
| Chemotherapy |  | 260 (72%) | 44 (83%) | 77 (76%) | 154 (84%) | 0.009 |
| Radiotherapy |  | 207 (57%) | 26 (49%) | 63 (62%) | 92 (50%) | 0.2 |
| Hormone therapy |  | 227 (63%) | 18 (34%) | 64 (63%) | 4 (2.2%) | <0.001 |
| HER2 IHC Score | Positive (3+) | 3 (0.9%) | 42 (82%) | 71 (77%) | 0 (0%) | <0.001 |
|  | Equivocal (2+) | 71 (21%) | 6 (12%) | 19 (21%) | 23 (13%) |  |
|  | Negative (0/1+) | 270 (78%) | 3 (5.9%) | 2 (2.2%) | 152 (87%) |  |
|  | Unknown | 18 | 2 | 9 | 8 |  |
| Family history of cancer |  | 56 (15%) | 6 (11%) | 14 (14%) | 35 (19%) | 0.5 |
| Lymph Node<br>Status | Negative | 124 (36%) | 27 (52%) | 28 (31%) | 101 (57%) | <0.001 |
|  | Positive | 225 (64%) | 25 (48%) | 63 (69%) | 77 (43%) |  |
|  | Unknown | 13 | 1 | 10 | 5 |  |
| Tumour Size<br>(Clinical Staging) | T0/TIS | 1 (0.3%) | 1 (2.2%) | 0 (0%) | 0 (0%) | 0.037 |
|  | T1 | 16 (5.0%) | 0 (0%) | 4 (4.4%) | 9 (5.4%) |  |
|  | T2 | 260 (82%) | 36 (80%) | 62 (69%) | 137 (83%) |  |
|  | T3 | 17 (5.3%) | 3 (6.7%) | 11 (12%) | 12 (7.2%) |  |
|  | T4 | 24 (7.5%) | 5 (11%) | 13 (14%) | 8 (4.8%) |  |
|  | Unknown | 44 | 8 | 11 | 17 |  |

## Results

### Somatic mutational landscape of breast tumours

Whole genome sequencing of tumour and normal tissues from 501 breast cancer patients (**Figure S1-S3**) identified about 4,593,630 somatic mutations (12.53% are insertions or deletions) (**Figure S4**, **Table S1**), focal copy number alterations, and over 90 thousand structural alterations that varied significantly among IHC subtypes. Simultaneously, rare germline pathogenic alterations were also detected in DNA repair genes in a small fraction of patients, frequently among TNBCs (**Figure S5 and S6, Table S2**). The TNBC tumours harboured the highest somatic mutation rate (median = 2.13/Mb range: 0.08-35.88), followed by HR+HER2+ (2.07/Mb range: 0.20-35.71), HR-HER2+ (1.78/Mb range: 0.39-35.69), and the lowest detected for HR+HER2-tumours (1.42/Mb range: 0.05-28.97) (**Figure S4**). Both somatic copy number alteration and structural variation burdens in TNBCs were highest.

We have detected 16 significantly mutated (somatic) genes (SMGs) mutated in ≥ 5% of the tumours in any subtype, and ≥ 2% in the overall breast cancer cohort which includes previously reported mutational drivers like *TP53*, *PIK3CA*, *KMT2C*, *MAP3K1*, *GATA3* etc. along with driver mutations in *ARID1A*, *ERBB2* which are not frequently reported in breast cancer cohorts (**Figure S7-S12**, **Table S3**). Additionally, in the overall cohort, we found novel breast cancer SMGs, namely *ISM2*, *TERF2*, *DHRSX* etc. which were mutated in over 1% of the patients. 25 most significant SMGs are illustrated in **Figure 1A-H**, **Table S3**) and not reported in previous studies (driver mutations identified in our cohort with respect to five major global cohorts are illustrated in **Figure 1B**). Compared to global breast cancer cohorts (details in supplementary), we found a median of 86.13% nonsynonymous SNVs, and 27.27% InDels in these driver genes to be unique in our breast cancer cohort, including all unique somatic mutations detected in *ISM2*, *QRICH2*, *TERF2*, *RBP3*, *CBX3*, and *DHRSX* genes (**Figure 1C, Table S4,** and **Figure S13**). Multiple novel missense mutations in driver genes were predicted to be pathogenic by orthogonal prediction algorithms (**Table S5**), and a median of 16.67% of the novel mutations reside in conserved protein domains (**Figure 1D, E**). Our in-silico model predicted some of the novel *PIK3CA* mutations - N345S, E418K, and Q546E - to be associated with loss of stabilizing interactions of the protein (**Figure S14**, details in supplementary). We show that the prevalence of specific somatic hotspot mutations particularly in *PIK3CA* and *TP53* (**Table S6, Figure S15**) were negatively correlated with their neo-antigenic potential (**Figure S16**), providing evidence for evolutionary selection during tumorigenesis. Through estimation of cancer cell fraction (CCF), we found 42.9% somatic mutations are sub clonal suggesting substantial lineage diversification and intra-tumour heterogeneity. Alongside frequently mutated genes, multiple of the novel driver genes like *TERF2*, *CBX3*, and *DHRSX* in this breast cancer cohort showed very high dN/dS ratio (**Figure 1F**), indicating their evolutionary selection in the context of breast tumorigenesis.

**Figure 1:**
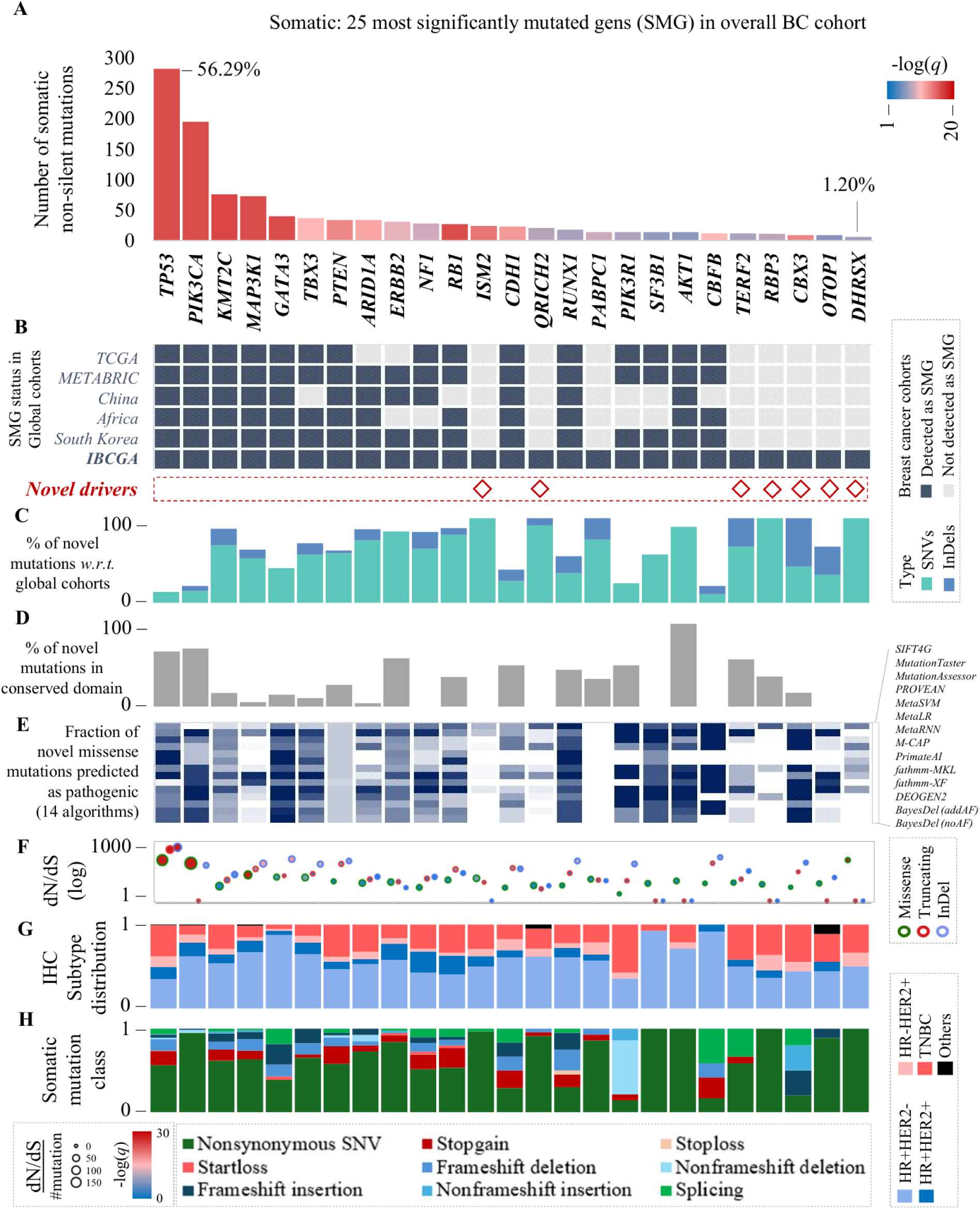
Genome alteration landscape of breast tumours. **(A)** Top 25 SMGs from WGS data of 501 breast tumors illustrated including 16 SMGs with over 5% frequency in any of the IHC subtypes (*TP53, PIK3CA, KMT2C, MAP3K1, GATA3, TBX3, NF1, ERBB2, RB1, AR1D1A,CDH1, PTEN, SF3B1, PIK3R1, CBFB,* and *AKT1).* **(B)** Comparison of SMGs in IBCGA cohort with previously identified SMGs in 5 major global breast cancer cohorts. 7 novel driver genes *ISM2, QRICH2*, *TERF2*, *RBP3*, *CBX3*, *OTOP1*, and *DHRSX* are significantly mutated in IBCGA cohort. **(C)** Percentage of novel somatic SNV and InDels in significantly mutated genes with respect to global breast cancer cohorts (35 datasets analysed; details in supplementary) are presented. **(E)** Pathogenicity of the novel missense mutations detected in the SMGs assessed through 14 orthogonal pathogenicity prediction algorithms listed on the right-hand side. **(F)** The dN/dS ratio (adjusted for GC content and background mutation rate) for the SMGs are illustrated showing strong selection for novel driver genes alongside previously characterized drivers. **(G)** The distribution of SMGs among the IHC subtypes HR+HER2- (n=260), HR+HER2+ (n=64), HR-HER2+ (n=45), and TNBC (n=128) **(H)** Nonsynonymous somatic mutations in the SMGs are further stratified into 9 variant functional classes as described in the bottom panel.

### Genome wide copy number alterations frequently affected genes involved in lipid metabolism

We identified recurrent arm-level amplification at chr6, and deletions at chr21 (details supplementary) in the breast tumours. Significant focal amplifications of *ERBB2*, *PI4KB*, *VMP1* [*MIR21* in same amplification peak], *PIK3CA*, *CCND1*, *MYC*, *ERLIN2*, *ECM1*, *MDM2* etc., and deletions of *CDKN2A*, *PTEN*, *RB1*, *ARID3A*, *BTNL3* etc., were detected in the overall cohort and these included previously known and novel genes (**Figure 2A, Table S7**). HER2+ tumours were found to be driven by *ERBB2* amplification (in 78.9% cases). Interestingly, we also detected *ERBB2* amplifications in 3.6% of the HER2- patients. TNBC tumours were significantly enriched with copy number amplification of *PIK3CA*, *PI4KB*, *TERT*, *TERC*, *SOX11*, *FGF14*, *GATA3*, and *EGFR*, and deletions of *RB1*, *ARID3A*, *BTNL3*, and *CREBBP* genes (**Table S7**). Copy number deletion of *PTEN*, *CDKN2A*, and amplification of *MYC* and *IL10* were present across all breast cancer subtypes.

**Figure 2:**
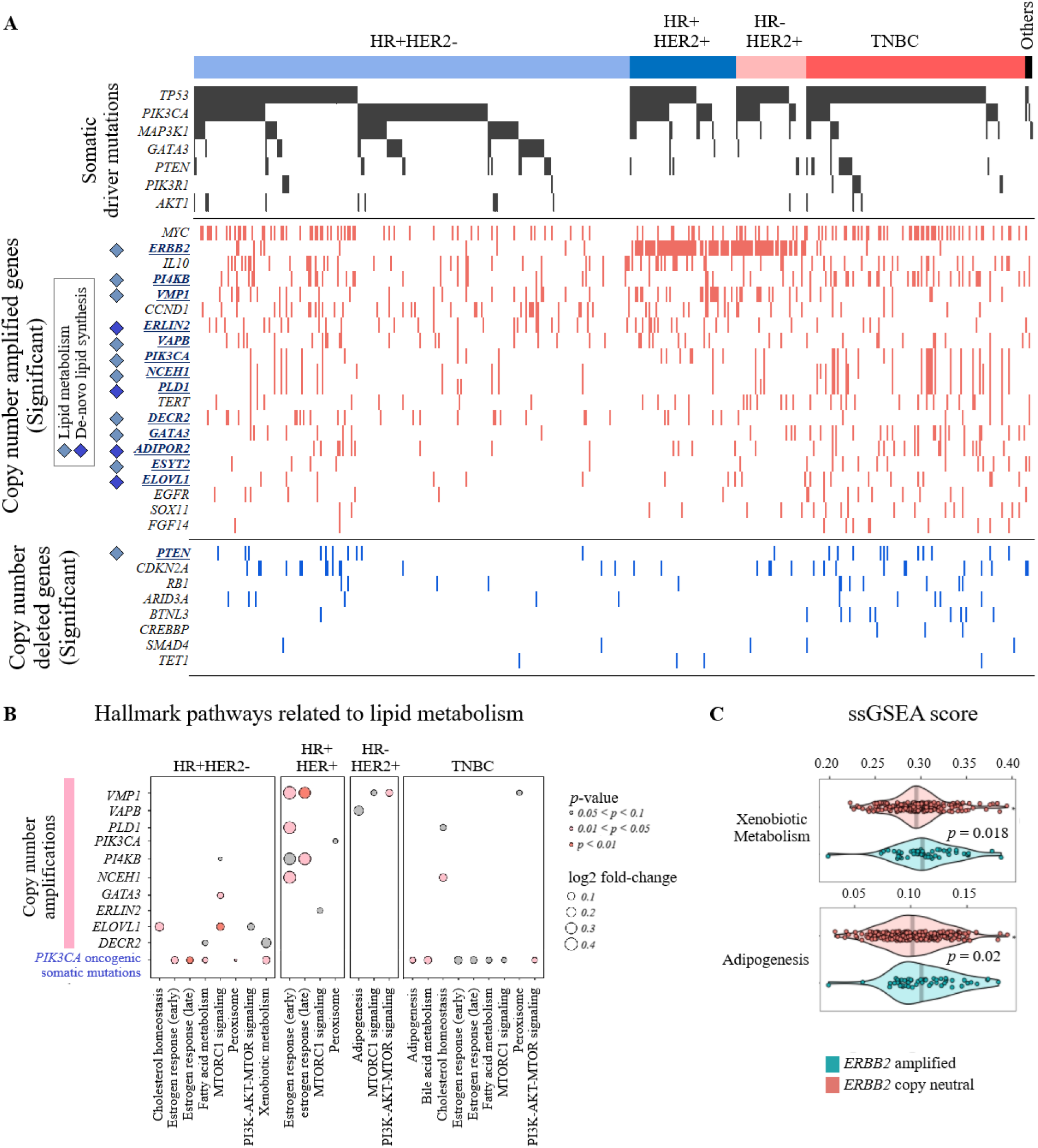
Somatic copy number aberrations in the breast cancer genomes. **(A)** Copy number amplification and deletion status (homozygous amplification or deep deletions) of genes belonging to significant (GISTIC, *q* < 0.1) focal copy number amplified or deleted regions (alteration length < 2Mb). Genes associated with lipid metabolism or de novo lipid synthesis are highlighted. **(B)** Differential enrichment of several lipid metabolism related hallmark pathways are shown to be associated with genomic alteration related genes with notable heterogeneity between IHC subtypes. **(C)** Copy number amplification of *ERBB2* was significantly associated with increased enrichment of gene expression of adipogenesis, and xenobiotic metabolism pathway in the breast cancer cohort.

Either PI3K-AKT activating somatic alteration or direct amplification of genes involved in lipid synthesis or lipolysis (16) is present among 70.26% of breast tumours, irrespective of their hormone receptor positivity status, and in TNBCs (**Figure S17**). About half of the hormone-positive breast tumors are either driven by activating mutations in *PIK3CA*, or by the amplification of *ERBB2* gene, both being directly involved in lipid metabolism including both lipid biogenesis and lipolysis. Additionally, a copy number amplification of *PIK3CA,* along with several key members of the PI3K-AKT pathway including *PI4KB* (17), *GATA3* (18), and copy number deletions of PI3K-AKT suppressor *PTEN* (19) was detected predominantly in TNBCs (**Figure 2A**). Among the significantly amplified genes in this breast cancer cohort, we found a frequent copy number amplification of *NCEH1*, *PLD1*, *ADIPOR2*, *ESYT2*, specifically in about 9-19% TNBC patients (**Figure 2A**, **Figure S17**, **Table S7**), which are known to be directly involved in lipid metabolism (20–23). Additionally, we observed significant expression enrichment of the glycolysis pathway in the context of *PIK3CA* activating mutations in TNBCs (**Figure S18**). Apart from this, several other lipid metabolism-related genes, *ERLIN2*, *VMP1*, *DECR2*, and *VAPB* (22–26) were frequently amplified among hormone-positive tumours (in about 9-11%) (**Figure 2A**, **Figure S17**, **Table S7**), and *ERBB2* and ELOVL*1* (27) were found to be frequently amplified specifically among HER2+ tumours. We found that the mRNA expression levels of multiple genes with copy number amplification were involved in the lipid metabolism (**Figure S17**). The alterations in lipid metabolism-related genes not only showed varying frequency among IHC subtypes but also showed differential expression of metabolic pathways detected through ssGSEA analysis (**Figure 2B**). We observed that the *ERBB2* copy number amplification was significantly associated with increased expressions of adipogenesis, and xenobiotic metabolism pathways (**Figure 2C**). Oncogenic *PIK3CA* mutations were found to be significantly associated with an increase of estrogen response, xenobiotic and fatty acid metabolism, and peroxisome activities in HR+HER2- tumours, while the same mutations, when in TNBC tumors were associated significantly with bile acid metabolism, adipogenesis, and PI3K-AKT-mTOR signalling in TNBC tumours (**Figure 2B**). *NCEH1* amplification, which frequently co-occurred with *PIK3CA* amplification, showed significant association with cholesterol homeostasis in TNBCs (**Figure 2B**). Contrastingly, amplification of *VMP1*, *PI4KB*, *NCEH1*, and *PLD1* was significantly associated with estrogen response, and PI3K-AKT-mTOR signalling in HER2+ tumours (**Figure 2B**). Across all four subtypes, these lipid-related copy-number alterations are not isolated events but co-selected components of a functional “lipid toolkit” that operates in an intimate partnership with *PIK3CA* oncogenic signalling or *ERBB2* amplification. Together, they establish a multi-compartment system spanning de novo synthesis, fatty-acid specialisation, intracellular trafficking, storage, and detoxification.

### Structural variation rewires ERBB2 regulation and genome stability

From WGS, we identified 90,555 high-confidence somatic structural variations (SVs) including local intrachromosomal breaks (65.2%), and intra and inter-chromosomal reshuffling events (20.2 and 14.5%) (illustrated in **Figure 3A, Figure S19-20**). The HR+HER2+ tumours harboured highest SV burden (median = 214), followed by TNBCs (157), HR-HER2+ (131), and lowest was detected in HR+HER2- tumours, and a fraction of SVs were also detected to be pathogenic through ACMG classification (**Figure 3B**). The type of SVs varied significantly among breast cancer subtypes (**Figure S21-23**) as evident from frequent duplications in TNBCs, and frequent rearrangements (inversion, and translocations), and deletions among HR+HER2+ tumours. High density of SVs, affecting ≥ 5% tumours, were recurrently detected in - (1) chr17:35-40Mb (944 SVs; encompassing *ERBB2*, *IKZF3*, *CDK12*), (2) chr11:65-70Mb (561 SVs; encompassing *CCND1*), (3) chr10:120-125Mb (408 SVs; encompassing *FGFR2*), (4) chr10:85-90Mb (237 SVs; encompassing *PTEN*), (4) chr9:5-10Mb (208 SVs, encompassing *KDM4C*), (5) chr6:150-155Mb (191 SVs, encompassing *ESR1*), and (5) chr6:155-160Mb (173 SVs, encompassing *ARID1B*) - regions containing key genes involved in chromatin organization, cell cycle, and growth (**Figure 3A**). The *ERBB2* flanking regions (76 Kb upstream and 143 Kb downstream) were enriched with SV breakpoints (specifically inversions and duplications) (**Figure 3B, E**), these genomic regions are also known to be enriched with breast tissue specific super enhancers (**Figure 3E**). We identified a novel structural rearrangement associated with *ERBB2* overexpression through hijacking strong promoter of nearby gene *IKZF3* (**Figure 3E**). From the CNV and SV analysis, we show two plausible mechanisms behind *ERBB2* overexpression - (1) focal copy number amplification of *ERBB2*, and (2) over enrichment of enhancers through frequent duplications and local-rearrangements near *ERBB2* upstream and downstream regions. Several recurrent genomic rearrangements in HER2+ tumours mostly converged towards *ERBB2* overexpression mediated tumorigenesis.

**Figure 3:**
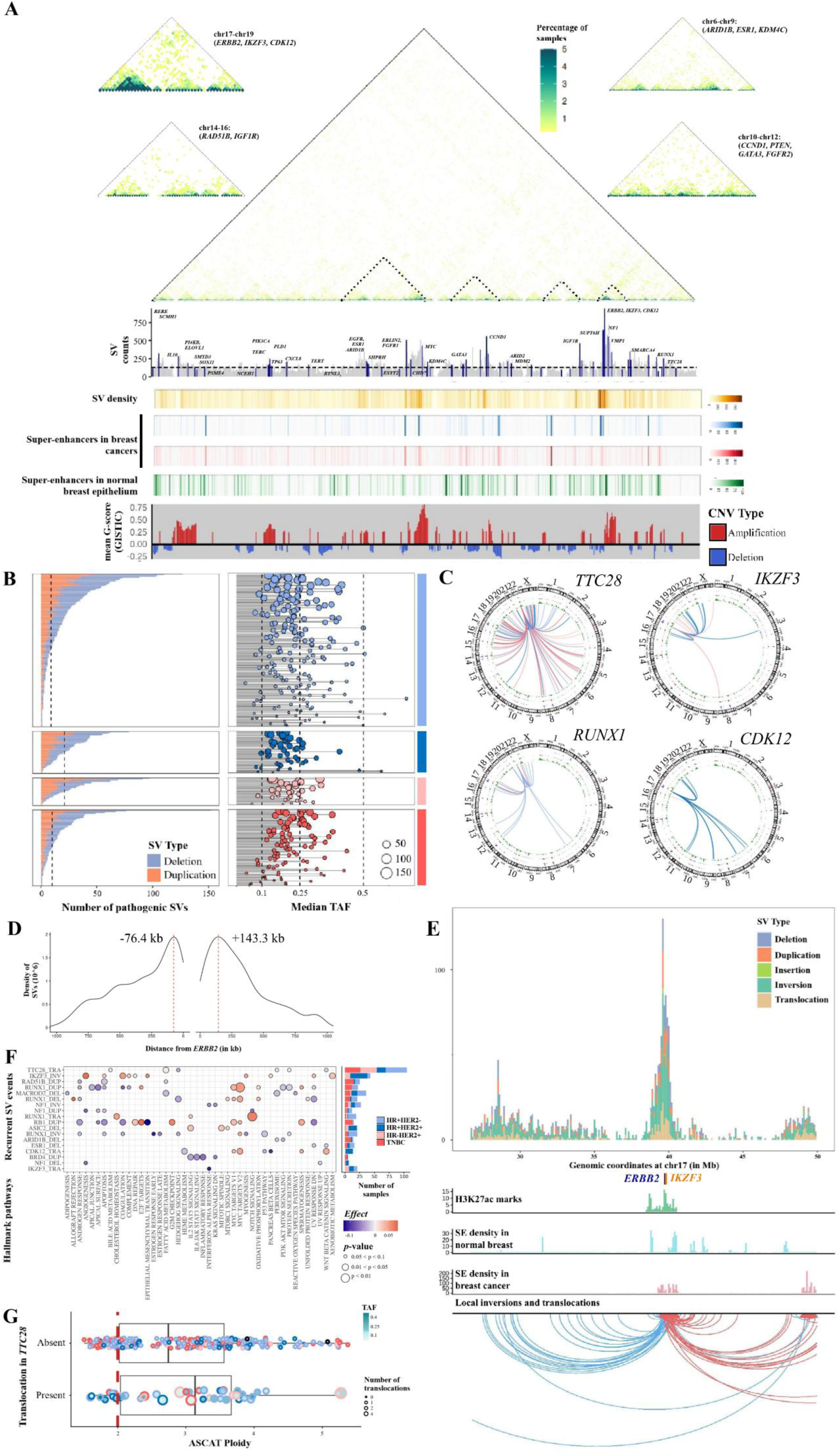
Integrated landscape of structural variations, copy-number alteration and transcriptomic deregulation in breast cancers. **(A)** From top to bottom: Genome-wide interaction map of inter-chromosomal and intra-chromosomal structural variants across 5 Mb windows. Frequent events between chr17-19 (top-left) encompassing *ERBB2*, *IKZF3*, and *CDK12*, chr6-9 (top right) encompassing *ARID1B*, *ESR1* and *KDM4C*, chr. 14-16 (bottom left) encompassing *RAD51B* and *IGF1R* and chr10-12 (bottom right) encompassing *CCND1, FGFR2, GATA3, ARID2* are highlighted. Middle, number of SVs across 5Mb windows indicating recurrent genes affected by SVs. and super-enhancers (SEs) in normal breast epithelium and breast cancer per window. Bottom, recurrent CNVs identified by GISTIC. **(B)** Left, distribution of pathogenic deletions and duplications (through ACMG classification) in each patient across IHC subtypes. Dotted line indicates median number of pathogenic SVs per IHC subtype. Right, median tumour allele fractions (TAFs) of pathogenic SVs in each patient. **(C)** Circos plots showing recurrent translocation events originating from *TTC28* (chr22), *IKZF3* (chr17), *CDK12* (chr17), and *RUNX1* (chr21). Color indicating IHC subtype of samples affected. **(D)** Distribution of structural variant breakpoints (BPs) within 1 Mb of *ERBB2*. **(E)** Top, Distribution of SV types affecting chr17:25-30 Mb encompassing *ERBB2* (dark blue) and *IKZF3* (orange). Middle, distribution of H3K27ac marks around *ERBB2* and *IKZF3* (light green). Number of super-enhancers (SEs) in normal breast epithelium (light blue) and breast cancer (pink). Bottom, Local 3’-to-5’ (blue lines) and 5’-to-3’ (red lines) inversion and translocation events affecting the locus. **(F)** Association between recurrent SV events and hallmark pathway activity inferred from transcriptomic data. Color indicates magnitude and direction of association, and size indicates level of significance (two-tailed Wilcoxon rank-sum test) (left). Number of SV events across IHC subtypes (right). **(G)** Distribution of ASCAT-inferred ploidies between patients with TTC28-L1 translocations and wild-type TTC28 patients. Colour indicates median TAF of translocation events, size indicates number of translocation events involving *TTC28* in each patient, and border color indicates IHC subtype

We identified the *TTC28* gene to be most frequently affected by aberrant translocations (12.1% patients, mostly abundant in HER2+) (**Figure 3C**). The majority of *TTC28* translocations (101 out of 105 events) breakpoints lie within 10Kb of an active L1 retrotransposon element which might lead to its inactivation. Although retrotransposon-mediated inactivation of *TTC28* has previously been reported in ovarian cancer (28), we first show that *TTC28* aberration is associated with increase in ploidy (i.e. whole genome duplication) in breast cancer (**Figure 3F**) as well as decreased expression of *TOP2A*, *PTTG1*, *KNTC1*, *NCAPD3*, *TPX2*, *GEN1*, *ECT2*, *GPSM2*, and *CEP350*, involved in chromatic assembly and mitosis - suggesting increased genome instability.

Although SV events were predominantly present in HER2+ tumours, we found recurrent translocations of key transcription factor *RUNX1* in HR+ tumours affecting 3.6% patients (**Figure 3C**), which has previously been shown in multiple other cancer types. Additionally, we found recurrent (≥ 5% patients) SVs driven aberration of genes like *ARID1B*, *ESR1*, *RB1*, *MACROD2* and *BRD4* in overall breast cancer cohort, and *ASIC2* in HER2+, and *RAD51B* in TNBCs. These SVs were shown to affect hallmark pathway activities such as angiogenesis, apical surface, apoptosis, and NOTCH signalling, inflammatory response, E2F targets (**Figure 3G**). Overall, we identified a wide spectrum of structural rearrangements in the breast cancer genome.

### Defective DNA repair and APOBEC signature modulates key transcriptional programs

We identified 13 key somatic single base substitutions (**Figure 4A, Table S8**), 6 insertion-deletion (**Table S9**), 17 copy numbers (**Figure 4B, Table S10**), and 2 predominant structural variation (**Figure 4E**) signatures in the breast cancer cohort. Single base substitutions are primarily contributed by double stranded break repair (DSBR) failure (SBS3), and APOBEC hyperactivity (SBS2 and 13), apart from tumour aging (primarily SBS5). We found the somatic mutation rate was correlated with the abundance of both DSBR-failure and APOBEC signatures separately. The homologous recombination defect (HRD) related signature ID6 was found to be significantly correlated with SBS3 (**Figure S24**). The DSBR failure related mutagenesis found to be the single dominant program in TNBC tumours (apart from tumour aging). The HRD-associated copy number signatures CX14 and CX1, and structural variation signature SV3 related to HRD were correlated with SBS3 and found to be significantly higher among TNBCs compared to others (**Figure S25**). The TNBCs with higher contribution of SV3 signature also showed significantly poor recurrence free survival as compared to the tumours with lower contribution (**Figure S25**). TNBC tumours with lower SBS3 related mutagenesis showed significantly high expressions of multiple pathways related to – (i) lipid metabolism, i.e., cholesterol homeostasis, bile acid and xenobiotic metabolism, and (ii) estrogen and androgen response (**Figure 4D, Figure S26**). Based on mutational signatures, hormone positive (HR+ and/or HER2+) tumours could be classified into three broad classes – (1) DSBR-failure high – APOBEC low (17.39% of tumours), (2) APOBEC high – DSBR failure low (21.47%), and (3) others. In DSBR failure high – APOBEC low hormone positive tumours, we found significantly higher frequency of – (i) rare germline alteration in *BRCA2* and *BRCA1*, (ii) somatic mutations in *TP53*, (ii) copy number deletions of *PTEN*, and *CDKN2A*, and (iii) copy number amplification of notably, *MYC*, *CCND2*, *BIRC5* (**Figure 4C**). We further show significant enrichment of E2F targets, and G2M checkpoint hallmark pathways in these tumours from the gene expression data. On the other hand, APOBEC high – DSBR failure low hormone positive tumours showed significantly higher frequency of – (i) somatic mutations in *PIK3CA*, *TBX3*, and *ARID1A*, (ii) copy number deletion of *SMAD4*, and (iii) copy number amplification of *ERBB2*.

**Figure 4:**
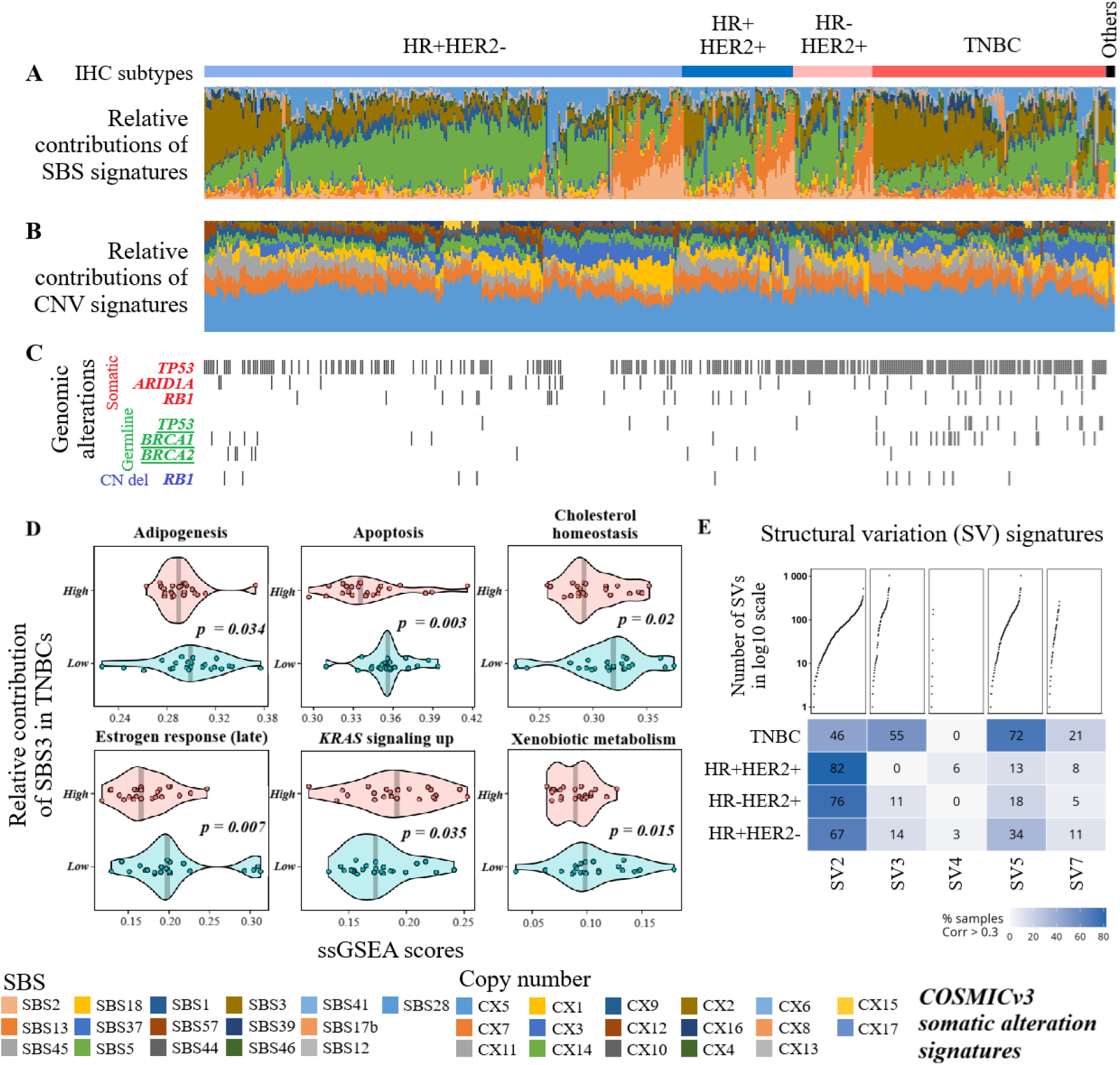
Somatic alteration signatures in breast cancer genome. **(A)** Somatic mutational signatures in Indian breast tumours. The somatic mutational signature SBS3 [median proportion of contribution = 14.78%], SBS5 [29.68%], SBS1 [3.73%]), SBS2 [3.47%], and SBS3 [4.30%] (associated with APOBEC driven mutagenesis) was found to be predominant in these tumours. The relative contribution of SBS2 and SBS13 related to APOBEC hyperactivity was most predominant among HER2+tumoursspecifically with HR positivity [median proportion of contribution was 17.86% ranging between 1.78-87.28% in these tumours]. The relative contribution of SBS3 (HR-defect) [median contribution of 28.33% in TNBCs], and SBS39 (unknown etiology) [median contribution of 4.17% in TNBCs] was found to be significantly (*p* < 5.043e-14, t-test) higher among TNBC tumours than other hormone positive subtypes [median contribution of SBS3, and SBS39 in hormone positive tumours respectively 11.74% and 1.43%]. We observed that HR-defect (relative contribution of SBS3) was significantly (*p* = 1.494e-10, *t*-test) associated with *TP53* somatic mutational status in the breast cancer cohort. **(B)** Contrasting differences of copy number alteration signatures in Indian breast tumours. 6 predominant copy number signatures CX5 [median proportion of contribution = 36.66%], CX3 [7.04%], CX1 [7.00%], CX14 [7.06%], and CX11 [9.64%], were detected in this cohort, which are associated with interhomolog recombination (IHR) with replication stress, IHR, chromosome mis-segregation, and replication stress, and additionally signature CX2 for which the etiology is not known. The contributions of CX14 and CX1 (both linked to chromosome mis-segregation) were found to be significantly greater among hormone positive tumours than TNBC. While CX3 (linked to IHR) was consistently found to be predominant in almost all TNBCs, we observed about one-third of the HR+/HER2-tumours have significantly greater contribution of CX3 than the rest of the tumours of the same group. Overall HER2+tumours (with or without ER/PR positivity) showed lower abundance of CX3 compared to both HR+/HER2- group and TNBC. **(C)** Genomic alterations associated with SBS3 prevalence along with TP53 alterations are shown with respect to mutational and copy number signature profiles across breast cancer subtypes. **(D)** Higher relative contribution of SBS3 (linked to DSBR-failure and HRD) was shown to be associated with expression level alterations of several pathways involved in estrogen response, oncogenic signalling and lipid metabolism. **(E)** The relative contributions of 5 predominantly identified somatic structural variation (SV) signatures are represented for each IHC subtypes of breast tumours.

Overall, somatic signature profiling identified two dominant mutagenesis programmes in breast tumours – (1) defective DNA repair and (2) APOBEC activity – that showed varying prevalence among the subtypes and were found to influence cell cycle, immune infiltration and energy metabolism.

### Three genomic features associated with poor recurrence-free survival

Presence of *PIK3CA* hotspots (E545K, E542K, and N345K), or *TP53* hotspots (R273H, R175H), [HR = 3.6 95% CI: 1.16-11.5, *p* = 0.027] (**Figure 5A**) and *SF3B1* somatic mutations [HR = 4.6 95% CI: 1.19-18.0, *p* = 0.027] (**Figure 5B**) significantly reduced recurrence free survival probability of HR+HER2- patients when jointly analysed with clinical covariates (age, and stage) through multi-variable Cox-PH model. These *PIK3CA* or *TP53* hotspot mutations also significantly decreased recurrence-free survival probability in HER2+ patients [HR = 3.93, 95% CI: 1.11-13.89, *p* = 0.0289] (**Figure 5C**) after adjusting clinical covariates. Contrastingly, in TNBC patients, recurrence-free survival probability was significantly (*p* = 0.017) decreased in presence of *PIK3CA* mutations (including hotspot and non-hotspots) or *GATA3* copy number amplification with a HR of 3.1 (95% CI: 1.23-7.8) (**Figure 5D**) when jointly analysed with clinical covariates through Cox-PH. Further by integrative analysis with the transcriptomics data, we show the three-recurrence associated genomic alterations were involved in activation of namely, EMT, E2F targets, cell cycle, myogenesis, glycolysis, angiogenesis, adipogenesis, androgen response, and fatty acid metabolism with strikingly different effect sizes among the breast cancer subtypes (**Figure 5E**). Additionally, 36 (7%) tumours in the cohort that developed distant metastasis (in liver, bone, lung, brain, and breast) during the follow-up, revealed frequent somatic mutations in *PIK3R1*, *CDH1*, and *ERBB2*, and copy number amplification in *MYC* and *ERBB2* (**Table S11**). Specific hotspot mutations in *PIK3CA* and *TP53* were repeatedly detected among these tumours.

**Figure 5:**
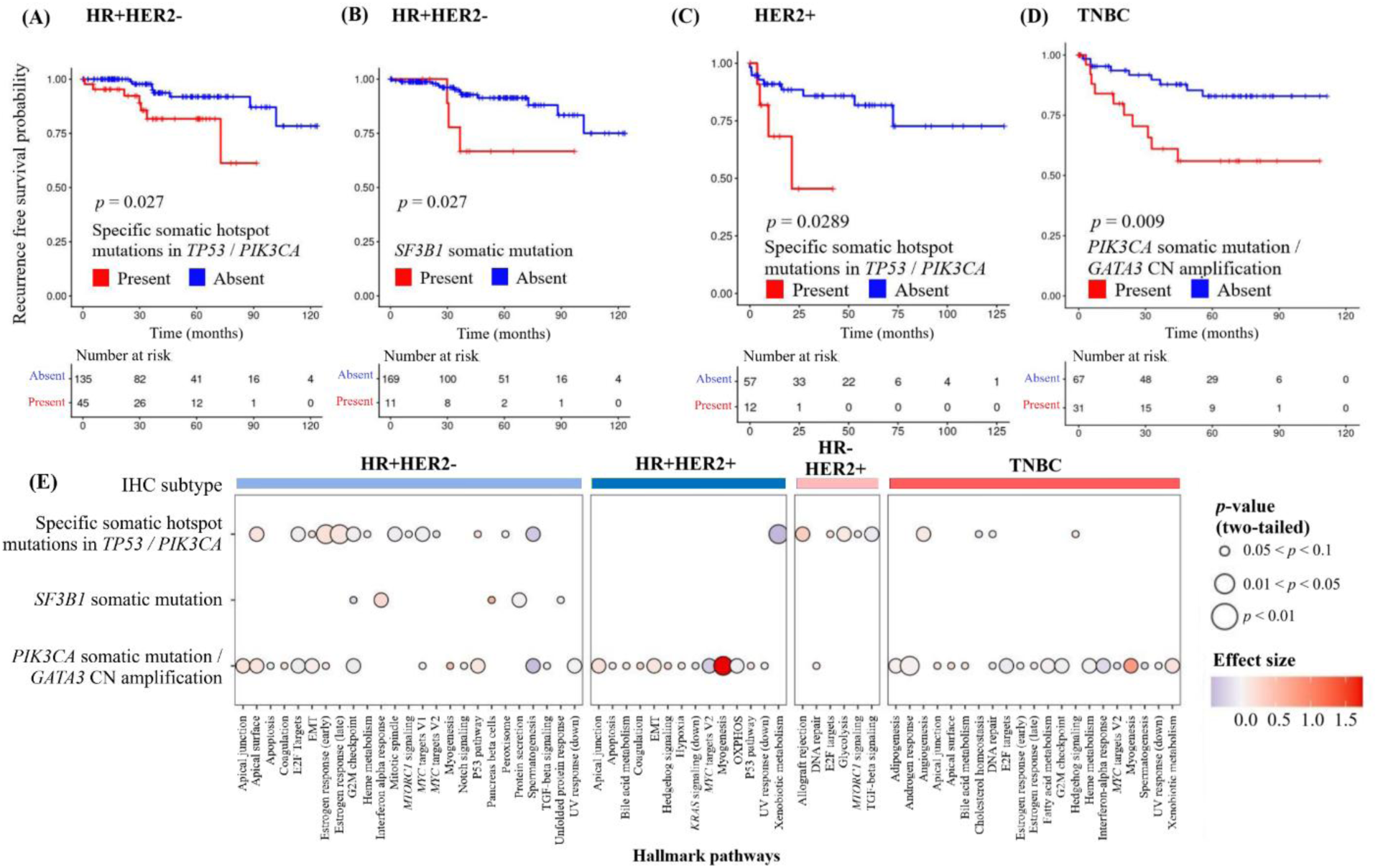
Genomic alteration features associated with poor recurrence-free survival. Integration of patient follow-up data (“disease recurrence”: recurrence and/or death) with somatic alterations in Indian female breast cancer patients followed by Cox-proportional hazard multivariate modelling with clinical covariates age, and tumour stage identifi–d - **(A)** specific hotspot mutations in *TP53* or *PIK3CA* in HR+HER2- patients, **(B)** *SF3B1* somatic mutations in HR+HER2- patients, **(C)** specific hotspot mutations in TP53 or PIK3CA in HER2+ patients, and **(D)** *PIK3CA* somatic mutations or *GATA3* copy number amplification – significantly increased hazard-ratio (HR) for recurrence-free survival, **(E)** The three genomic features associated with significant reduction of recurrence-free survival are shown to regulate various pathways related to tumourigenesis with significant heterogeneity between IHC subtypes of breast tumours.

### Transcriptional heterogeneity of breast tumours

We performed unsupervised clustering using whole-transcriptome data (*N* = 438) (**Table S12**) and identified four major expression clusters including (C1) Basal-proliferative (BaPro) [*n* = 170], enriched for TNBCs (61.2%); (C2) Luminal-proliferative (LumPro) [*n* = 132], predominantly composed of Luminal B tumours (61.4%)] with a smaller fraction of TNBCs (8.3%); (C3) Luminal-differentiated (LumDiff) [*n* = 92], enriched for Luminal A (38.0%) and Normal-like subtypes (28.3%); and (C4) HER2-androgenic (HER2-AR) [*n* = 44], enriched for tumours classified as HER2 subtype by PAM50 prediction **(Table S13)** and comprising a subset of TNBCs (43.2%); illustrated in **Figure 6A (Table S17, Figure S27,S28,S29)**. Collectively, these clusters captured distinct proliferative, luminal differentiation, and HER2-androgen signalling programs across the cohort as summarized in **Table S20**. The tumours of BaPro cluster harboured recurrent *TP53* somatic mutations; the LumPro cluster was significantly enriched for somatic mutations in *ERBB2* and *PIK3CA*; and LumDiff tumours were primarily characterized by enrichment of *PIK3CA* mutations, suggesting PI3K pathway dependence in a relatively lower-proliferative context (**Figure 6A**, **Table S18** and **S19**).

**Figure 6:**
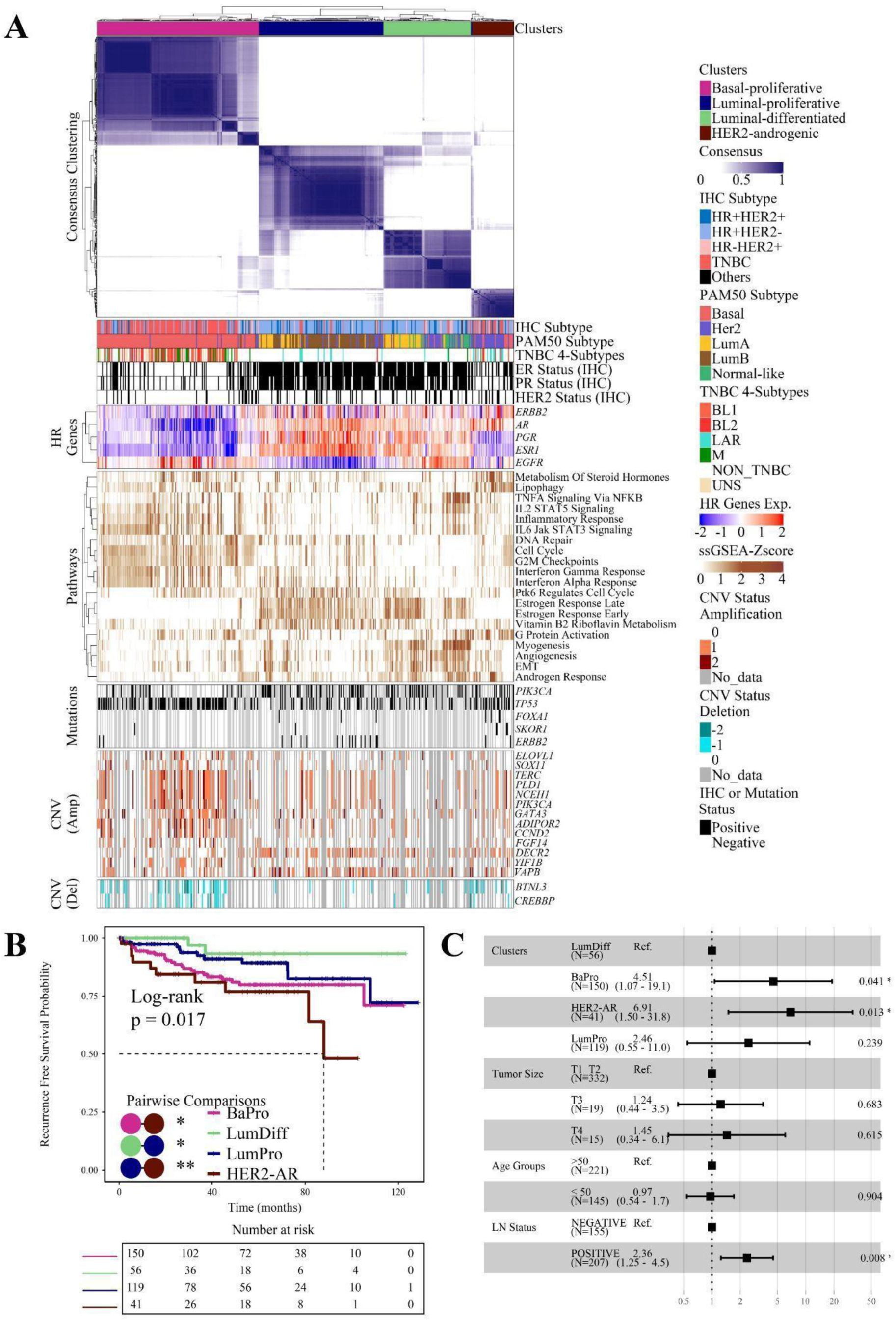
Molecular characterization and prognostic significance of consensus clustering subtypes. **(A)** Consensus clustering heatmap of 438 breast tumours showing four robust transcriptional clusters (Basal-proliferative, Luminal-differentiated, Luminal Proliferative, HER2 Androgenic) with distinct patterns of hormone receptor gene expression, mutation profiles, copy-number alterations, and clinical immunohistochemical (IHC) subtypes. Cluster-specific enrichment of molecular features highlights biologically distinct tumour subgroups. **(B)** Kaplan–Meier analysis of recurrence-free survival (RFS) across the four clusters demonstrates significant outcome differences (log-rank *p* = 0.017), with the HER2–androgenic cluster exhibiting the poorest prognosis. **(C)** Multivariable Cox proportional hazards regression confirms the independent prognostic value of cluster assignment after adjustment for key clinical covariates, including age, lymph node status, and tumour size

These transcriptomic tumour clusters also varied significantly with respect to the tumour immune microenvironment **Figure 6B**. The BaPro cluster exhibits significant upregulation of IL6-JAK-STAT3, IL2-STAT5, and Interferon Gamma (IFNG) pathways, and enrichment of onco-immunological signatures like CSF1 or macrophages, as well as higher leukocyte infiltration driven by an interferon response. Interestingly, a small subset of TNBCs belonging to the BaPro cluster (14%) were classified as Basal-like Immune Suppressed (BLIS) subtype by Burstein classification, marked by high proliferation, low score of interferon signatures, and fewer immune infiltrate, unlike classical basal types **(Table S14)**. The LumPro and LumDiff clusters, characterized by luminal proliferative and differentiated tumours, showed significantly (*q* < 0.05) higher enrichment of TGF-beta signalling, stromal content, upregulation of ECM glycoproteins, and collagen content, compared to others. Interestingly, the LumDiff cluster showed the highest and most consistent enrichment of TGF-beta signalling, angiogenesis, and epithelial to mesenchymal transition (EMT) signatures. The recurrence-free survival of the HER2-AR cluster comprising HER2+ and a fraction of TNBCs was found to be significantly poorer (*p* = 0.017, log-rank) compared to the other patient clusters (**Figure 6B-C**) **(Table S15, S16)**. In these HER2-AR tumours, we identified upregulation of pathways related to steroid hormone metabolism, lipophagy, and G-protein– coupled receptor signalling, indicating a metabolically specialized and hormone-regulated tumour state as well as increased infiltration of regulatory T-reg cells.

Overall, our comprehensive gene expression profiling of breast tumours revealed four transcriptionally distinct clusters with striking differences in oncogenic programs, energy metabolism, cytokine signalling, and tumour and immune microenviroment - suggesting a requirement for deeper phenotyping and classification for breast cancer **(Table S12)**. Given the substantial representation of TNBCs across these clusters, we also performed TNBC-specific transcriptomic clustering, which revealed a metabolically distinct fatty acid–enriched subgroup associated with the poorest survival (*p* = 0.039, log-rank) (**Figure S30A-C**, **Table S23).**

### Transcriptionally HER2-Enriched TNBCs in the Indian Patients

Emergence of transcriptome data on breast tumours revealed mRNA level expression of *ERBB2* gene in a fraction of HER2-negative tumours, exposing limitations in classical IHC-based testing (17) and misclassification among TNBCs (29), specifically in understudied populations gene expression analysis comparing tumours with an IHC sc. Therefore, to systematically classify breast tumours with HER2-like transcriptional activity, we derived a 144-gene signature from differential ore ≥3 (high protein level expression of HER2) with those with an IHC score of 0 (no protein level expression) (**Table S21**). Ten of these HER2-specific differentially expressed genes, including *GRB7*, *CDK12*, *MIEN1*, etc., including *ERBB2,* mapped to the chr17q12 locus and were uniformly overexpressed (**Table S21**). We used this 144-gene signature and re-classified all HR-tumours that resolved into two distinct transcriptional s–ates - (1) a HER2-Enriched (HER2-E) cluster marked by ERBB2 overexpression and downstream pathway activation, and (2) a basal-like cluster (**Table S22**). Strikingly, about one-fourth of classical TNBCs (27 of 137 with available gene expression data) were found to be transcriptionally very similar to tumours with high protein level HER2 expression, as illustrated in **Figure 7A**. We have defined these TNBCs as HER2-enriched TNBCs. Notably, HER2-enriched mRNA tumours showed strong representation of the luminal androgen receptor (LAR) subtype, with 18 of 43 HER2-E cases (41.9%) classified as LAR. These tumours were also frequently identified as HER2-enriched by PAM50 classification.

**Figure 7:**
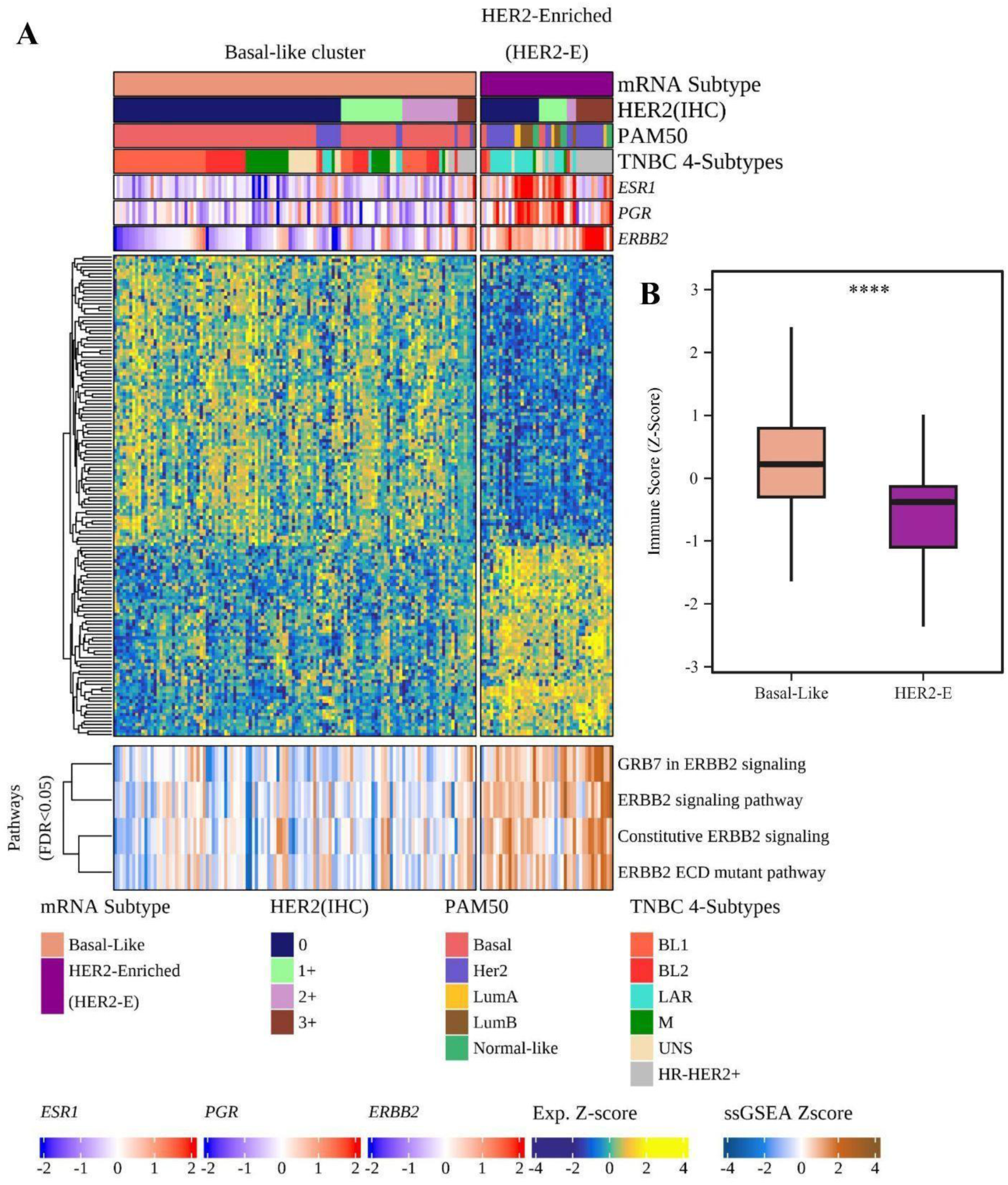
HER2 IHC Stratification Reveals Distinct ERBB2 Expression and Subtype Patterns. **(A)** Transcriptomic heatmap comparing Basal-like and HER2-Enriched (HER2-E) tumours based on HER2/IHC-associated gene expression profiles. Annotation tracks display HER2 IHC score, PAM50 subtype, TNBC 4-subtype classification, and expression levels of key markers (*ESR1*, *PGR*, *ERBB2*). Pathway activity scores derived from ssGSEA demonstrate increased activation of ERBB2-related signalling programs in the HER2-E group, including *GRB7*/*ERBB2* signalling and constitutive *ERBB2* pathway activation. **(B)** Boxplot illustrating immune score differences between subtypes. HER2-E TNBC tumours exhibit significantly lower immune scores compared with Basal-like TNBCs, indicating a relatively immune-cold and less infiltrated tumour microenvironment.

Overall, mRNA subtype and PAM50 classification were strongly associated (Fisher’s exact test, *p* < 0.01). This agreed with the findings from a recent study on HER2-low Chinese breast cancer patients (30). These HER2-E TNBCs exhibited significantly lower immune scores than basal-like TNBCs (**Figure 7B**), suggesting a colder, less immune-infiltrated tumour microenvironment. These results put further emphasis on developing population-specific molecular markers for more accurate classification of breast tumours to improve patient outcomes with precision therapeutics.

### PD-L1 upregulation via interferon-responsive JAK-STAT signaling in basal-proliferative tumours

Substantial variation of immune cell infiltration, states and expression of immunoregulatory factors are observed among the breast tumours which also differed significantly between molecular and IHC subtypes (**Figure S31-S34**). The BaPro tumours which also included a fraction of classical hormone positive tumours were found to be the most immune enriched subtype along with significant enrichment of HALLMARK immune pathways including JAK-STAT signalling, and IFNγ-response. Interestingly Th (T-helper) cell differentiation associated - (i) transcription factors *STAT1* and *STAT4*, (ii) key kinases *JAK2*, *JAK3*, and (iii) *NOTCH1* - were found to be overexpressed in basal-proliferative tumours. Consistent with the above findings, the upregulation of Th1 cytokines (IFNγ, and IL12) in basal-proliferative tumours indicates that this cluster represents a Th1-enriched immune phenotype (**Figure S35**). The basal proliferative tumours also showed upregulation of immune-checkpoint molecules including *CD274* (encodes PD-L1), *CTLA4*, *LAG3* and *TIGIT*. The KEGG path view maps indicate that the upregulation of PD-L1 in these tumours is driven by interferon-responsive JAK-STAT (*STAT1*) and HIF1a (*HIF1a*) signalling pathways.

### Landscape of clinically actionable molecular alterations in breast cancer cohort

Several rare germlines pathogenic or likely pathogenic variants in DNA repair genes, previously associated with hereditary cancer predisposition - *BRCA1*, *TP53*, *BRCA2* and *CHEK2*, *RAD50*, *RAD51D*, *MUTYH*, *ATM* - detected in ≥ 2% and 1-2% Indian breast cancer patients. Rare germline variants in *BRCA1* and *TP53* were significantly enriched among TNBC tumours, and rare germline *BRCA2* variants were predominantly detected in HER2+ tumours. Only 2.5% of patients with a reported family history of cancer harboured pathogenic rare germline alterations. Patients with rare germline mutations in specifically *BRCA1/2* exhibit higher abundance of DNA repair failure related mutagenesis (as evident from higher contribution of SBS3 and SV3 signatures). PARP inhibitor driven synthetic lethal strategies may be exploited in these sets of breast cancer patients with DNA repair alterations.

FDA approved drugs for breast ductal carcinoma in-situ are available against 42 somatic mutations detected in 194 (38.72%) patients - comprising 35 targetable *PIK3CA* mutations, 5 *PTEN* mutations, 1 *AKT1* and 1 *ESR1* mutation (**Figure S36**). We identified three *ERBB2* mutations in 5 patients, six somatic mutations in *TP53*, *ATM*, *EGFR*, *IDH1*, *KRAS*, *SMARCA4* in 10 patients three mutations in *CDKN2A*, *FGFR1*, and *PPP2R1A* in 3 patients for which targetable molecules are available and some of which are in clinical trials. Additionally, our transcriptomic cluster revealed HER2 negative TNBCs who can be benefitted from HER2-targetable drugs along with HER2+ patients. Despite high immune infiltration basal-like tumours showing overexpression of immune checkpoint molecules, ICI-therapeutics may be effective in these patients.

## Discussion

We report the first whole-genome study establishing a comprehensive molecular landscape of breast cancer in an Indian cohort. The data generated provides a critical resource. Through multi-omics integration over 500 Indian breast tumours, we capture the mutational and transcriptional heterogeneity among Indian breast cancer patients, which will aid precision therapeutics. Earlier studies in Caucasian populations have associated >10% of familial breast cancer with germline BRCA1/2 mutations (31). In contrast, less than 3% of the Indian breast cancer patients with a family history harboured pathogenic rare germline variants in DNA repair genes. While we identified the somatic mutations in classical driver genes like *TP53* and *PIK3CA*, our analysis revealed over 80% unique mutations in 16 driver genes in this cohort. Alongside, we found the prevalence of somatic mutations in *ARID1A* and *ERBB2*, which are not typically reported as primary drivers in other breast cancer cohorts. Some of these somatic alterations in silo or in combination - such as oncogenic mutations in *PIK3CA* or *TP53*, *SF3B1* amplification, *PIK3CA* mutation or *GATA3* amplification, showed a higher abundance of SV3 signature which significantly reduced recurrence-free survival of the patients in our cohort. Interestingly, these recurrence-associated genomic alterations are found to be involved in differential regulation of pathways related to cell cycle, oncogenic signalling, and metabolism among the IHC subtypes. Crucially, we identified widespread deregulation of energy metabolism pathways - either through activating somatic mutations in PI3K pathway genes or focal copy number amplifications of genes such as *NCEH1*, *PLD1*, etc. involved in lipid metabolism or de-novo fatty acid synthesis, even in traditionally metabolically cold TNBC tumours. Our comprehensive study also highlights the presence of actionable somatic alterations in a large fraction of these breast cancer patients.

Deeper transcriptomic profiling of the cancer cohort establishes a refined molecular taxonomy of Indian breast cancers, revealing four transcriptionally distinct subtypes, that we define as Basal-proliferative (BaPro), Luminal-proliferative (LumPro), Luminal-differentiated (LumDiff), and HER2-androgenic (HER2-AR), with unique biological features and clinical trajectories. Despite large agreements between the transcriptomic subtype and classical IHC subtype, transcriptomic profiling was able to highlight IHC-related misclassifications. The BaPro subtype emerges as a genomically unstable that is marked by high prevalence of DSBR failure-related mutagenesis and greater somatic alteration burden. Still, it depicts an immune-active disease state, characterized by concurrent proliferative and interferon signalling. Despite immune infiltration, the BaPro tumours showed features of immune tolerance marked by CD4+ T cell infiltration and higher expression of several immune checkpoint molecules. In contrast, the LumPro subtype demonstrates estrogen-driven proliferation with *ERBB2* and *PIK3CA* alterations, while LumDiff tumors represent a transcriptionally mature, low-proliferative state with favourable outcomes. Most strikingly, the HER2-AR subtype defines a hormonally regulated, high-risk group with the poorest survival, enriched for androgen signalling. Importantly, our analysis demonstrates that HER2-associated transcriptional programs extend beyond tumors classified as HER2-positive by immunohistochemistry. The TNBCs within the HER2-AR cluster, which we define as HER2-ive, were not only found to be transcriptionally distinct from classical TNBCs, but they also lacked interferon signalling and displayed overall lower immune infiltration, and relatively lower abundance of M1 macrophages. The molecular profiles identified in our large cohort of Indian breast cancer patients are consistent with prior studies on smaller Indian cohorts (32). Specifically, the shared key driving alterations and gross intrinsic subtypes across these studies confirm that Indian patients exhibit a distinct molecular landscape compared to Western populations. In addition to the previous knowledge, our multi-omics data generated from over 500 breast cancer patients significantly improved patient sub-stratification and identified previously unreported driving alterations involved in metabolic reprogramming, progression, and poor outcomes in Indian populations. Collectively, these insights establish a new benchmark for breast cancer stratification in South Asia, transforming our understanding of the disease’s heterogeneity in respect to precision therapeutic strategies that address the unique molecular vulnerabilities of the Indian cohort.

## Methods

### Patient recruitment and sample collection

The study was designed as an ambispective analysis, including both prospectively and retrospectively collected samples. All the patients recruited were treatment-naïve women, aged 18 years or older, and diagnosed with breast carcinoma. The retrospective samples were selected where they had a minimum of 3 years of follow-up data and were treatment naive. These were procured from biorepositories maintained by hospitals post all ethical clearance (IRB reference: RGCIRC/IRB-BHR/41/2022; KVV/IEC/01/2025). For prospective sample collection, patients were enrolled prospectively from multiple participating hospitals across India. After a rigorous series of clinical, pathological, and sequencing quality assessments, 671 patients were retained for downstream genomic and transcriptomic analyses. The study was approved by the Institutional Human Ethics Committee of the CSIR–Institute of Genomics and Integrative Biology (Ethics Approval ID: CSIR-IGIB/IHEC/2020-21/02). Written informed consent was obtained from all enrolled patients.

For each patient, detailed clinico-pathological information—including AJCC stage, ER/PR/HER2 status by IHC, nodal status, menopausal status, Ki-67 index and family history—was systematically recorded (Supplementary table 1). If HER2 expression could not be conclusively determined by IHC, fluorescent in situ hybridisation was performed for confirmation. Tumours were categorized into HR+, HER2−, HR+, HER2+, HR-, HER2+, and triple-negative breast cancer (TNBC). A minimum follow-up of three years was available for all patients, documenting treatment, recurrence, and survival outcomes. Tumour biopsies were either snap-frozen or preserved in RNAlater at 2–8 °C and later stored at −80 °C. For retrospective samples, the adjacent normal tissue was collected, and for prospective samples, peripheral blood was collected as matched normal. All protocols adhered to established ethical guidelines for human subject research.

### DNA and RNA isolation and sequencing

Whole genome sequencing was performed on tumour samples obtained as per routine clinical care surgery or biopsy either as fresh or frozen tissue. High-quality DNA and RNA were extracted together from the fresh frozen tumour and adjacent normal tissues using the Qiagen AllPrep DNA/RNA/miRNA Universal Kit. The quality of the isolated nucleic acids was assessed by spectrophotometry (A260/A280), fluorimetry, and RNA Integrity Number (RIN value), and only those that exceeded specific thresholds were eligible for further library processing. The Illumina TruSeq DNA PCR free (Illumina) was used for library preparation. Sequencing was performed on the Illumina Novaseq 6000 (Illumina) platform with an average depth coverage of 40X for tumour and adjacent normal tissue with S4 reagent kit (Illumina).

For whole transcriptome sequencing (WTS), RNA was extracted using the Qiagen AllPrep DNA/RNA/miRNA Universal Kit. The libraries were prepared after rRNA removal by using NEBNext rRNA Depletion Kit v2, followed by directional library preparation by employing NEBNext Ultra II Directional RNA Library Prep Kit. Libraries were validated by Agilent TapeStation (DNA 1000 Screen Tape) and Qubit dsDNA HS Kit. The libraries were sequenced by paired-end sequencing (2×150 bp) protocol on the NovaSeq6000 platform with the S1 reagent kit.

### Processing and analysis of WGS data to identify germline and somatic mutations and copy number alterations

Raw WGS data at about ∼ 40X for paired tumour and normal samples were demultiplexed with Illumina adapter information through bcl2fastq (version: v2.19.0.316) followed by quality assessment through FastQC (version: 0.11.7). Only high-quality paired-end reads were mapped to the human reference genome (GRCh38) followed by post alignment processing (sorting, duplicate marking, base quality score recalibration, and merging) through BWA-MEM, Picard and GATK based accelerated pipeline. Post-alignment QC was done using MapInsights (33) and utilized the CalculateContamination module of GATK (4.0.5.1) to verify tumour-normal parity using the genomAD resources. Germline joint genotyping was performed from normal samples using gVCF calling with accelerated HaplotypeCaller, followed by joint genotyping with GLnexus (version: v1.4.1), and functional annotation with annovar and ClinVar (version: Release:2025-06-01). The pathogenic germline variants were identified as per ACMG guidelines for breast cancer. Somatic mutations were identified by jointly analyzing tumour and paired normal samples through Mutect2 (GATK version: 4.0.5.1) using ‘--normal-lod -10 and --force-active’ parameters and genomAD as germline resources to increase sensitivity. Somatic call sets were further filtered to obtain only high-quality mutations utilizing information such as depth, strand orientation, variant allele frequency, low complexity regions (homopolymer and/or sequence similarity) (32). Further variant annotations (including pathogenicity prediction by multiple algorithms and InterPro domain annotations) for high-quality somatic mutations were performed through annovar, and population-specific germline resources (India-specific in-house control sets including > 1800 individuals). Any polymorphic sites (AF > 0.01 in 1000Genome populations, population-specific resources) were removed from the somatic mutation call set. We have utilized MutSig2CV for the identification of significantly mutated genes (SMGs), and somatic mutation rate (as mutations per Mb of genome and exome separately), in the overall cohort and within clinical sub-groups. Variants in the SMGs were manually verified through visual inspection using IGV (version: 2.17.4) by two independent individuals, followed by removal of potentially artefactual variants. We utilized a Random Forest framework (34) that integrated pathogenicity scores from 11 existing algorithms to classify missense mutations identified in significantly mutated genes. The tumour and normal WGS data were segmented using dense population-specific germline marker sets through AscatNGS, followed by identification of (a) both arm-level and focal copy number amplification and deletions, and (b) significantly amplified and deleted genes through GISTIC (version:v2.0.22). Some of the copy number events were further validated through sample-specific qRT-PCR data generation. We have utilized Maftools (version 2.22.0) for statistical assessment of mutual co-occurrence and exclusivity of somatic alteration events in the overall cohort and the clinical subgroups.

### Detection of somatic mutational and copy number signatures

We have utilized accelerated SignatureAnalyzer with COSMICv3-reference to identify single base substitution (SBS) signatures from SNVs through the extraction of substitutions based on tri-nucleotide content, followed by cosine similarity calculation. Similarly, somatic insertions and deletions were used for the detection of indel (ID) signatures. The results from signature fitting through SignatureAnalyzer were verified through an orthogonal method using SigProfileExtractor . From the genome-wide somatic copy number profiles (generated through AscatNGS: 4.5.0), we have detected copy-number signatures as described by Steele et al.2022.

### Identification of somatic structural variations

We identified structural variants (SVs) from WGS data of 501tumoursand their matched normal samples using a multi-caller strategy. The aligned BAM files were analysed with three tools—GRIDSS (Genomic Rearrangement Identification Software Suite), Manta, and SvABA—and the results were compiled in VCF format. Variants detected by at least two of the callers were considered high-confidence, resulting in consensus SV calls for 439 samples. GRIDSS (v2.13.2) (35) was run with its default parameters for somatic SV detection. GRIPSS incorporates evidence from discordant read pairs, split reads, and soft-clipping to generate high-confidence SV calls for the identification of translocations and complex rearrangements. Manta v1.6.0 (36) was used in somatic mode for tumour-normal pairs, detecting a wide range of SV types. It integrates paired-end and split-read signals and is optimized for rapid SV detection. SvABA v1.1.0 (37) was used to call somatic SVs via a local assembly-based approach. It uses a custom implementation of SGA (String Graph Assembler). Manta and SvABA were used in a chromosome-wise chunked mode to increase computational efficiency. However, since this strategy may lead to incomplete detection of inter-chromosomal translocations (TRA), the break ends of translocation calls reported by GRIPSS, which was run on the entire genome without chunking. Following SV calling, individual VCF files were merged using Jasmine v1.1.5 (38) to generate a consensus call set. Jasmine clusters SVs across samples and tools based on breakpoint proximity, strand orientation, and SV type. A maximum positional distance threshold of 500 bp was used for merging, and the SVs supported by at least two or three callers were retained to ensure high-confidence consensus calls. The translocations reported by GRIPSS were manually validated for their presence in the merged VCF files. The consensus call sets were annotated with AnnotSV v3.4.4. (39) Then, a BED format file was created focusing on protein-coding regions and guided by a reference GTF annotation file, and coordinates were expanded 2kb upstream using the bedtools slope function (40). Downstream analysis was performed with in-house custom scripts to process and merge SV calls from three SV callers (GRIDDS, MANTA, and SvABA) and to obtain supported variants from Jasmine.

### RNA-Seq Data Processing and Transcript Quantification

Adapter trimming and removal of low-quality bases from the raw RNAseq data were carried out using Trimmomatic v0.39 (41). To assess and eliminate ribosomal RNA (rRNA) contamination, reads were first aligned to the rRNA reference sequence KY962518.1 using bbsplit.sh (from BBMap v38.18) to estimate rRNA content. Subsequently, bbduk.sh was employed to remove rRNA reads using the same reference. A second pass of bbsplit.sh validated rRNA depletion efficiency. All preprocessing steps were executed on a high-performance computing cluster with default parameters unless otherwise specified. Clean reads were aligned to the human reference genome (GRCh38.p14, accession: GCA_000001405.29, release: December 2013), using the splice-aware aligner STAR v2.7.10b (42). Sample-level QC was also analysed after alignment by monitoring sequencing metrics such as uniquely mapped read percentages (≥75%) and total read counts (>10 million) from STAR BAM files, summarized using MultiQC v1.27 and reported in Supplementary Table 1. Gene-level quantification was performed using the featureCounts function from the Subread v2.0.6 package (43). For transcript-level quantification, RSEM v1.2.28 (44) was used with a reference prepared using rsem-prepare-reference, based on the GRCh38.p14 genome and matched GENCODE GTF annotation. Expression values were reported in both TPM (Transcripts Per Million) and FPKM (Fragments Per Kilobase Million) units via the rsem-calculate-expression function.

### Normalization and batch effect correction

The initial dataset consisted of 466 tumour samples (141 TNBC, 324 non-TNBC, 1 other) across 28 experimental batches, with expression data for 62,754 genes. Batches with fewer than three samples were excluded. Genes expressed in fewer than 10% of samples were removed, resulting in a final matrix of 455 samples (140 TNBC, 314 non-TNBC, 1 other) and 25,556 genes across 23 batches. Library size estimates were computed per sample, and tumour purity was estimated using tidyestimate v1.1.1. To quantify technical and biological sources of variation, VariancePartition v1.32.5 was used. To remove technical artifacts while preserving biological signals, we applied the RUVIII-PRPS normalization approach (45). Negative control genes were selected based on low contribution from biological signals (e.g., PAM50 subtypes) and high variance explained by technical variables (e.g., batch, purity, library size). Pseudo Replicates of Pseudo Samples (PRPS) were constructed to estimate unwanted variation. Normalization was conducted using the RUVIII model, and its performance was evaluated via principal component analysis and variance decomposition pre- and post-correction.

### Transcriptome-Based Molecular Subtyping

Intrinsic subtypes were assigned using the PAM50 gene signature with the AIMS algorithm (46), implemented via the AIMS R package v1.32.0. This method classifies tumours into five subtypes: Luminal A, Luminal B, HER2-enriched, Basal-like, and Normal-like, based on pairwise gene expression comparisons. For samples classified as triple-negative breast cancers (TNBCs) by immunohistochemistry, further subtyping was performed using two independent frameworks. The TNBC subtype classification (47) stratified tumours into six transcriptionally defined subtypes: BL1, BL2, IM, M, MSL, and LAR. To focus on tumour-intrinsic biology, IM and MSL subtypes were excluded as per Lehmann et al., 2016 (48), yielding four consolidated subtypes: BL1, BL2, M, and LAR. In parallel, the Burstein classification (49) was applied, categorizing TNBCs into LAR, Mesenchymal (MES), Basal-like Immune Activated (BLIA), and Basal-like Immune Suppressed (BLIS) subtypes based on correlation with published centroids.

### Unsupervised Consensus Clustering and Downstream Analyses

To stratify tumours based on transcriptional profiles, consensus clustering was performed on 454 samples using the top 5,000 most variable genes, implemented in the ConsensusClusterPlus R package (v1.66.0). Clustering used hierarchical agglomeration with Euclidean distance and Ward.D2 linkage, 1,000 resamplings, and 80% item resampling. The optimal number of clusters (k) was selected using the delta area plot, CDF curve, and average silhouette width. Samples with negative silhouette scores were excluded, and the clustering procedure was repeated until all remaining samples exhibited positive silhouette widths (50). We assessed pathway activity by applying single-sample Gene Set Enrichment Analysis (ssGSEA) to MSigDB Hallmark and C2–Reactome gene sets using the GSVA R package (v1.50.5) (51–53). Cluster-specific genomic alterations, including somatic mutations and copy number variations (CNVs), were analyzed using Chi-square (χ²) tests implemented in the stats R package (v4.3.3). To explore transcriptional heterogeneity within TNBCs, consensus clustering was independently performed on the clinically defined cohort of 140 TNBC samples using the top 3500 variable genes. Clustering was conducted using k-means agglomeration with Euclidean distance and Ward.D linkage, applying 1,000 resampling iterations with 80% item resampling. Downstream pathway analysis was done using the Hallmark gene set (54), and survival analyses were then carried out as described above.

### HER2 IHC Stratification and Transcriptomic Characterization

Hormone Negative (HR-) tumours were stratified into HER2 immunohistochemistry (IHC) categories (0, 1+, 2+, and 3+) according to clinical pathology scoring. The *ERBB2* gene was quantified from RUVIII-normalized RNA-seq expression data and compared across HER2 IHC groups (45) . To identify transcriptional programs associated with HER2-high biology and to detect HER2-like tumours within hormone receptor–negative patients, differential gene expression analyses were performed across HER2 IHC strata. Genes differentially expressed between HER2 IHC3+ and HER2 IHC0tumourswere first identified. To capture heterogeneity within lower HER2-expressing groups, the HER2 IHC0, IHC1+, and IHC2+ categories were further subdivided based on median *ERBB2* expression. Differential expression analyses were then performed between ERBB2-high and ERBB2-lowtumourswithin each respective IHC category. Genes consistently upregulated in IHC3+tumoursand shared with at least two additional lower-IHC ERBB2-high comparisons were defined as a HER2-associated overlap signature. The HER2-associated overlap gene set was subsequently used as input for consensus clustering restricted to hormone receptor–negative tumours, enabling identification of transcriptionally defined HER2-like subgroups independent of clinical HER2 IHC status. To characterize signaling differences between Basal-like and HER2-like tumour groups, ssGSEA pathway activity scores were computed as described above using MSigDB Hallmark and C2–Reactome gene sets through the GSVA framework (v1.50.5) (51–53).

### Immune signatures and pathway scoring

Immune related transcriptional programs were assessed using well established gene sets from MSigDB, including Hallmark, KEGG, GO, and the immune collection described by Thorsson et al. Additional pathways relevant to tumour immunobiology, such as macrophage differentiation, IFNG-signalling, lymphocyte chemotaxis, antigen processing, cytotoxic effector functions, Th1 and Th2 differentiation, adhesion signalling, and extracellular matrix programs curated by Naba et al., were also included. Cell type specific transcription factors and lineage markers from the Human Protein Atlas were incorporated to support the interpretation of immune phenotypes. For all curated pathways, enrichment scores were calculated for each sample using ssGSEA and compared across consensus clusters, TNBC subgroups, and clinical categories.

### Immune cell deconvolution and microenvironment estimation

Immune and stromal cell populations were inferred using EPIC, xCell, and CIBERSORT with the LM22 reference matrix in absolute mode and 1,000 permutations. To minimize batch related effects, FPKM matrices from each sequencing batch were analyzed separately. Only samples meeting the CIBERSORT significance threshold were retained. Tumour purity adjusted infiltration was estimated using ESTIMATE, and ssGSEA was applied to RUVIII normalized matrices to derive immune, stromal, and extracellular matrix scores. Results were then combined to enable comparisons across clinical subsets, PAM50 subtypes, and TNBC clusters.

### Estimation of tumour infiltrating immune cells and cross cohort comparison

Immune cell fractions were estimated for all transcriptomes using EPIC, xCell, and CIBERSORT. For cross cohort benchmarking, STAR FPKM expression matrices and clinical data for TCGA BRCA were downloaded from UCSC Xena and processed using the same normalization, deconvolution, purity adjustment, and pathway scoring strategy. Only samples with positive immune scores from ESTIMATE were included in abundance comparisons.

## Data Availability

Processed and raw sequencing data from the Indian Breast Cancer Genome Atlas (IBCGA) will be accessible through the IBCGA data portal (https://csir-ibcga.igib.res.in/). The portal provides public access to processed datasets with gene-wise query capabilities and interactive visualisation tools. Raw sequence data are available under controlled access via the same portal. Researchers are required to submit a data access request outlining the proposed research to the Data Management Group. Raw data access is possible following review of the proposal keeping with institutional and regulatory guidelines.

## Ethics Statement

This study was reviewed and approved by the Institutional Ethics Committee of CSIR Institute of Genomics & Integrative Biology (CSIR-IGIB) . Ethical approval for patient sample collection and use was obtained from the institutional ethics/review board of all participating hospitals. Patient tumor tissue samples were obtained from institutional biorepositories at the participating centres. Written informed consent had been obtained from all participants at the time of sample collection. All samples and associated clinical data were de-identified prior to analysis. All experiments and procedures were performed in accordance with applicable ethical regulations and guidelines.

## Acknowledgement

We acknowledge the Council of Scientific and Industrial Research (CSIR), India, for funding the CSIR–Indian Breast Cancer Genome Atlas (IBCGA) project (HCP0043). We thank all collaborating institutions and partner hospitals for their continued support. We especially thank all collaborating clinicians for their critical inputs and support staff for their assistance in administering the project across multiple sites. We remain especially grateful to the patients and their families for participating in the project. We also acknowledge the guidance received from the Project Monitoring Committee and CSIR for project management.

