## Supplemental Data for "Whole-Genome Landscape of Breast Cancers from India shows Distinct Clinically Actionable Subtypes"

**Supplementary results**

**Detailed somatic mutational landscape of Indian Breast Cancer Genome (IBCGA) cohort**

The analysis of 501 paired tumour and normal tissue specimens catalogued a total of 4,593,630 somatic mutations. This comprised 59,234 coding mutations - including 39,141 missense, 2,249 nonsense, and 2,640 frameshift/in-frame indels and others (nonstop 102, splice 1,097, startloss 337) - as well as 13,668 silent and 4,534,396 non-coding mutations. The median number of somatic mutations per tumour was 5,209, with a range extending from 158 to 114,814. The variation in mutation burden across the four major subtypes was statistically significant ($p$ = 1.5e−05, ANOVA). The HER2+ (HR+/-) and TNBC tumours demonstrated a significantly higher number of somatic mutations than HR+/HER2-tumours ($p < 0.015$).

The 16 significantly mutated driver genes were identified with the following frequencies: *TP53* (56.49%), *PIK3CA* (38.92%), *KMT2C* (15.17%), *MAP3K1* (14.77%), *GATA3* (7.98%), *TBX3* (7.58%), *NF1* (5.78%), *ERBB2* (5.58%), *RB1* (5.39%), *ARID1A* (4.79%), *CDH1* (4.59%), *PTEN* (3.59%), *SF3B1* (2.79%), *PIK3R1* (2.39%), *CBFB* (2.39%), and *AKT1* (2.00%). In addition, previously established breast cancer drivers such as *RUNX1*, *NOTCH1*, *BRCA1*, *CREBBP*, *BRCA2*, *NCOR1*, *FOXA1*, and *ATRX* were found to be mutated in over 2% of patients. No activating *TERT* promoter mutations were detected in this cohort. Several known hotspot mutations were detected at notable frequencies, including H1047R/L/Y (17.17%), E545K/A (7.98%), and E542K (4.59%) in PIK3CA, and R175H/G (2.59%) and R273H/C/L/P (4.39%) in TP53. No *ESR1* hotspots were identified in these primary tumors, which are often associated with poor outcomes. The PIK3CA E545K mutation was predominantly found in tumours lacking a *TP53* mutation. We also report that 63.26% of *GATA3* mutations were truncating in nature. We compared somatic SNVs found to be present within the 16 significantly mutated genes of the IBCGA cohort against 35 global breast cancer cohorts, totalling 17,706 samples. We observed that frequently mutated genes, such as *TP53* and *PIK3CA*, exhibited a high proportion of known mutations (81% and 82%, respectively) in comparison to global cohorts. In contrast, other genes, including *RB1*, *PIK3R1*, *NF1*, and *KMT2C*, consisted entirely of unique mutations. Further analysis identified significant ($q < 0.2$) enrichment of somatic mutations in the regulatory regions of nine genes, including the promoters of *CCDC107* and *HES1*, which were present in 2-5% of patients.

**Assessment of pathogenicity of somatic driver gene mutations and mutational hotspots**

All missense mutations in *PIK3CA*, *TP53*, *RB1*, *NF1*, *AKT1*, *SF3B1*, *CBFB*, *PTEN*, *PIK3R1*, and 96.5% of *ARID1A*, *ERBB2* and 85.7% of *CDH1* gene were predicted to be pathogenic by the machine learning based framework, on the other hand less proportion of missense mutations in *RB1* (62.5%), *GATA3* (47.3%), *MAP3K1* (29.2%), *KMT2C* (23.0%), and *TBX3* (20.5%) were classified to be deleterious. We found all PIK3CA missense mutations including - D1017H, G106R, G106V, V344G, N1044Y are found to be pathogenic, along with known oncogenic mutations like H1047R, E542K, E545K, N345S etc. Likewise, all TP53 missense mutations that include known oncogenic mutations like R175H, R273H etc. were predicted to be pathogenic.

Somatic hotspot mutations H1047R/L/Y, E545K/A, and E542K in PIK3CA, and R175H/G, R273H/C/L/P, R342*/frameshift, R213*, Y220C, R248W/Q/P/L, p.R282W/G in TP53, E17K in AKT1 genes were detected in the Indian breast cancer cohort. Unlike H1047R, E545K hotspot mutation of PIK3CA was predominantly detected in tumours in absence of mutations in TP53. Overall, the E545K hotspot of PIK3CA was less frequent among both HER2+ and TNBC tumours. We report frequent S133R mutations in MAP3K1 which was exclusively present in HER2- negative tumours irrespective of HR positivity status. Frequent N345K mutation in PIK3CA, K700E in SF3B1, and V7G in NF1 genes were exclusive to HR+HER2- tumours. The particular *NF1* hotspot mutation was found predominantly in tumours with both *TP53*, and *PIK3CA* mutated. Majority of GATA3 mutations detected in this cohort (predominantly found in HR+HER2- tumours) were truncating (63.26%) in nature (frameshift insertion/deletion, splice-site, and start codon). We have noted frequent multiple MAP3K1 mutations in the same tumours.

**Structure-based mutation profiling reveals PIK3CA to harbour unique mutations at previously unknown functional surfaces**

Next, we sought a structure-based understanding of mutations by integrating genomic data with high-confidence protein models, enabling us to identify potential mechanistic effects of key driver mutations. To start with, we focused on missense mutations from 500 patients with annotations of mutation positions and the corresponding transcript. We found mutations in 13,568 genes and at 38,125 unique positions. We compared mutations in the top four genes

(PIK3CA, TP53, MAP3K1, and KMT2C) with the TCGA-BRCA dataset and identified unique mutations (Figure 3A). Among these mutations (n=151), 58.27% occurred at highly conserved residues (Figure 3B), with 52.98% within functional domains, 39.07% predicted to be deleterious, and 46.35% had known disease associations. TP53 and PIK3CA exhibited the highest proportion of these features. In particular, *PIK3CA*, presented 19 unique mutations with similar frequency, amongst which 52.63% are at highly conserved residues. These mutations are also located on the protein functional domains like p85-binding domain (G106V), ras-binding domain (A222D), Phosphoinositide 3-kinase C2 (D352H, D369H, C407W, E418K, E453Q), Phosphoinositide 3-kinase family, accessory domain (Q546E, Q582K) and Phosphatidylinositol 3- and 4-kinase domain (M1010I, E970K) that forms the catalytic domain of the protein. Some of these variants (G1049S, M1043V, C407W, V344G, A222D, G106V) are also likely to destabilise the protein which might interfere with the protein functioning. Around 40% of these mutations (G106V, V344G, E418K, E453Q, Q546E, E970K, M1043V, and G1049S) are also related to several cancer and non-cancerous diseases like CLOVES syndrome, Cowden syndrome, PIK3CA-related disorder, Neoplasm, etc. Apart from these 60% of them are predicted to be deleterious and pathogenic in nature (G106V, A222D, V344G, N345S, D352H, D369H, E418K, Q582K, D1017H, G1049S). These mutating residues are present on the different protein-protein interaction interfaces. Ten mutants (D352, C407, E453, V344, N345, D369, E418, D454, Q546, D1017) are at the interface of three PIK3CA interacting partners (PIK3R1, PIK3R2, PIK3R3), four of them (Q582, E970, M1010, G1049) at the interface of two partners (PIK3R1, PIK3R2) and one mutant, I1058M is at interaction interface of PIK3R3 protein as validated based on pDockQ scores. Molecular docking of PIK3CA and PIK3R1 showed the effect of mutations at the interface. Complexes showed that critical salt bridges, hydrogen bonds were lost in the case of some mutants (N345S, E418K, E453Q, Q546E), as compared to the wild type. This explains the effect of the protein mutations (V344G, N345S, E418K, Q546) on the interface that may perturb the interactions. Our analyses reveal that V344G and E418K are the most crucial mutating residues for PIK3CA function. Lastly, analysis of recurrent mutations revealed high-frequency hits in TP53, MAP3K1, and KMT2C. TP53 variants clustered in the DNA-binding domain (K132R, A138V, P151S, R175G, Y205H, M237I, S241F, R249S/W, E258K, C277F) and tetramerisation motif (R337L), both highly conserved and disease-associated. In MAP3K1, S133R and F14V occurred in 13 and 9 samples, respectively; all MAP3K1 variants (F14V, K32T, V128G, K146Q, D132A, S133R, P257L) localised to unstructured loop regions. KMT2C mutations (F48V, V125I, R886H, P1606L,

Q3836K) were also confined to unstructured regions, with R886H (8 samples) predicted to be highly destabilising and pathogenic. Together, our findings suggest that Indian-specific mutations, especially those in critical domains or flexible regions, points toward selective pressure on regulatory and signalling hubs that differ in Indian breast cancer genomes.

**Quantitative analysis of copy number alterations across breast tumour subtypes**

Genomic analysis identified six chromosome arms (1q, 7q, 8q, 16p, 20p, and 20q) that were significantly amplified and twenty-one arms that were significantly deleted ($q < 0.05$) across the 501 tumors. Significant focal amplifications were quantified for key oncogenes, including *MYC* (25%), *ERBB2* (20%), *ECM1* (15.89%), *PI4KB* (14%), *CCND1* (12%), and *PIK3CA* (8%). Deletions were quantified for tumour suppressors such as *CDKN2A* (6%) and *PTEN* (4%). Subtype-specific patterns were evident, with *ERBB2* amplification occurring in 78.89% of HER2+ tumors. Although somatic mutations in *PIK3CA* were significantly less in TNBCs, the incidence of the copy number amplification of the gene was prevalent. The overall copy number alteration burden was significantly higher in TNBC tumors, with a median proportion of the genome altered of 42.51%, compared to 26.62% in other subgroups ($p = 1.582e{-05}$, t-test). This was driven by both significantly higher amplification ($p = 0.0003$) and deletion ($p = 4.757e{-06}$) events in TNBCs.

**Comparison of somatic mutational and copy number alterations of IBCGA cohort with TCGA breast DCIS**

Somatic mutations in *TP53*, *TBX3*, and *RB1*, in both HR+HER2- and HER2+ (irrespective of HR status) are found to be altered in a significantly higher proportion of patients in the Indian cohort as compared to the similar tumours in TCGA cohort. Mutations in *GATA3* in HER2+ tumours, and *CDH1* in both HR+HER2- and HER2+ tumours were found to be significantly less in this cohort in comparison with the TCGA cohort. In contrast to the TCGA cohort, somatic mutations in *SF3B1* were not found among Indian HER2+ tumours. Frequency of somatic mutations of *TP53* (82%), *PIK3CA* (16%), *PTEN* (10%), *PIK3R1* (6%), *NF1* (6%), and *ERBB2* (3%) was found to be very similar ($p > 0.05$, Fisher's exact test) with the TNBC tumours of TCGA cohort [*TP53*: 83%, *PIK3CA*: 12%, *PTEN*: 7%, *PIK3R1*: 4%, *NF1*: 4%, and *ERBB2*: 3%]. In contrary, we found significantly ($p < 0.05$, Fisher's exact test) higher mutational frequency of *KMT2C* (17%), *ARID1A* (9%), and *MAP3K1* (8%) in TNBC in Indian cohort compared to the TCGA [*KMT2C*: 6%, *ARID1A*: 2%, and *MAP3K1*: 1%]. *AKT1* was found to be mutated in 2% of Indian TNBC patients, while it is not mutated in TNBC patients in TCGA.

Focal amplification of *MYC*, *PIK3CA*, *PI4KB*, *TERC*, *GATA3*, *CCND2*, *EHF*, *ELOVL1*, etc., were common among Indian TNBC tumours and TCGA cohort, in contrary, amplification of *AKR1C3* (in 20% patients of Indian TNBC cohort), *NCEH1* (19%), *PLD1* (17%), *IL10* (16%), *ADIPOR2* (13%), *TERT* (13%), deletion of *BTNL3* (7%) were exclusive to the Indian cohort compared to TCGA. We also noted a lower fraction of Indian TNBC tumours harbouring focal deletion of *PTEN* (19%), in comparison with the TCGA cohort. Amplifications of *ERBB2*, *MYC*, *CCND1*, *ERLIN2*, *PI4KB* etc. were found in HER2+ (irrespective of HR status) tumours from both Indian and the TCGA cohort, while amplifications of *VMP1*, *IL10*, *FGFR1*, *TERT*, *GATA3*, *AKR1C3*, *MDM2*, *ELOVL1*, *TERC*, and *IGF1R* were almost exclusively found in the particular subtype of Indian patients. Interestingly, the frequency of *MYC* amplification was significantly ($p < 0.05$, Fisher's exact test) higher among Indian HR+HER2- tumours (23%) as compared to TCGA (9%). Alongside, amplification of *VMP1*, *FGFR1*, *DECR2*, *BIRC5*, *EHF*, *NCEH1*, *MDM2*, *AKR1C3*, *ADIPOR2*, *LSS*, *TERT* etc. were almost exclusive to the particular subtype among Indian patients compared to TCGA.

**Genomic correlates of distant metastasis**

In the 36 patients who developed distant metastasis, primary tumours showed a significantly higher frequency of copy number amplifications in *MYC* (47% vs. 23% in non-metastatic) and *ERBB2* (42% vs. 18%). Somatic mutations in *CDH1* were also significantly higher in tumours that developed metastasis (14%) compared to those that did not (4%). Furthermore, the TP53 hotspot mutation R175H was significantly enriched among metastatic tumours (8%) compared to non-metastatic cases (2%) ($p < 0.05$, Fisher's exact test).

**Signatures of genomic instability and telomere dynamics**

Of the 16 somatic mutational signatures identified, SBS3, which is linked to DNA double strand break repair (DSBR) failure / homologous recombination deficiency (HRD), had a median contribution of 14.78% across the cohort. The contribution of SBS3 was significantly higher in TNBCs (median contribution of 28.33%) compared to hormone-positive tumours (11.74%) ($p < 5.043\text{e}{-14}$, *t*-test), and its prevalence was significantly associated with *TP53* mutational status ($p = 1.494\text{e}{-10}$, *t*-test).

The estimated tumour telomere length ranged from 1.31Kb to 12.60Kb, with a median of 2.94Kb. The relative telomere length compared to paired normal tissue varied significantly among subtypes ($p$ = 0.0019, Kruskal-Wallis test). Telomere shortening was significantly associated with mutations in *TP53* and *KMT2C* ($p < 0.0068$, Wilcoxon test), while elongation was associated with mutations in *PIK3CA*, *MAP3K1*, and *TBX3* ($p < 0.018$). Telomere shortening was also significantly correlated with the proportion of the genome altered (Pearson's correlation coefficient = -0.3, $p$ = 1.746e−11).

**Germline alteration and somatic non-coding mutational landscape**

We detected rare germline mutations (excluding "benign" reported in ClinVar database) in breast cancer associated DNA repair genes - BRCA1 (in 5% patients), TP53 (3%), BRCA2 (2%) to be present in ≥ 2% patients and *CHEK2*, *RAD50*, *RAD51D*, *MUTYH*, and *ATM* to be present between 1-2% Indian breast cancer patients. While rare germline mutations in *BRCA2* and *TP53* were significantly (p < 0.05, Fisher's exact test) enriched in TNBC tumours, rare germline *BRCA2* mutations were predominantly detected in HER2+ tumours (irrespective of HR status). We found only 2.5% of patients with reported family history of cancer harboured pathogenic rare germline alterations in *BRCA1*, *BRCA2*, or *TP53* genes and 0.8% of the patients had rare germline alteration in *CHECK2*, *RAD51D*, *MUTYH* or *ATM* genes.

We found significant ($q < 0.2$) enrichment of somatic mutations in regulatory regions of 9 genes - in promoters of *CCDC107*, *EIF2S3L*, *PRDM2*, *HES1*, in UTRs of *TAGAP*, *TRIM50*, both promoter and UTR (3' or 5' untranslated region) of *WDR74*, and TSS (transcription start site) of *RMRP*, *MTND2P21*. These mutations were present in 2-5% of the patients in this cohort. 85.71% promoter mutations in *EIF2S3L* were detected in the HR+/HER2- tumours.

**Somatic structural variation landscape in breast cancer cohort**

We identified 90,555 somatic structural variations (SVs) encompassing 438 patients in our cohort, of which 59,078 (65.2%) represented local intrachromosomal breaks (<5 Mb), although intrachromosomal (20.2%; 18,331) and interchromosomal (14.5%, 13,144) genomic reshuffling events (>5 Mb) were also noted. Although, HR+HER2+ tumours had the highest SV burden (median: 214 SVs per patient), followed by TNBC (157 SVs), HR-HER2+ (131 SVs), and HR+HER2- (69 SVs) tumours, we noted subtype-specific variations in the type of SVs as, TNBC tumours had the highest burden of duplication events (median: 37 per patient), whereas, HR+HER2+ had highest burden of deletion (50 SVs), inversions (94 SVs), and translocations (32 SVs). We identified pathogenic deletion and duplication events (ACMG classification), which showed high tumour allele fractions (TAF $\geq$ 10%). HER2+ tumours showed a higher burden of pathogenic events (median: 74 per patient), followed by TNBC (38 pathogenic SVs) and HR+HER2- (29 pathogenic SVs), indicating possible clinical actionability. We integrated the large-scale copy-number patterns (G-scores) along with the SV calls to construct a genomic interaction map between 5Mb genomic segments. We noted enrichment of super-enhancers in breast cancer and normal breast epithelium, in the regions flanking 1Mb of ERBB2 (median 76 kb upstream and 143 kb downstream), which might disrupt enhancer-promoter interactions, TAD boundaries, and chromatin organisation. Recurrent SVs included genes, like, *ASIC2* (22% of HER2+ patients), *MACROD2* (9.5% of patients), *RAD51B* (12% of TNBC patients), as well as, several chromatin remodelers (total 466 SVs across 159 patients), like, *NF1* (23% of patients), *ARID1B* (4% of patients), *ESR1* (3.8% of patients), *RB1*, *BRD4*, etc.

A large fraction of SV breakpoints (50-53%) were enriched in protein-coding genes and their 2-kb upstream regions. SV breakpoints showed a preferential overlap with known fragile sites annotated in the HumCFS database (Z-score = 19.196, $p < 0.001$) (1). Genes proximal to fragile sites and frequently disrupted include *TTC28, IKZF3, MACROD2, ASIC2*, and *CDK12*, among others. Comparison with PCAWG confirmed recurrent alterations in these and other genes, reflecting both shared and cohort-specific SV patterns.
We further observed tumour subtype-specific SV patterns: more duplications in TNBC and inversions in HR+/HER2+ subtypes, with deletions common across all subtypes. COSMIC mutational signatures analysis also showed differential distribution of non-clustered SVs across subtypes: TNBC had more small deletions/duplications, HR+/HER2+ had large inversions/translocations. The signature SV3, known to be associated with homologous recombination deficiency, was enriched in TNBC samples, while the SV2 signature of unknown etiology was prevalent in HER2+ samples (2). We found that the top 25% of samples with high cosine similarity to the SV3 signature showed significantly better survival probability within the TNBC cohort (log-rank $p < 0.05$). Significant differences were noted in the structural variation landscape of HRD-high versus HRD-low patients ($p < 0.001$, Wilcoxon Rank Sum Test).

**PAM50 intrinsic subtype composition highlights distinct patterns in IBCGA**

We evaluated the distribution of PAM50 intrinsic molecular subtypes across the IBCGA cohort and compared it with three major external breast cancer datasets (TCGA,

METABRIC, and SCAN-B). Subtype assignments were generated using the AIMS classifier (3). A global comparison of subtype frequencies across cohorts demonstrated a highly significant difference in overall distribution (global $p < 0.01$).

Within the IBCGA cohort, Basal tumours constituted the largest subtype, accounting for 38.4% of cases. This proportion was notably higher than that observed in TCGA (17.5%), METABRIC (16.7%), and SCAN-B (9.3%). In contrast, Luminal A tumours represented only 14.3% of IBCGA cases, compared with 36.1% in TCGA, 27.1% in METABRIC, and 48.3% in SCAN-B (**Figure S13A, Table S13**).

Luminal B tumours comprised 19.5% of the IBCGA cohort, a frequency comparable to TCGA (18%) but lower than METABRIC (27.8%) and higher than SCAN-B (11.7%). HER2-enriched tumours accounted for 20.8% of IBCGA cases, similar to TCGA (20.4%) and exceeding the proportions observed in METABRIC (14.9%) and SCAN-B (11.6%). Normal-like tumours were the least frequent subtype in IBCGA (7%), compared with 8% in TCGA, 13.5% in METABRIC, and 19.1% in SCAN-B (**Figure S13A, Table S13**).

Overall, the intrinsic subtype landscape of the Indian IBCGA cohort is characterized by a marked predominance of Basal-like tumours and a reduced representation of Luminal A disease relative to predominantly Western cohorts, underscoring important population-level differences in tumour biology.

**TNBC molecular subtype distribution across cohorts relative to IBCGA**

Given the high prevalence of triple-negative breast cancer (TNBC) within the IBCGA cohort, we next examined TNBC-specific transcriptional heterogeneity using three established molecular classification frameworks: TNBCType-4 (4), TNBCType-6 (5), and the Burstein subtype scheme (6). Subtype distributions were compared across IBCGA, TCGA, METABRIC, SCAN-B, and FUSCC cohorts. Global comparisons were performed for each classification model.

Under the TNBCType-6 model, subtype composition differed significantly across cohorts (global $p < 0.01$). In the IBCGA cohort, IM (21.9%) and UNS (23.1%) were among the most frequent subtypes, followed by BL1 (14.1%) and LAR (14.1%). Compared with other cohorts, IBCGA demonstrated a higher proportion of UNS tumours relative to TCGA (10.1%), METABRIC (11.5%), SCAN-B (11.9%), and FUSCC (12.7%). The mesenchymal

(M) subtype was less frequent in IBCGA (12%) than in TCGA (22.7%) and SCAN-B (18.4%), while MSLtumourswere comparatively lower in IBCGA (4.9%) than in most external datasets.(**Figure S13C, Table S15**) In contrast, the TNBCType-4 model showed no significant difference in overall subtype distribution across cohorts (global $p = 0.135$). Within IBCGA, BL1 represented the largest group (32.1%), followed by BL2 (19.8%), LAR (18.4%), and M (17%). These proportions were broadly comparable to those observed in TCGA, METABRIC, SCAN-B, and FUSCC, indicating a conserved subtype structure across populations under this framework. (**Figure S13D, Table S16**)

The Burstein classification revealed a highly significant difference in subtype distribution across cohorts (global $p < 0.01$). In IBCGA, BLIA was the predominant subtype, comprising 64.7% of TNBC cases - substantially higher than TCGA (44.7%), METABRIC (54.8%), SCAN-B (45.8%), and FUSCC (38.4%). Conversely, BLIS tumours accounted for only 14.8% of IBCGA cases, compared with markedly higher proportions in TCGA (38.2%) and SCAN-B (30.6%). The MES subtype was relatively rare in IBCGA (2.1%) compared with METABRIC (10.9%) and FUSCC (12.5%), while LAR frequencies in IBCGA (18.4%) were comparable to SCAN-B (18.7%) but lower than FUSCC (24.1%) (**Figure S13B, Table S14**).

Overall, these findings indicate that TNBC transcriptional heterogeneity within the Indian IBCGA cohort depends on the classification framework applied. While the Lehmann TNBCType-4 model demonstrates broadly conserved subtype architecture, the TNBCType-6 and Burstein schemes reveal significant shifts in subtype composition, particularly the marked enrichment of BLIA and reduced BLIS representation in IBCGA. These results underscore population-level differences in TNBC biology and highlight the importance of context-specific molecular stratification.

**Evaluation of Batch Effect and Normalization**

Post-normalization analyses demonstrated a substantial reduction in variance associated with technical factors, including batch effects, library size, and tumour purity, while enhancing variance attributable to biologically relevant signals such as PAM50 subtypes. PCA performed before normalization revealed pronounced batch effects, with samples clustering predominantly by sequencing site, indicative of strong technical bias. Following normalization, this batch-driven separation was resolved, and samples no longer clustered by sequencing origin. Importantly, normalization also improved biological resolution: samples of the same PAM50 subtype clustered more closely together, and distinct separation between subtypes became more apparent in PCA space. These results confirm that the normalization strategy effectively mitigated technical artifacts while preserving and amplifying meaningful biological variation. **(Figure S14)**

**Transcriptional Sub-clustering Reveals Two Distinct TNBC Subtypes with Different Biological Programs**

Consensus clustering of 140 clinically defined TNBC tumours based on the top 3,500 most variable genes identified two major transcriptional subgroups: LAR-like TNBC (N=39) and Basal-like TNBC (N=101), which showed strong concordance with PAM50 and TNBC4-subtype (5)) classifications. The LAR-like subgroup exhibited elevated expression of hormone receptor genes (AR, PGR, ESR1, ERBB2, EGFR) and significant enrichment of metabolic pathways, including oxidative phosphorylation, fatty acid metabolism, adipogenesis, peroxisome activity, and hormone response signatures (estrogen and androgen response), while showing downregulation of interferon gamma and interferon alpha pathways that may contribute to worse survival outcomes. In contrast, the Basal-like subgroup was characterized by enrichment of proliferative and inflammatory programs such as mitotic spindle assembly, MYC targets, G2M checkpoint, E2F targets, interferon responses (gamma and alpha), inflammatory response, IL2-STAT5 signalling, IL6-JAK-STAT3 signalling, and TNFα signalling via NF-κB ($q < 0.05$) **(Figure S16A) (Table S23)**. Kaplan-Meier survival analysis revealed that patients with LAR-like TNBC experienced significantly worse recurrence-free survival compared to those with Basal-like TNBC (log-rank $p = 0.039$), with this divergence becoming more pronounced over time. Multivariable Cox proportional hazards regression analysis demonstrated that LAR-like TNBC classification remained independently associated with increased recurrence risk ($p = 0.047$) after adjustment for age group and lymph node status, supporting the independent prognostic relevance of these transcriptional subtypes beyond traditional clinicopathological variables. **(Figure S16A)**

**Differential immunoregulation and cell abundance**

We identified differential immunoregulation at the pathway level across BC subtypes using single-sample Gene Set Enrichment Analysis (ssGSEA), in which significant enrichment of

T-cell-mediated immunity, allograft rejection, and interferon response pathways was observed in Basal-like tumours irrespective of HR/HER2 status ($q < 0.05$; one-tailed Wilcoxon rank-sum test). Consistent with these findings, both mRNA expression of lineage-specific markers (*CD8A, CD8B, CD4, CD3E* and *CD163*) and immune deconvolution revealed higher putative infiltration of macrophages and CD4+ T-cells in Basal-like tumours (*p*adj < 0.05; one-tailed Wilcoxon rank-sum test). The HR+/HER2- samples classified as Basal-like by the PAM50 schema showed significant enrichment of immune pathways similar to that of TNBCs, unlike HR+HER2- samples classified as Luminal-A ($q < 0.05$). A majority of Basal-like samples scored above the median for the proliferative (92%; $p < 0.05$; One-vs-Rest Fisher's exact test), wound healing (87%; $p < 0.05$; One-vs-Rest Fisher's exact test), CSF1 response (73%; $p < 0.05$; One-vs-Rest Fisher's exact test), and interferon gamma (64%; $p > 0.5$; One-vs-Rest Fisher's exact test) signatures. On the contrary, the HR+HER2- samples that were classified as either Luminal-A or Normal-like by the PAM50 schema showed significant enrichment of Epithelial-Mesenchymal Transition (EMT) and Angiogenesis pathways, and these samples also had correspondingly higher TGF-beta score and elevated expression of collagen and ECM glycoproteins ($q < 0.05$; one-tailed Wilcoxon rank-sum test).
The four transcriptionally distinct sample clusters obtained by Iterative Consensus Clustering revealed pronounced differences in the tumour Microenvironment (TME) within the IBCGA cohort. The BaPro (C1) cluster, comprising mostly TNBCs (61%) and HR+HER2- (24%) exhibited significant upregulation of IL6-JAK-STAT3, IL2-STAT5, and Interferon Gamma pathways. In contrast to other sample clusters, 92% of the BaPro (C1) cluster samples presented a basal-like transcriptional profile along with significant enrichment of onco-immunological signatures like CSF1 response/macrophage, Leukocyte infiltration, and interferon response (Thorsson et al., 2018). The immune cell abundance estimates showed that the BaPro (C1) cluster had a high ratio of M1 relative to M2 macrophages and a higher infiltration of T-lymphocytes relative to total lymphocytes. A small subset of TNBC samples was clustered independently in HER2-androgenic (C4), and these samples were molecularly distinct from the TNBC dominant cluster BaPro (C1) due to **i**) increased HER2 transcriptional activity ($p < 0.05$), **ii**) lower enrichment of Interferon and JAK-STAT signalling pathways ($q < 0.05$), and **iii**) higher representation of Luminal Androgen Receptor (LAR) subtype samples (16/19; 84%) ($p < 0.05$; two-sided Fisher's exact test). Among the BaPro (C1) sample cluster, we found a subset of TNBC samples (15/104; 14%) that were characterized by high proliferation, low score of interferon signatures, and fewer immune infiltrates, and these were classified as Basal-like Immune Suppressed (BLIS) subtype by

Burstein classification The LumPro (C2) and LumDiff (C3) clusters consisted of mostly HER2- samples (~75%) but differed in their molecular subtypes. LumPro (C2) and LumDiff (C3) clusters were comprised of Luminal-B and Luminal-A (& normal-like) samples, respectively. Compared to all other clusters, including LumPro (C2) cluster, the LumDiff (C3) cluster scored significantly higher on TGF-beta signalling, stromal content, ECM glycoproteins, and collagen content ($q < 0.05$).

**Immune cell transcriptional factor and cytokine profile across BC subtypes**

Using annotations from Human Protein Atlas and KEGG resources, we identified the immune cell-specific transcriptional factors and cytokines, and these were used to profile BC samples. We noted significant overexpression of transcriptional factors (*STAT1, RUNX3, STAT4*) involved in the differentiation of T-helper cells in the TNBC samples, whereas the transcriptional factors closely associated with estrogen receptor signalling that were also involved in the regulation of T-cell commitment (*GATA3*) were upregulated in the HR+/HER2- cohort ($q < 0.05$; one-tailed Wilcoxon rank-sum test). In addition, the HR+HER2- samples showed overexpression of genes belonging to the nuclear receptors superfamily (*RXRA, RARA*) that were involved in the polarization of monocyte-derived macrophages (7) . Also, the transcriptional factors involved in the differentiation and function of TH17 (*RORC*) and Tregs (*FOXP3*) cells were also significantly overexpressed in HR+HER2- ($q < 0.05$; one-tailed Wilcoxon rank-sum test). On further evaluation using the KEGG pathview maps, we found that transcriptional factors and kinases involved in the differentiation of Th1 and Th2 cells (*JAK2, JAK3, STAT1, STAT4, NOTCH1*) were upregulated in TNBC and indicative of active CD4+ T-cell polarization driven by JAK-STAT signalling. The activation of interferon-responsive JAK-STAT signalling (*STAT1*) and HIF1a-mediated oxidative stress response (*HIF1A*) in TNBC tumours may be driving *PDL1* expression in TNBC tumours through autocrine and paracrine mechanisms ($p$adj < 0.05) (7), (8), (9) . Moreover, the cytokines associated with Th2-cell function (*IL5*) were upregulated in the HR+HER2-cohort, whereas cytokines associated with Th1 response (*IFNG, IL12A*) were significantly overexpressed in TNBC samples ($q < 0.05$; one-tailed Wilcoxon rank-sum test).

The consensus clustering further revealed differences in the expression profile of transcriptional factors within subtypes. Although the Th17-related transcriptional factor

RORC was significantly upregulated in both HR+HER2- dominant LumPro (C2) and LumDiff (C3) clusters, the significant overexpression of the regulator of CD4+ regulatory T-cells (*FOXP3*) was limited to LumDiff (C3). Similarly, the TNBC-dominant clusters- BaPro (C1) and HER2-androgenic (C4) clusters both shared the upregulation of HIF1a, but only BaPro samples showed overexpression of transcriptional factors involved in JAK-STAT (*STAT1*, *STAT4*) signalling. The overexpression of cytokines involved in JAK-STAT signalling (*IFNG*, *IL12A*, and *IL12B*) in the BaPro (C1) cluster further supports this observation.

**Supplementary figure and legends**

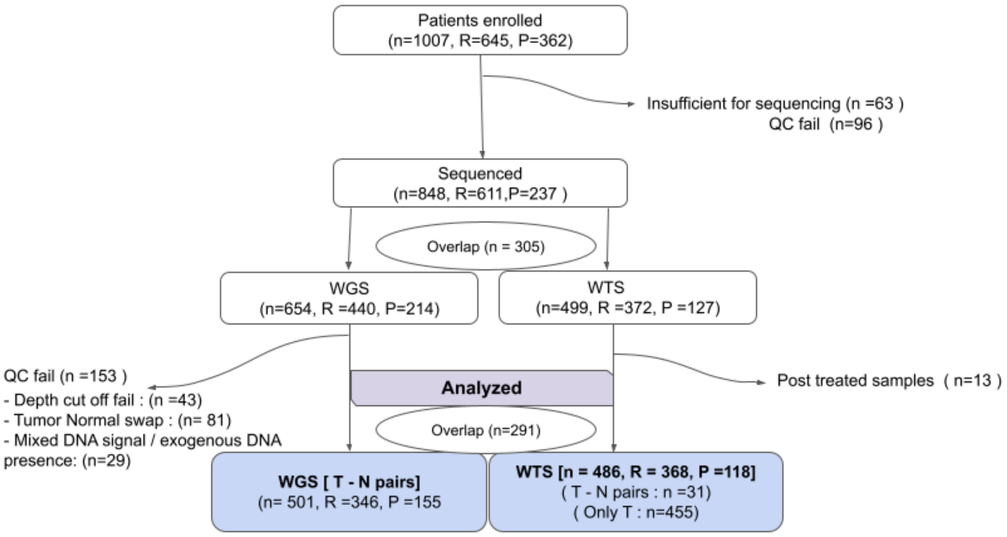


Figure S1: The CONSORT diagram summarizes patient enrollment, sample processing, sequencing, and downstream analysis, with participants stratified into prospective (P) and retrospective (R) cohorts. The diagram shows sample inclusion, quality control filtering, sequencing modalities (WGS and WTS), and the final datasets used for analysis.

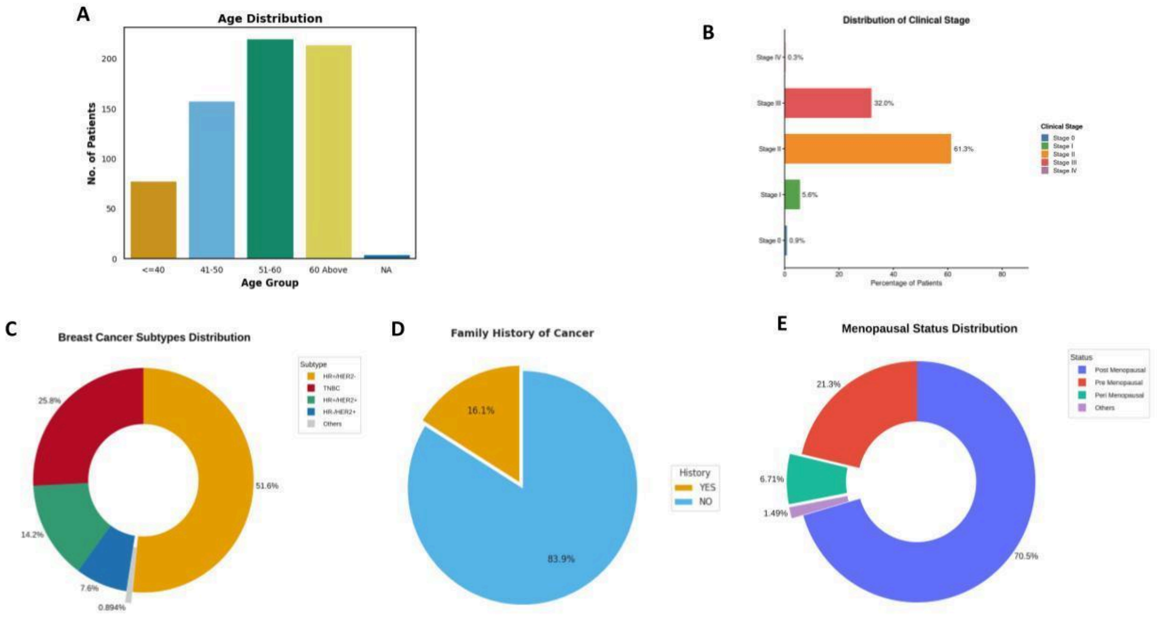

Figure S2: Clinicopathological characteristics of the Indian breast cancer cohort. (A) Age distribution of patients at diagnosis, (B) Distribution of cancer stages at presentation, (C) Molecular subtypes of breast cancer, (D) Percentage of patients with a reported family history of cancer, (E) Menopausal status of the cohort.

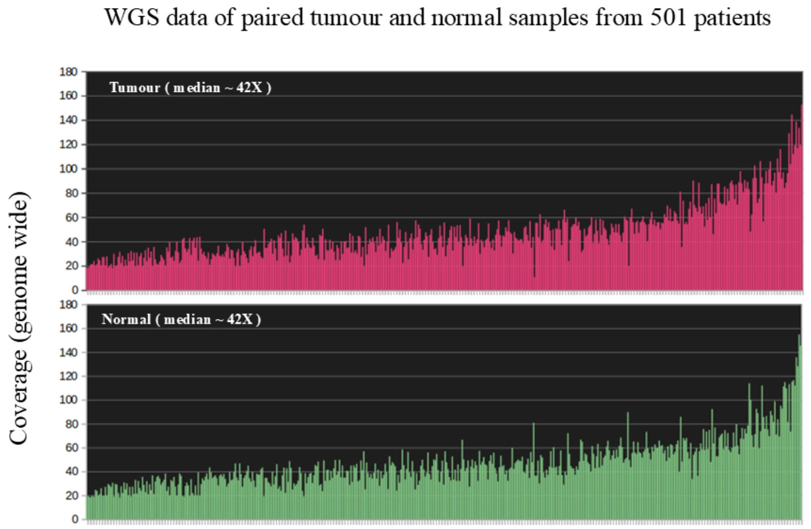


Figure S3: So Out of 501 patients 337 (67%) patients have both tumour and normal depth of coverage ≥ 30X. 113 (22%) have both tumour and normal depth of coverage ≥ 50X and 39 (8%) have both tumour and normal depth of coverage ≥ 70X.

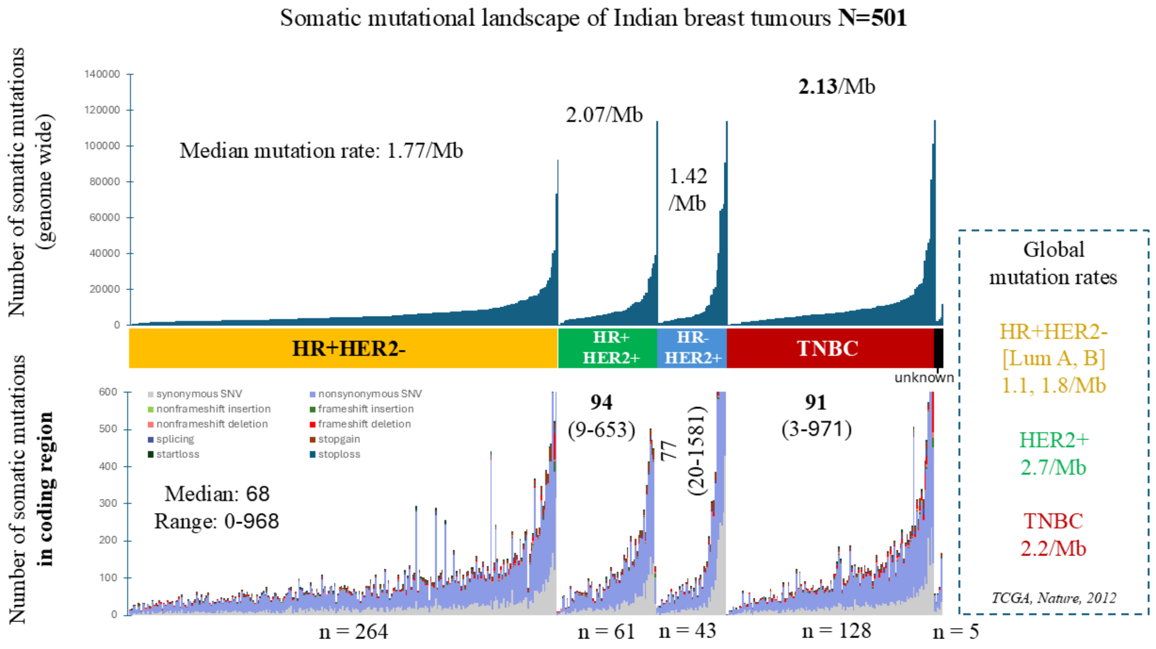


Figure S4: The somatic mutation rate (calculated as mutation per Mb of genome) varied significantly (p = 1.5e-05) between subtypes, HR+HER2-, HR+HER2+, HR-HER2+, and

TNBC patients harboured 1.42/Mb, 2.07/Mb, 1.78/Mb, and 2.13/Mb genome wide somatic mutation rate with highest obtained for TNBC tumours. The somatic mutation rate of breast tumour from TCGA is shown in right.

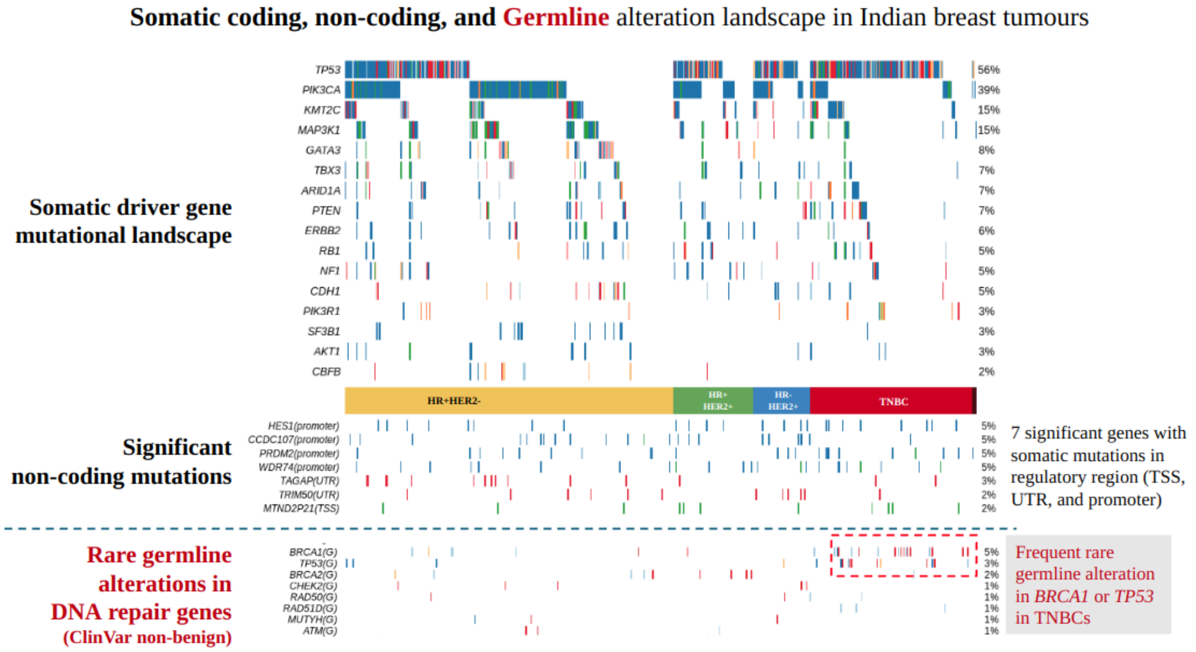


Figure S5: Somatic (coding significantly mutated genes, significantly altered noncoding mutations), and germline (rare germline pathogenic or likely pathogenic alterations in previously known breast cancer associated genes altered over 1% patients). [Represented: 16 significantly mutated (somatic) genes (SMGs) mutated in ≥ 5% of the tumours in any subtype, and ≥ 2% in the overall breast cancer cohort]

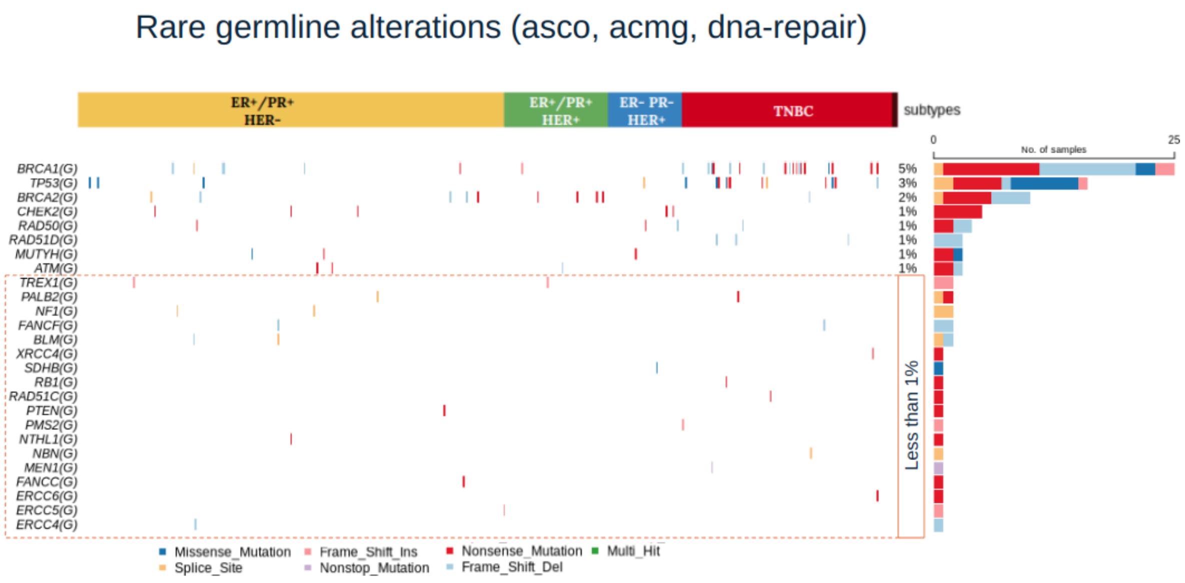


: Germline pathogenic or likely pathogenic mutational profile of DNA repair genes from ASCO and ACMG in IBCGA cohort.

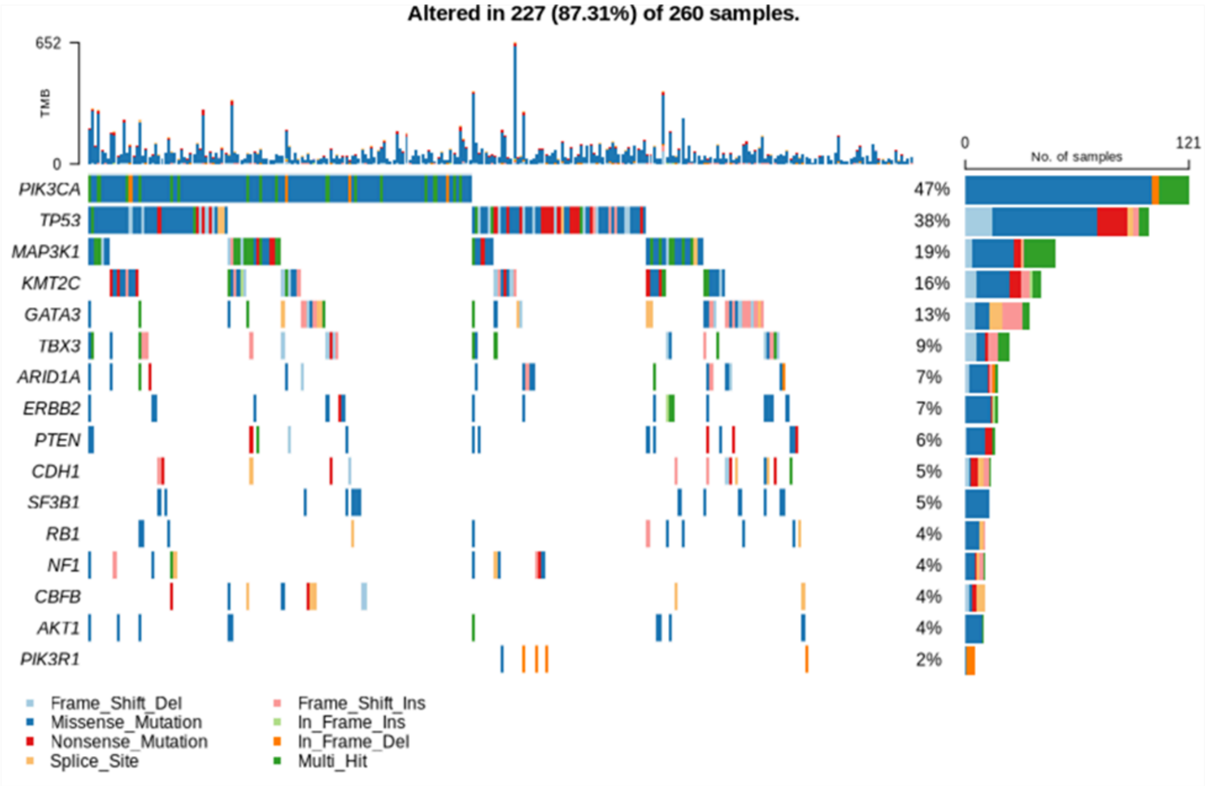


Figure S7: Somatic mutational landscape of 260 HR+HER2- patients. A median of 4542 somatic mutations ranging between 158 and 92699. [Represented: 16 significantly mutated (somatic) genes (SMGs) mutated in ≥ 5% of the tumours in any subtype, and ≥ 2% in the overall breast cancer cohort]

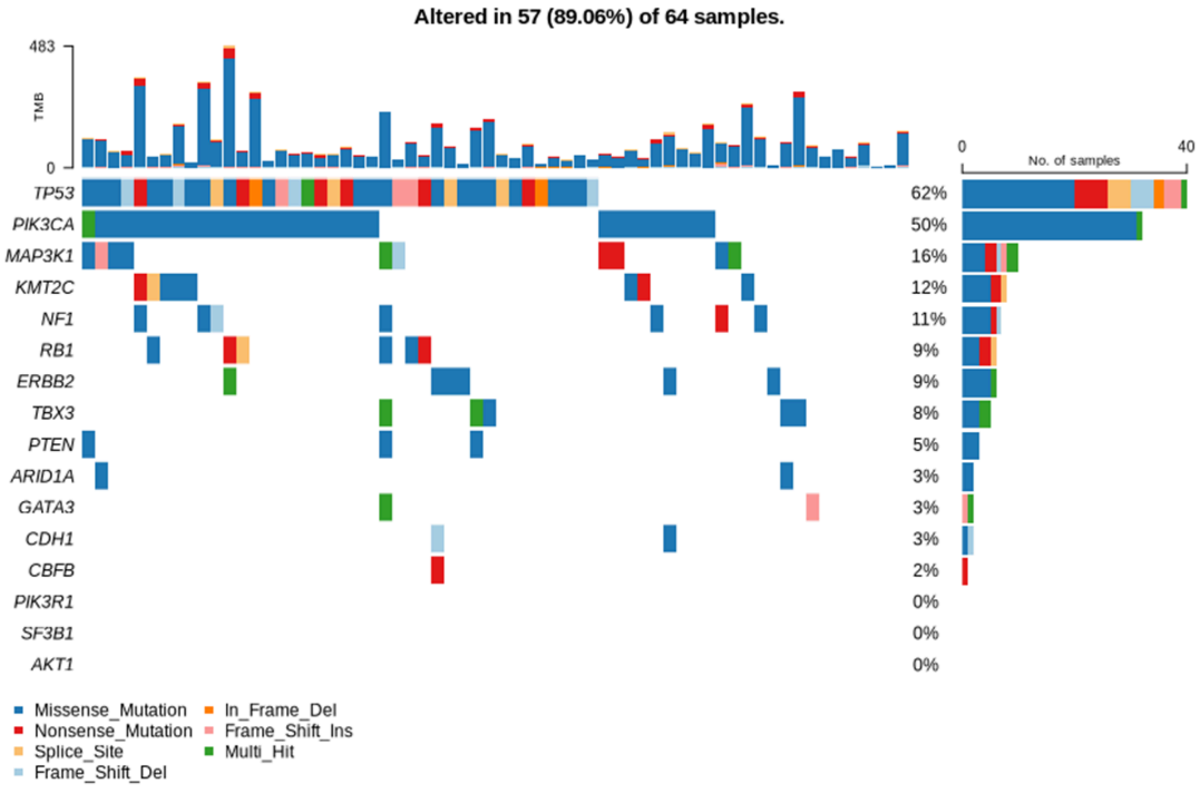


Figure S8: Somatic mutational landscape of 64 HR+HER2+ patients. A median of 6632 somatic mutations ranging between 653 and 114273. [Represented: 16 significantly mutated (somatic) genes (SMGs) mutated in ≥ 5% of the tumours in any subtype, and ≥ 2% in the overall breast cancer cohort]

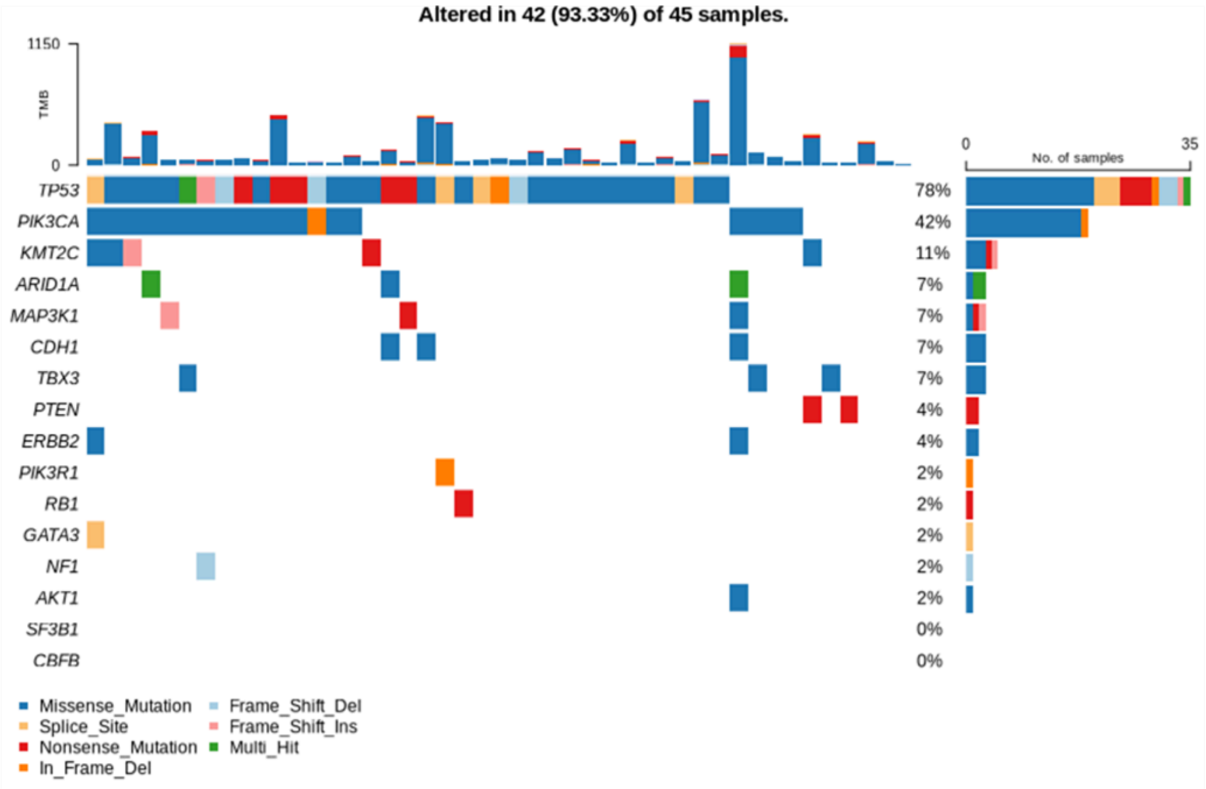


Figure S9 Somatic mutational landscape of 45 HR-HER2+ patients. A median of 5690 somatic mutations ranging between 1241 and 114214. [Represented: 16 significantly mutated (somatic) genes (SMGs) mutated in ≥ 5% of the tumours in any subtype, and ≥ 2% in the overall breast cancer cohort]

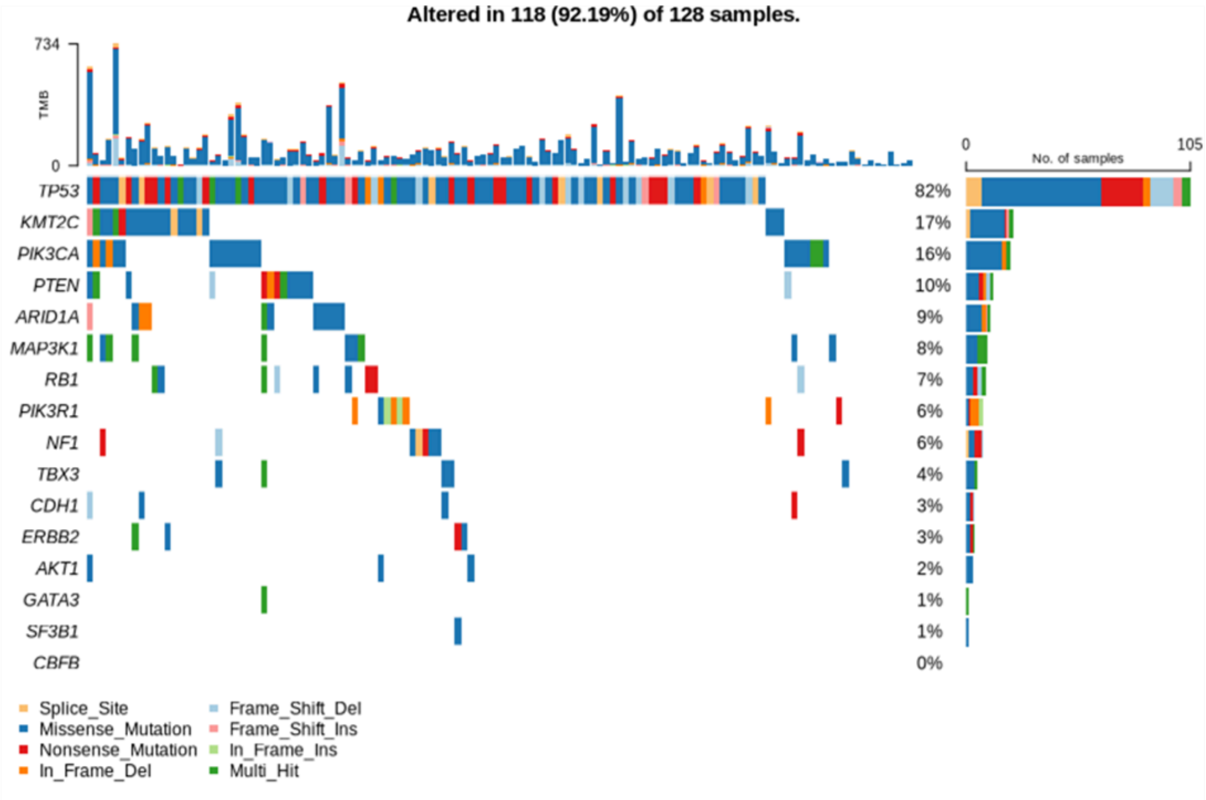


Figure S10: Somatic mutational landscape of 128 TNBC patients. A median of 6812 somatic mutations ranging between 256 and 114814. [Represented: 16 significantly mutated (somatic) genes (SMGs) mutated in ≥ 5% of the tumours in any subtype, and ≥ 2% in the overall breast cancer cohort]

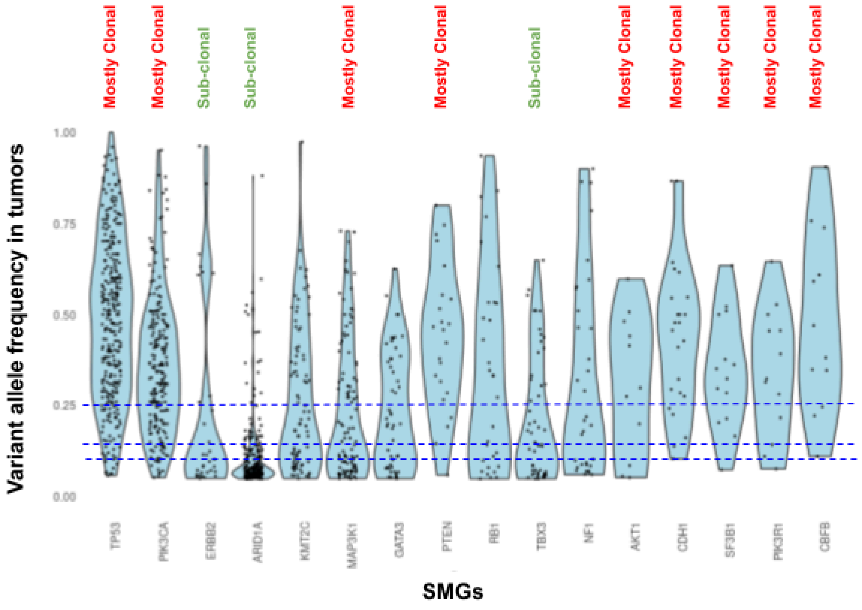

Figure S11: Clonal architecture of driver genes in the IBCGA cohort derived from variant allele frequencies of the somatic mutations (capped at 0.05 lower threshold). [Represented: 16 significantly mutated (somatic) genes (SMGs) mutated in ≥ 5% of the tumours in any subtype, and ≥ 2% in the overall breast cancer cohort]

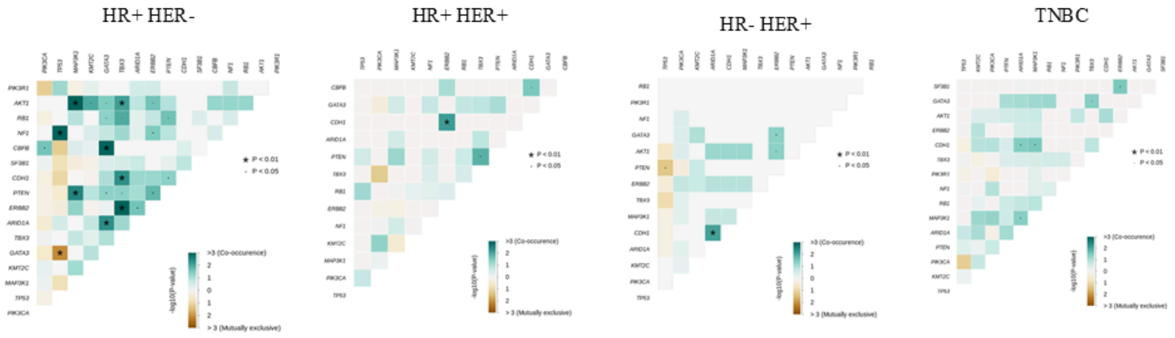


Figure S12: Co-occurrence and mutual exclusivity of somatic alterations in four subgroups of breast cancer patients. Significant co-occurrence of GATA3 with CBFB, ERBB2 with TBX3, or CDH1 mutations was found in this cohort, while somatic mutations in TP53 with GATA3 or MAP3K1, PIK3CA with PIK3R1, etc., were mutually exclusive. [Represented: 16 significantly mutated (somatic) genes (SMGs) mutated in ≥ 5% of the tumours in any subtype, and ≥ 2% in the overall breast cancer cohort]

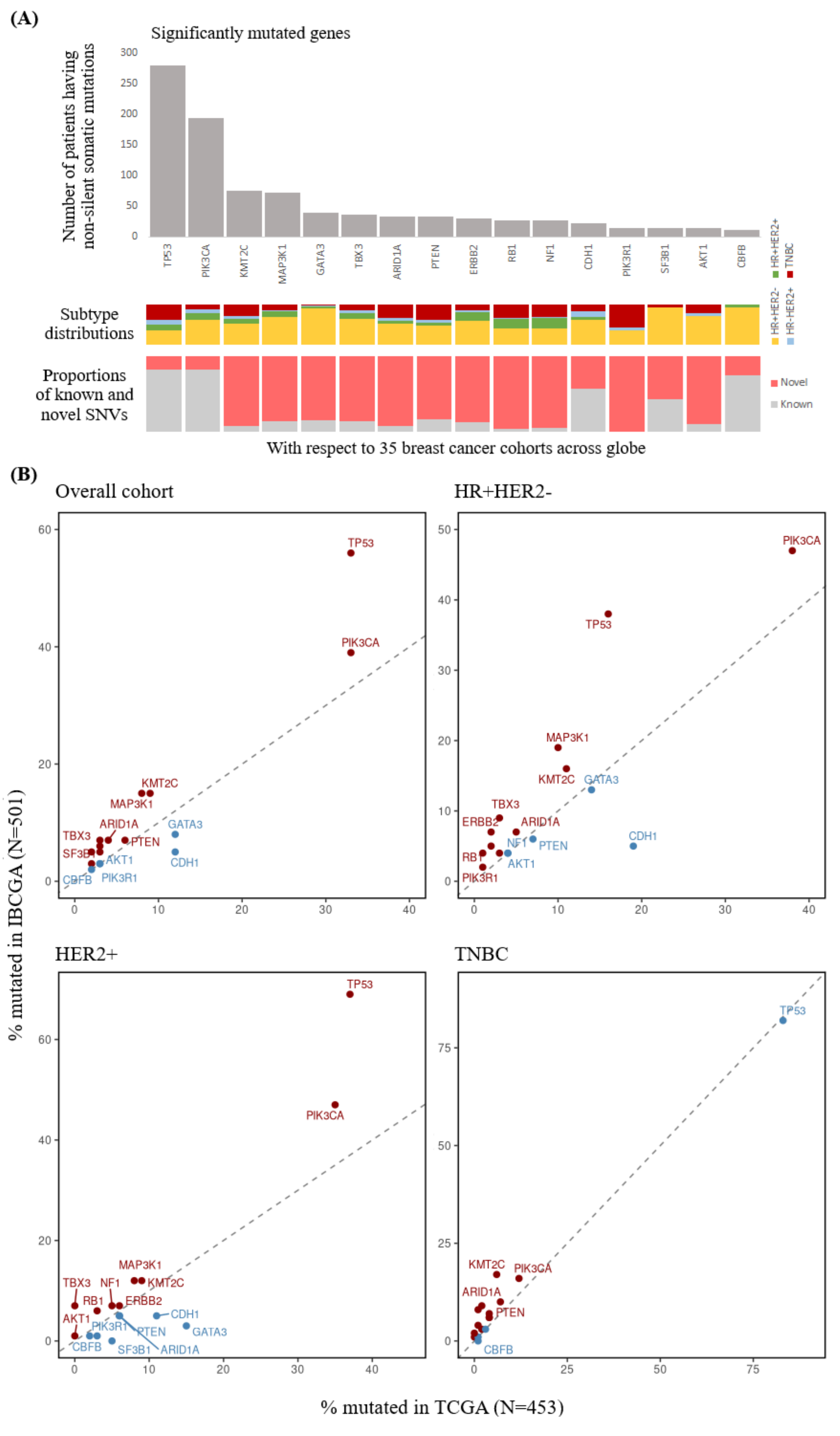


Figure S13: (A) The somatic SNVs in significantly mutated genes in IBCGA cohort is compared with 35 global breast cancer cohorts. (B) Comparison of somatic mutational landscape of HR+HER2- breast tumours from Indian cohort with TCGA breast cancer cohort.

Somatic mutations in TP53, TBX3, and RB1, in both HR+/HER2- and HER2+ (HR+/-) are found to be altered in significantly higher proportion of patients in Indian cohort as compared to the similar tumours in TCGA cohort. Mutations in GATA3 in HER2+ tumours, and CDH1 in both HR+HER2- and HER2+ tumours were found to be significantly less in this cohort in comparison with the TCGA cohort. In contrast to the TCGA cohort, somatic mutations in SF3B1 were not found among Indian HER2+ tumours. Frequency of somatic mutations of TP53 (82%), PIK3CA (16%), PTEN (10%), PIK3R1 (6%), NF1 (6%), and ERBB2 (3%) was found to be very similar ($p > 0.05$, Fisher's exact test) with the TNBC tumours of TCGA cohort [TP53: 83%, PIK3CA: 12%, PTEN: 7%, PIK3R1: 4%, NF1: 4%, and ERBB2: 3%]. In contrary, we found significantly ($p < 0.05$, Fisher's exact test) higher mutational frequency of KMT2C (17%), ARID1A (9%), and MAP3K1 (8%) in TNBC in Indian cohort compared to the TCGA [KMT2C: 6%, ARID1A: 2%, and MAP3K1: 1%]. AKT1 was found to be mutated in 2% of Indian TNBC patients, while it is not mutated in TNBC patients in TCGA. [Represented: 16 significantly mutated (somatic) genes (SMGs) mutated in $\geq$ 5% of the tumours in any subtype, and $\geq$ 2% in the overall breast cancer cohort]

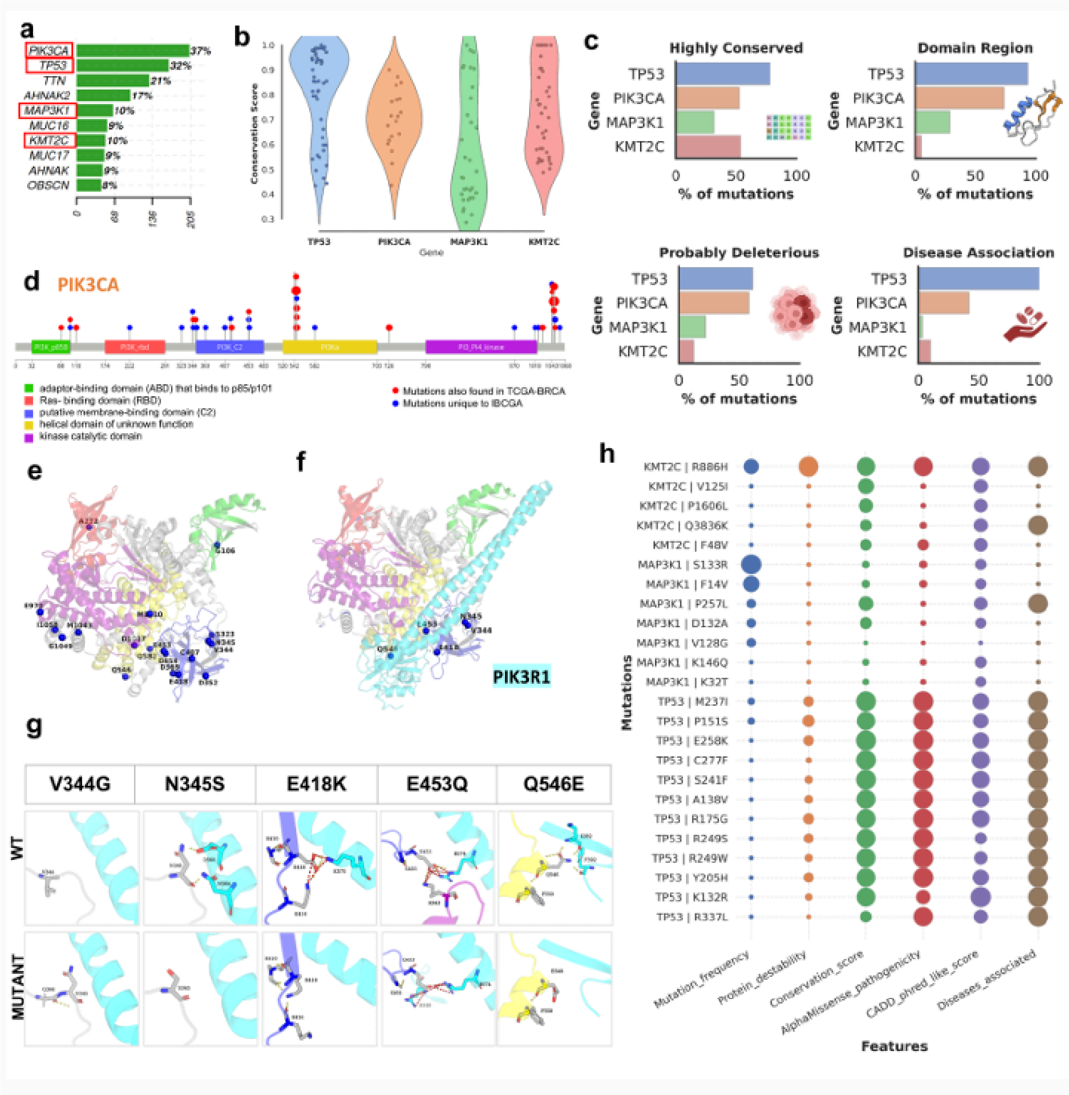


Figure S14: (a) The horizontal bar graph shows the top 10 genes that harbors major missense mutations, amongst which *PIK3CA*, *TP53*, *MAP3K1* and *KMT2C* are well known breast cancer driver genes. (b) The horizontal bar plots show the percentage of unique IBCGA mutations localized at the highly conserved residue position, present in the functional domain region, predicted to be probably deleterious (CADD Phred-like score >25.0), and having disease associations in case of the top four genes. (c) The lollipop plot depicts the mutation on the PIK3CA and TP53 protein. The size of the circle represents the comparative frequency of the mutations. The mutations that are unique to IBCGA and not reported in TCGA-BRCA are shown in red. (d) Unique mutations from IBCGA are highlighted and labelled on the PIK3CA protein in red. (e) Interaction of PIK3CA wildtype and mutants with the PIK3R1(cyan) protein. The residue interaction at the interface shows the loss of interaction in mutants (V344G, N345S, E418K, Q546E) as compared to wild type. The dashes represent the hydrogen bond (yellow) and salt bridge interactions (red).

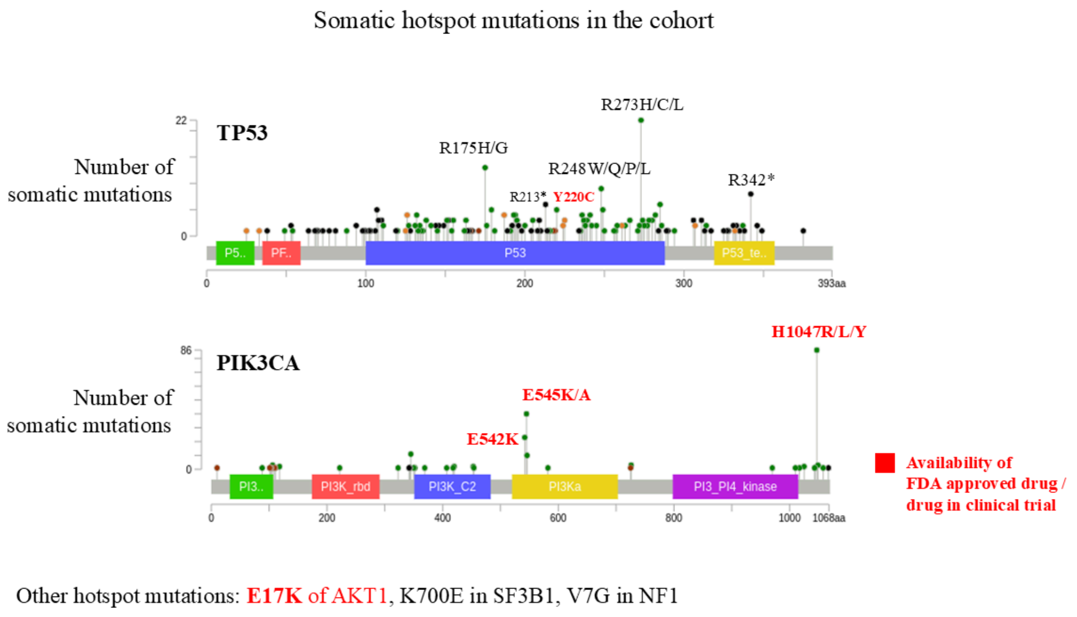


Figure S15: Prevalence of somatic hotspot mutations in Indian breast cancer cohort. FDA approved drugs are available for multiple of these hotspot mutations in PIK3CA, and Y220C of TP53, and E17K of AKT1 genes.

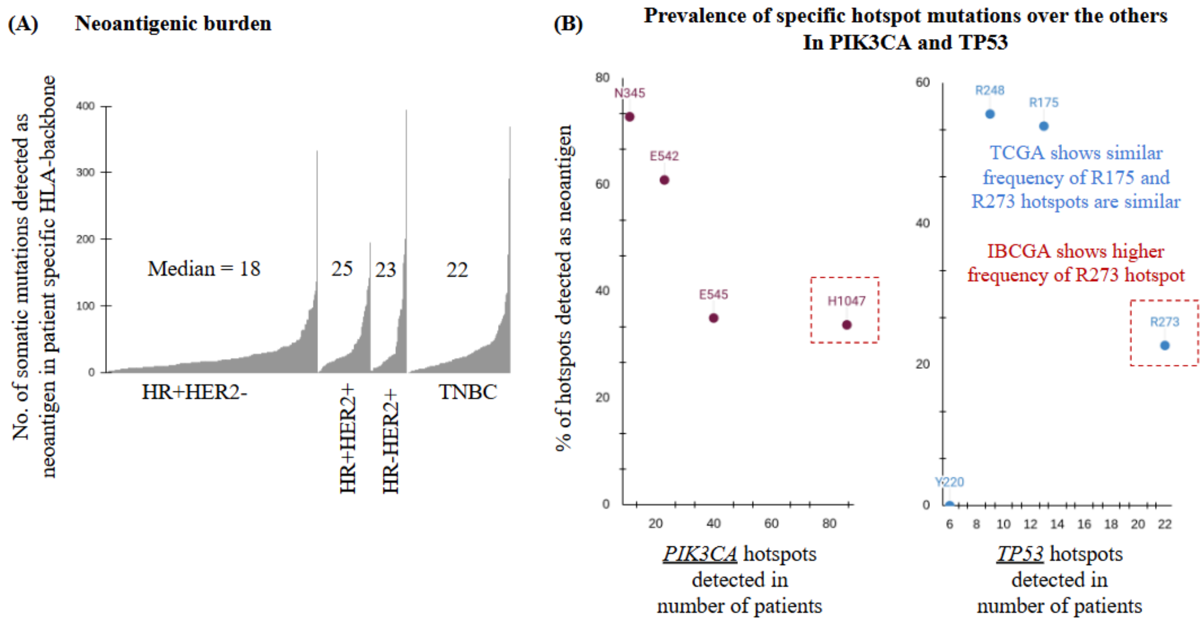


Figure S16: Landscape of neoantigenic somatic mutations in breast tumours from Indian patients. (A) Distributions of neoantigenic somatic mutations (on patient-specific MHC-I backbone), among breast cancer IHC subtypes. (B) Prevalence of specific hotspot mutations in PIK3CA and TP53 are shown to be correlated with immune escape capability. Immune escape capability is defined such that it has an inverse correlation of the proportion of mutations detected as neoantigen in the number of patients.

A

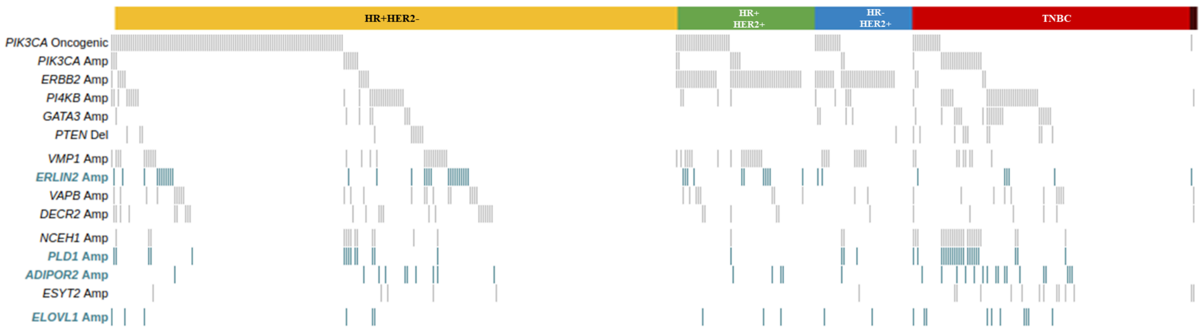


B

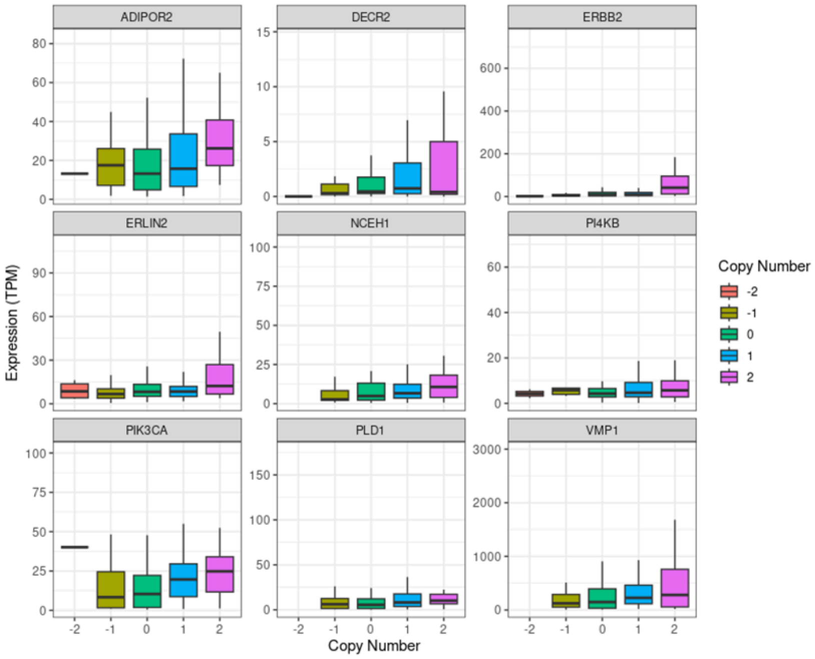


Figure S17: (A) Landscape of somatic alterations that activate the PI3K-AKT pathway and copy number amplification genes that are directly involved in lipid metabolism in breast tumors. Four genes highlighted are involved in “de novo fatty acid synthesis” (shown in blue colour). (B) Correlation of copy number with gene expression in breast tumors.

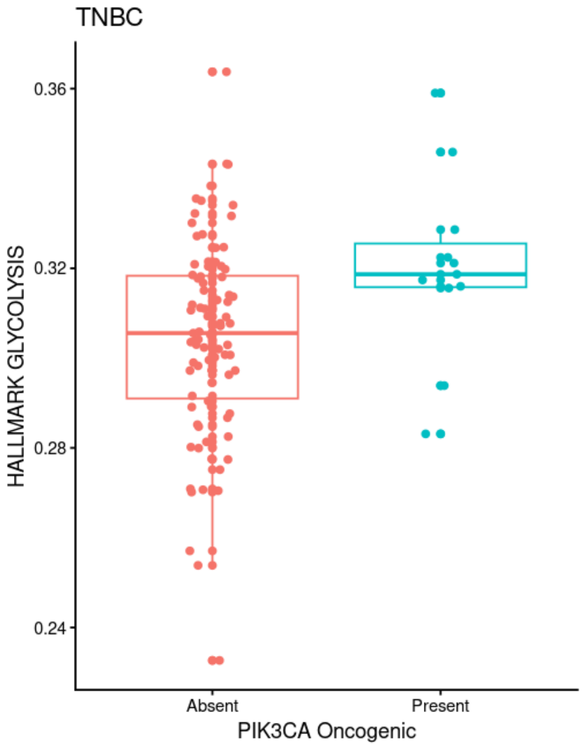


Figure S18: ssGSEA scores across hallmark pathways in TNBC samples with either PIK3CA somatic mutations or GATA3 CNV amplification.

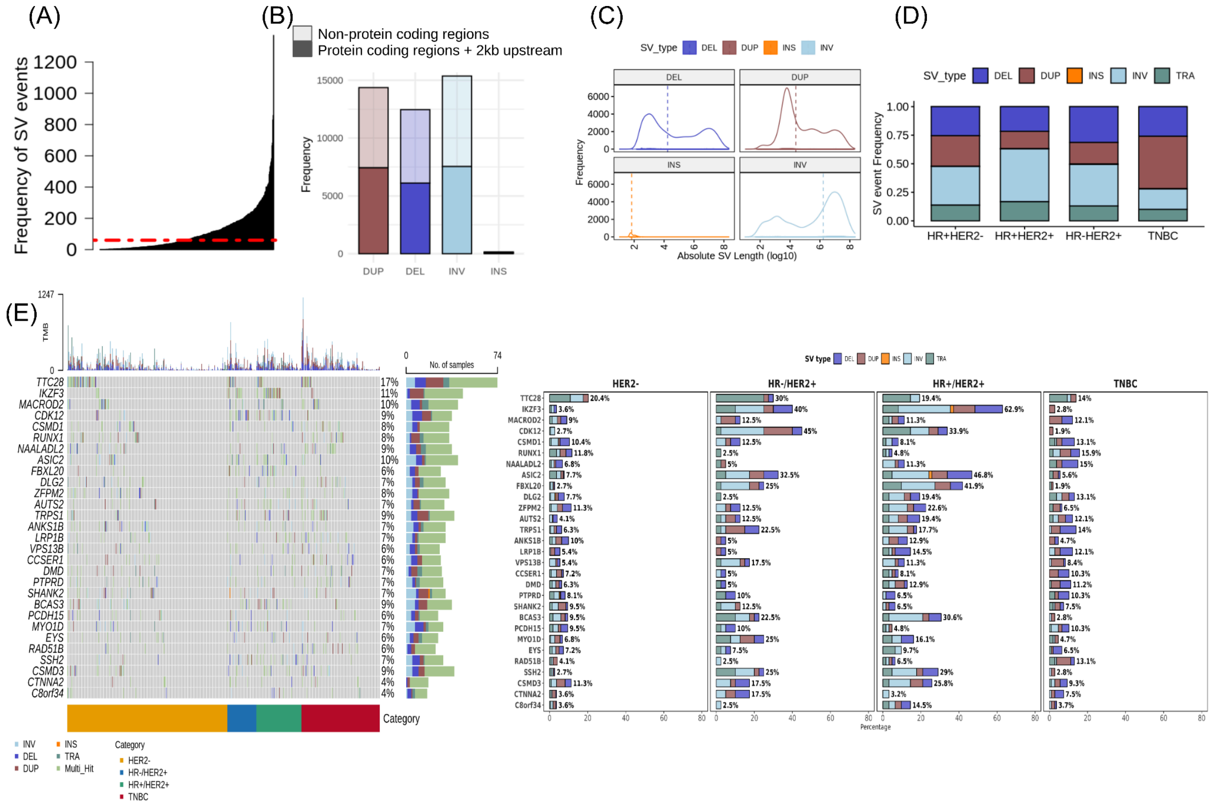


Figure S19: Landscape of structural variants (SVs) across breast cancer subtypes. (A)

Distribution of SV event frequency across all samples, highlighting their overall occurrence. (B) Illustrates the distribution of SV overlap with 2kb upstream of protein coding genes and non-coding regions with respect to SV types, the distribution for [Duplication (DUP), Deletion (DEL), Inversion (INV), Insertion (INS)], the distribution Other regions: DEL (6355), DUP (6939), INS (92), INV (7829), 2kb upstream of protein coding region: DEL (6108), DUP (7428), INS (64), INV (7539). (C) Length distributions for major SV classes (DEL, DUP, INS, INV) Log-scaled density plots depict the distribution of absolute SV lengths. (D) Shows the proportion of each SV type across cancer subtypes (HER2-, HR+/HER2-, HR+/HER2+, TNBC). This illustrates how the SV frequencies vary for each type like HER2+ and TNBC show enrichment for specific SV types. (E) Left: Oncoplot depicting SV events for 30 recurrently altered genes, stratified by breast cancer subtype and SV type. Top bars represent tumour mutational burden (TMB). Right: Barplots of subtype-specific frequency of SVs in each gene, colored by type.

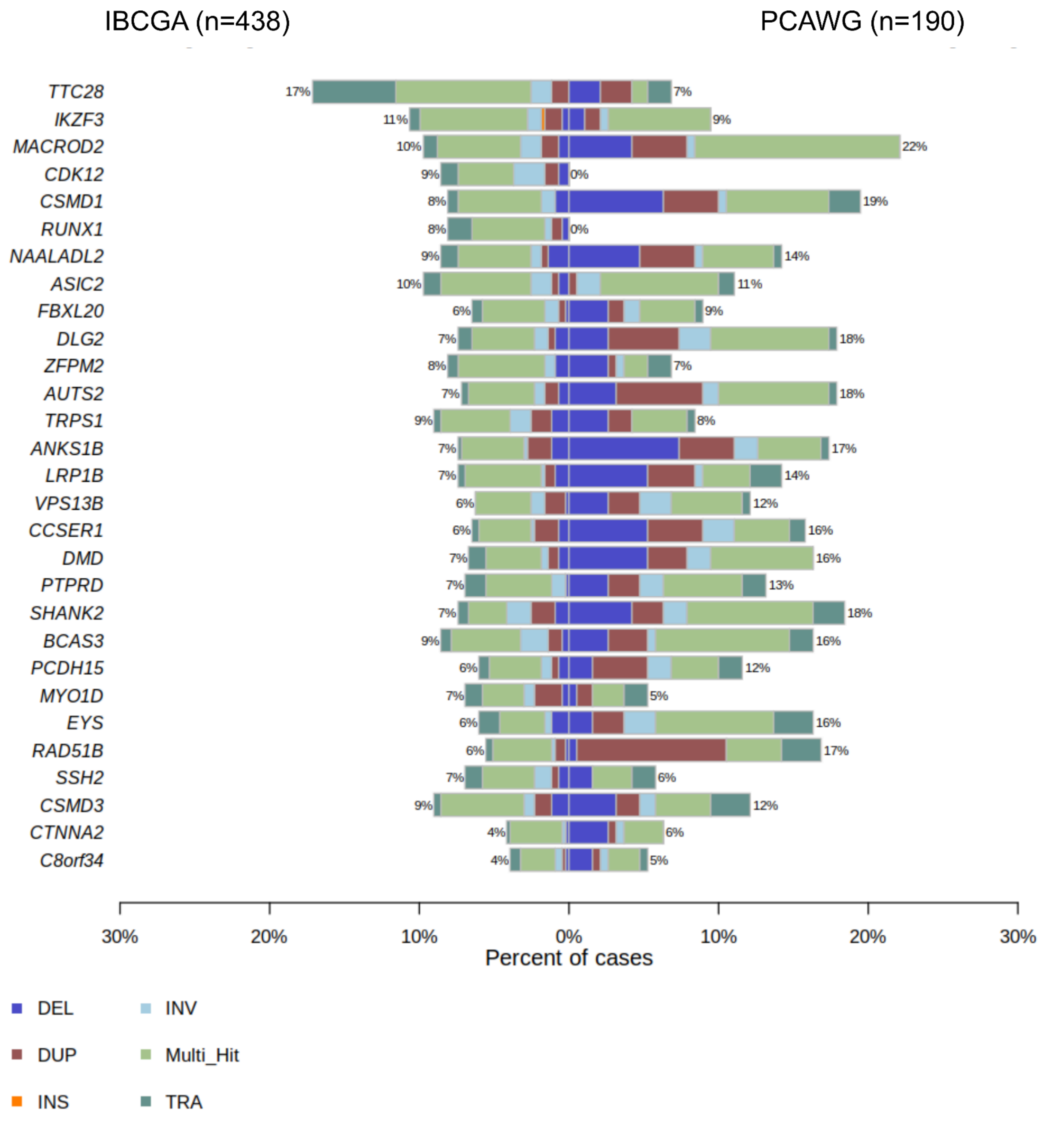


Figure S20: This co-barplot compares the distribution and recurrence of structural variants (SVs) in breast cancer samples from the IBCGA and PCAWG cohorts for the 30 recurrently altered genes by SVs, highlighting shared and cohort-specific patterns.

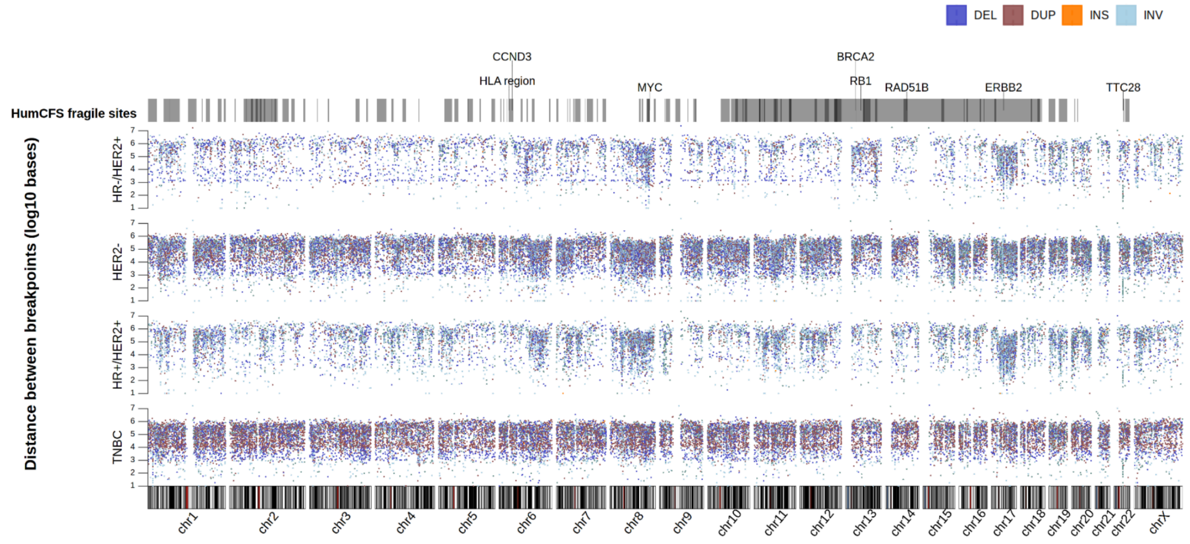


**(B)**

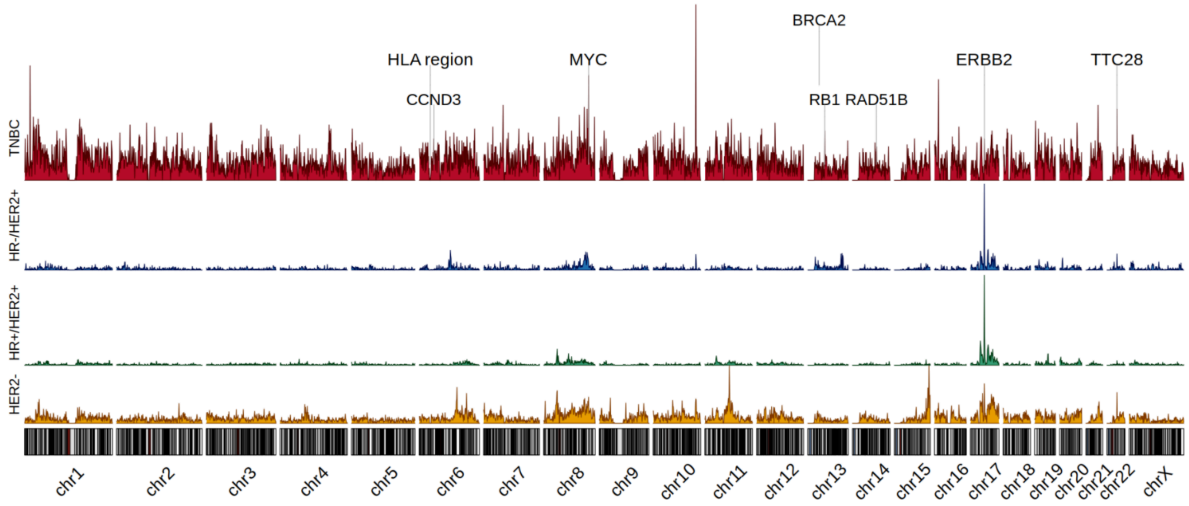


Figure S21: (A) Genome-wide distribution of structural variant breakpoints across subtypes. (B) Density tracks stratified by SV type highlight subtype-specific enrichment of breakpoints across chromosomes and subtypes. Top vertical track: represents annotated common fragile sites.

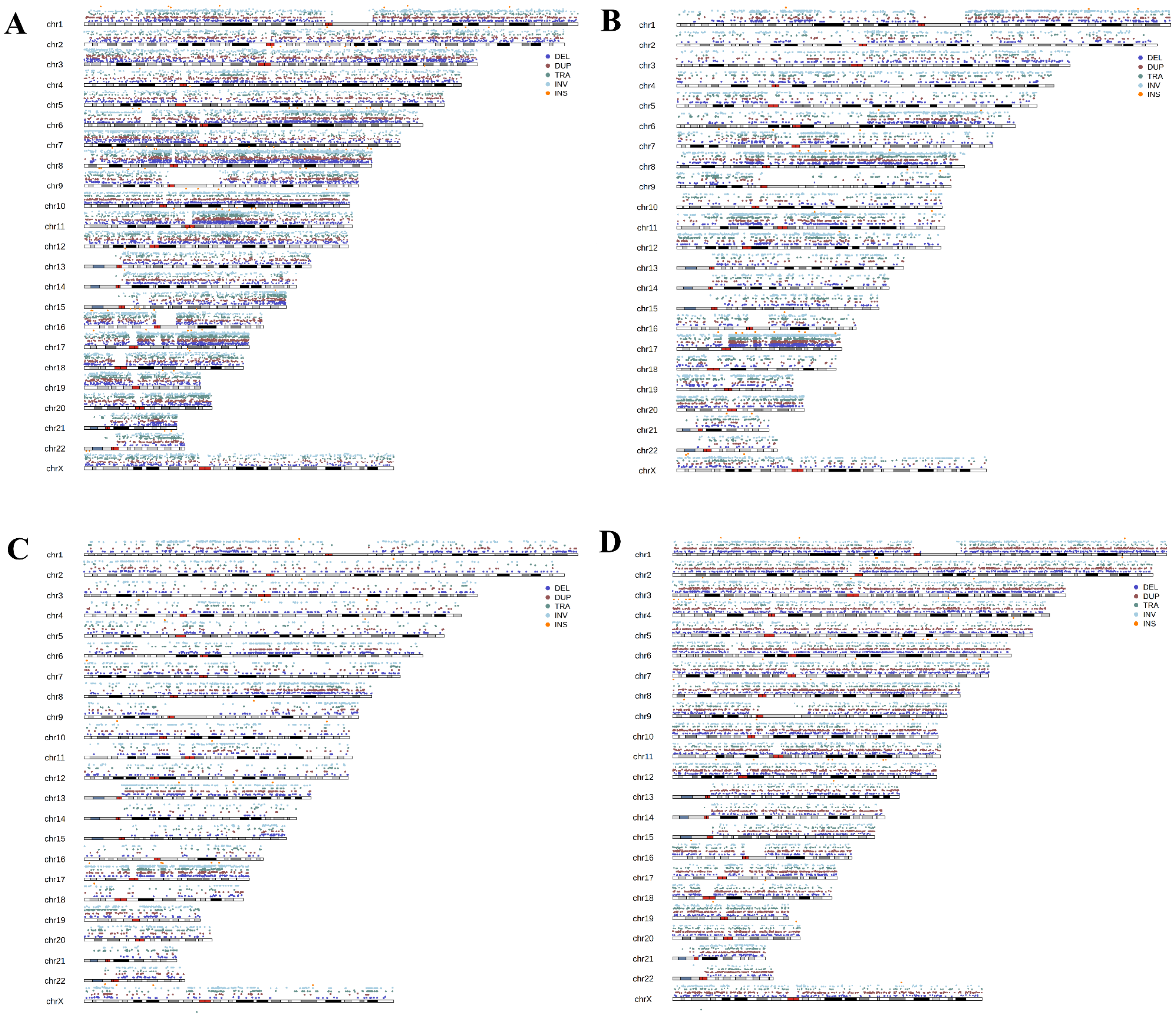


Figure S22:The karyoplots represents distribution of structural variant breakpoints along each chromosome for IHC subtypes - (A) HR-HER2+, (B) HR+HER2+, (C) HR+HER2-, and (D) TNBCs.

(A)

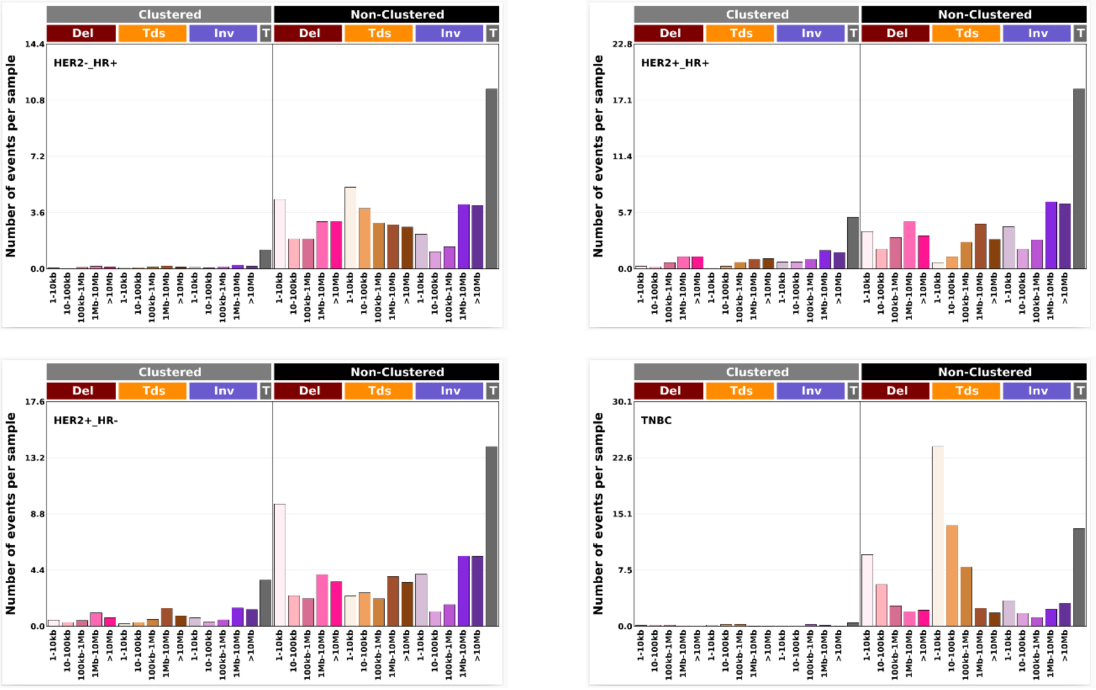


Figure S23: (A) Top bar plots represent the SV events per subtype categorised as per COSMIC SV32 classification schema.

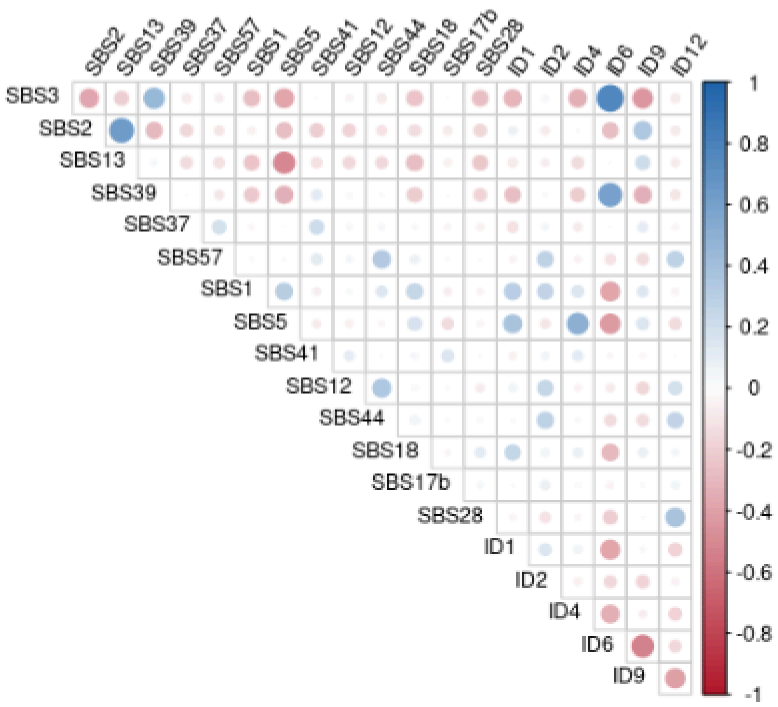


Figure S24: Inter correlations between somatic mutational single base substitutions (SBS) and insertion deletion (ID) signatures. SBS2 and SBS13 signatures related to APOBEC hyperactivity were positively correlated. The homologous recombination defect (HRD) related signatures SBS3 and ID6 showed highest positive correlation.

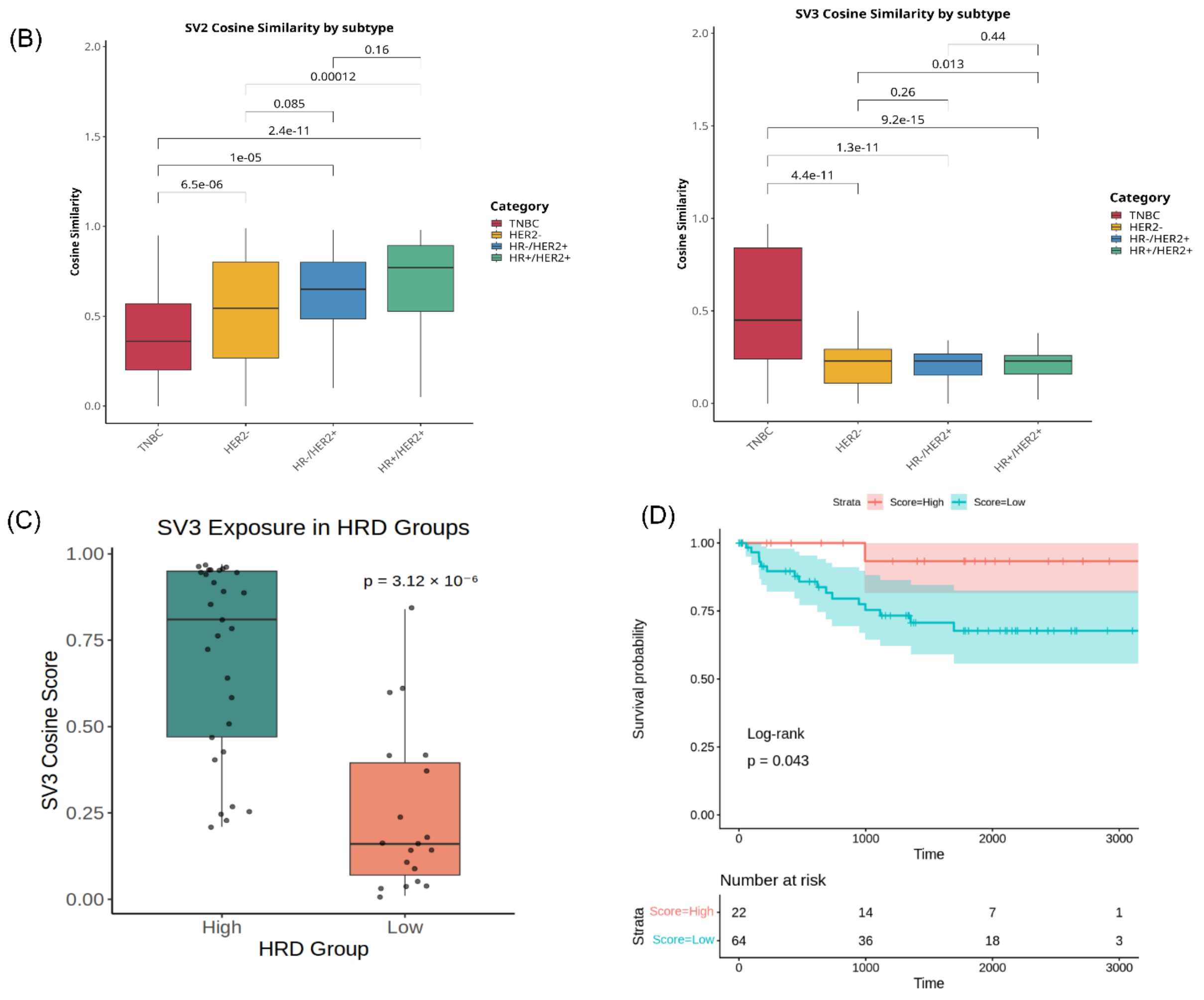


Figure S25: (B) Box plots represent the cosine similarities of SV2 and SV3 signatures with respect to all the 4 subtypes. (C) bottom left box plot represents the SV3 cosine scores stratified by HRD status, SV3 contribution is significantly higher in HRD-high tumours compared to HRD-low tumours (Wilcoxon test, $p$ = 3.12 × $10^{-6}$). (D) Kaplan-Meier curves show disease-free survival probability comparing the samples with high cosine similarity (upper quartile; cosine similarity ≥Q3) with the remaining of the cohort (< Q3) within the TNBC cohort.

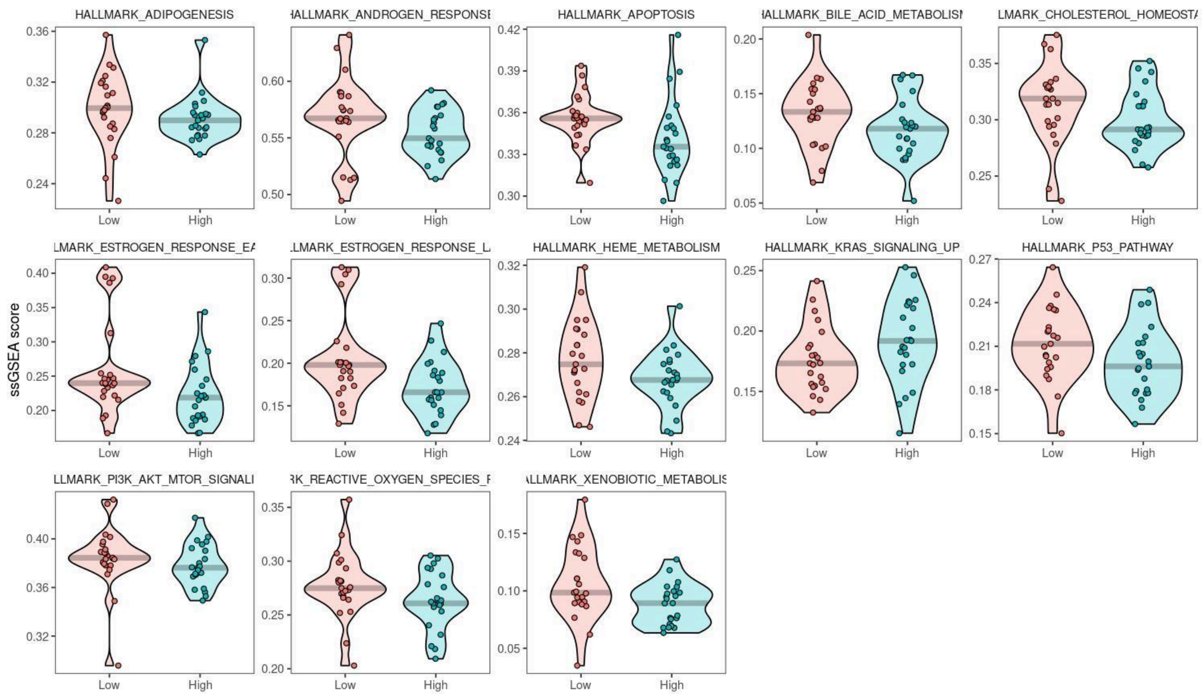


Figure S26: Significantly different ssGSEA enrichment of hallmark pathways in presence of high (> Q3), and low (< Q1) SBS3 relative abundance in TNBC tumours.

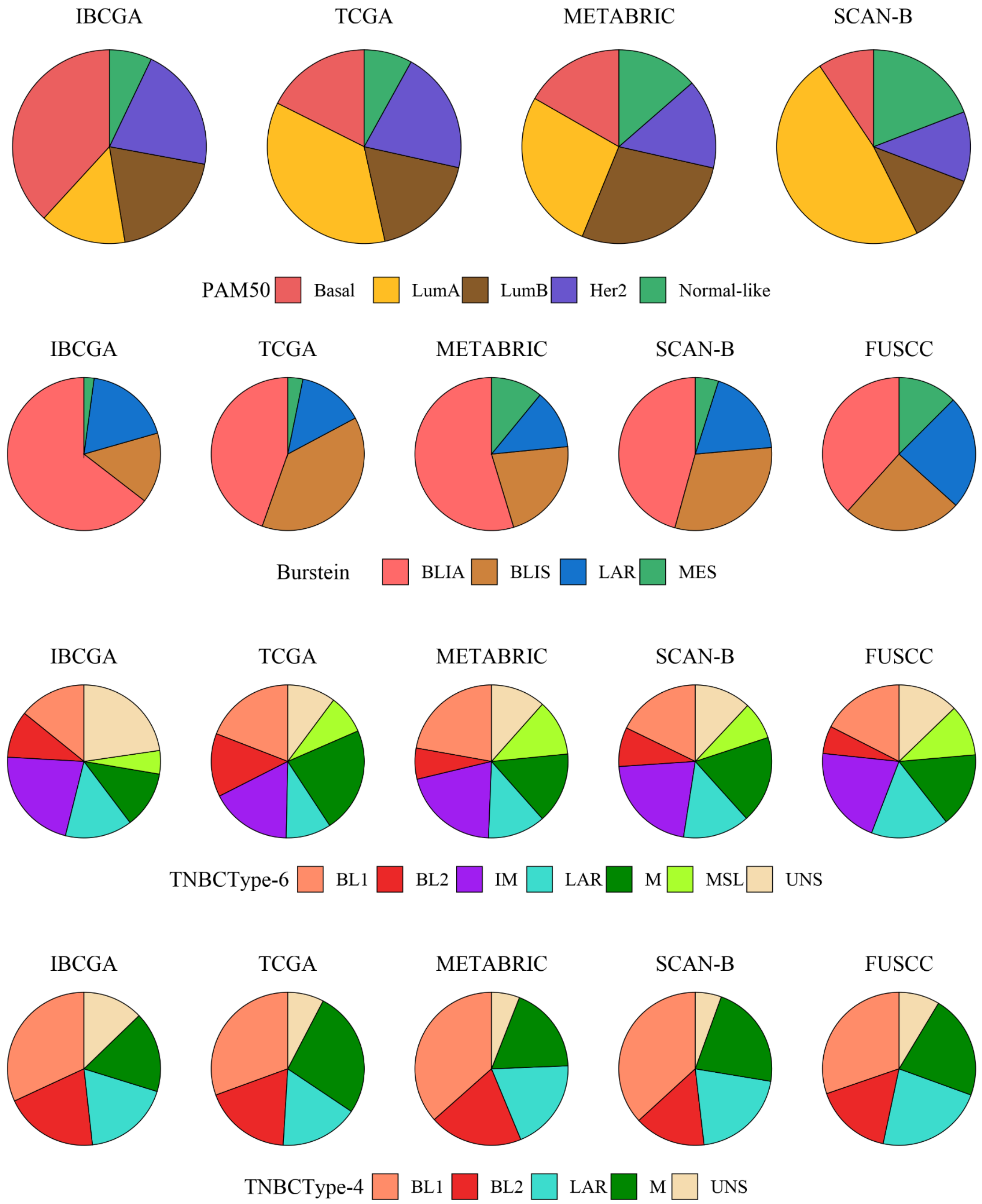


**Figure S27: Distribution of molecular subtypes across breast cancer cohorts**

(A) **PAM50 subtype distribution** across IBCGA, TCGA, METABRIC, and SCAN-B cohorts. The proportions of Basal, LumA, LumB, Her2, and Normal-like tumours are shown as pie charts for each dataset. Significant differences in subtype composition were observed among cohorts (global $P = 1.09 \times 10^{-157}$).

(B) **Burstein TNBC subtype distribution** (BLIA, BLIS, LAR, MES) across IBCGA, TCGA, METABRIC, SCAN-B, and FUSCC cohorts. The relative frequencies varied significantly between cohorts (global $P = 2.73 \times 10^{-12}$).

(C) **TNBC type-6 subtype distribution** (BL1, BL2, IM, LAR, M, MSL, UNS) across IBCGA, TCGA, METABRIC, SCAN-B, and FUSCC cohorts. Cohort-dependent differences were statistically significant (global $P = 0.00832$).

(D) **TNBC type-4 subtype distribution** (BL1, BL2, LAR, M, UNS) across IBCGA, TCGA, METABRIC, SCAN-B, and FUSCC cohorts. No significant differences were observed among cohorts (global $P = 0.135$).

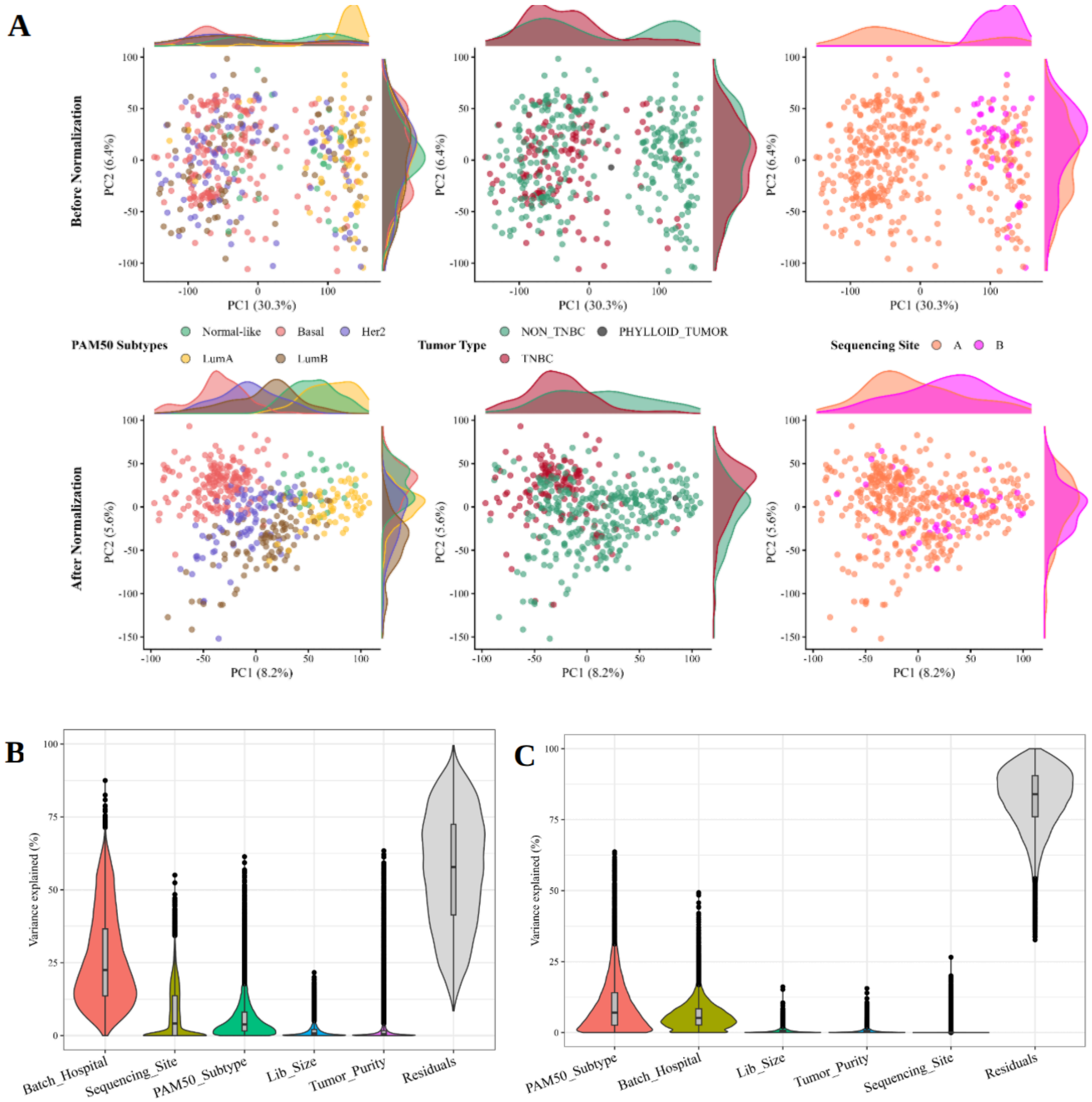

**Figure S28: Normalization reduces batch effects and enhances biological signal in PAM50 subtyping**

**(A)** Principal Component Analysis (PCA) of gene expression data before (top) and after (bottom) normalization. Prior to normalization, samples show poor separation by PAM50 subtype with substantial overlap between TNBC and non-TNBC groups, and clustering by sequencing site consistent with a strong batch effect. Following normalization, separation by biological subtypes (PAM50 classification and tumour type) becomes more distinct, while clustering by sequencing site is markedly reduced, indicating effective correction of batch-driven variation.

**(B)** Variance partition analysis before normalization reveals that batch (sequencing site) contributes more to the variance in gene expression than biological subtype, underscoring the confounding effect of technical factors.

**(C)** After normalization, PAM50 subtype becomes the major contributor to gene expression variance, indicating successful mitigation of batch effects and restoration of biologically relevant variation.

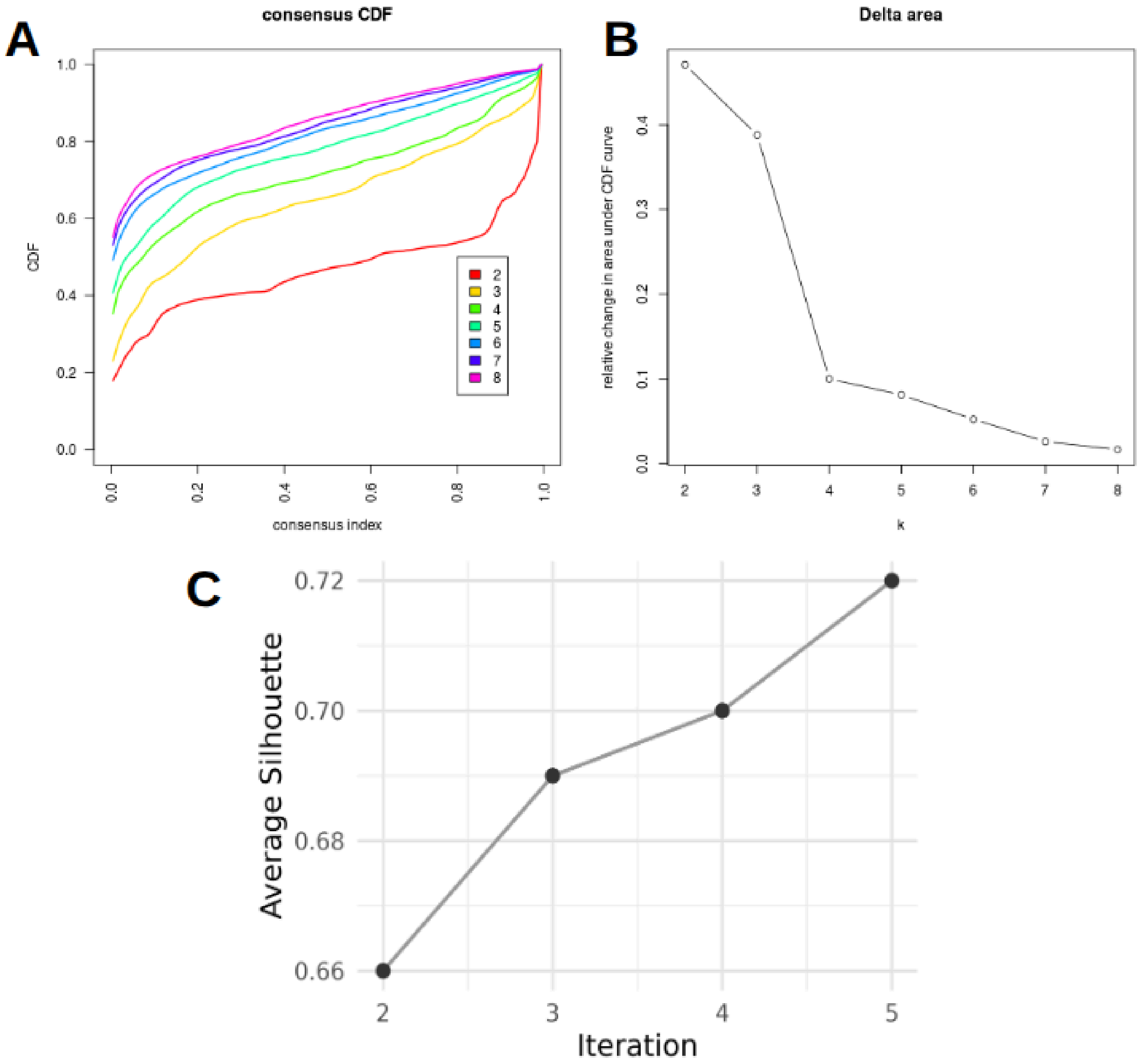


**Figure S29. All Breast tumours Consensus clustering stability and silhouette assessment.**

**(A)** Consensus cumulative distribution function (CDF) curves for cluster numbers k=2 to k=8, showing increasing clustering stability with k =4.

**(B)** Relative change in the area under the CDF curve (delta area plot), highlighting diminishing gains in stability beyond k=4.

**(C)** Average silhouette widths increased over five iterative reclustering rounds following removal of negative-silhouette samples, indicating enhanced cluster cohesion and separation.

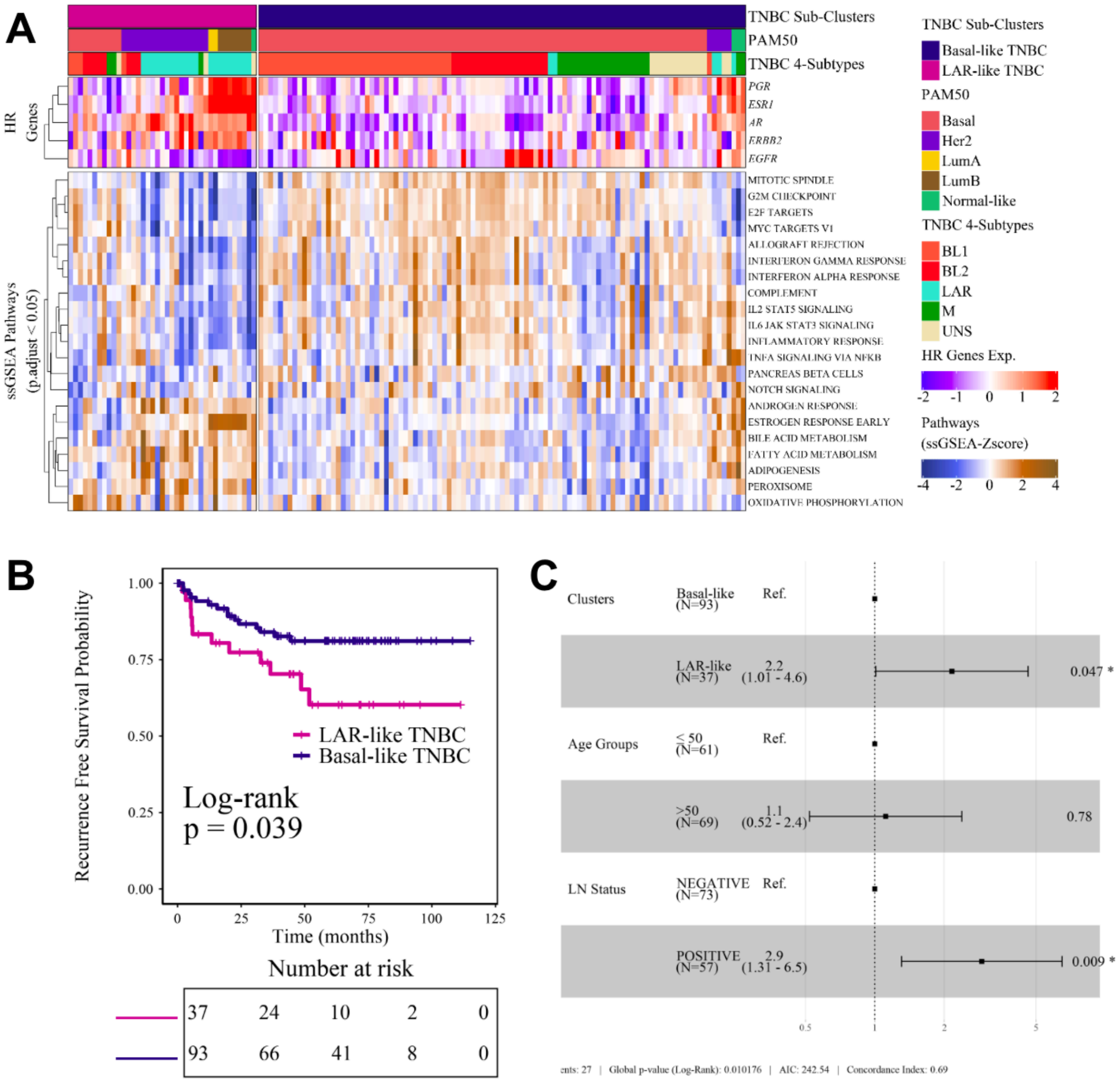


**Figure 30. Transcriptional sub-clustering and prognostic relevance of TNBC tumors**

**(A)** Heatmap showing consensus clustering of clinically defined TNBC tumours based on the top 3,500 most variable genes, identifying two major transcriptional subgroups: LAR-like TNBC and Basal-like TNBC. The top annotation indicates concordance with PAM50 and TNBCtype-4 subtype classifications. The lower panel displays significantly enriched Hallmark pathways derived from ssGSEA scores (FDR-adjusted $p < 0.05$), highlighting distinct biological programs between the two TNBC sub-clusters. **(B)** Kaplan–Meier analysis of recurrence-free survival (RFS) comparing the two TNBC sub-clusters. Patients classified as LAR-like versus Basal-like TNBC exhibited significantly different RFS outcomes (log-rank $p < 0.039$). **(C)** Multivariable Cox proportional hazards regression forest plot evaluating the independent prognostic impact of TNBC sub-clusters after adjustment for clinical covariates including age group and lymph node status. Cluster assignment remained significantly associated with recurrence risk ($p$ = 0.047), supporting the clinical relevance of the identified transcriptional subtypes.

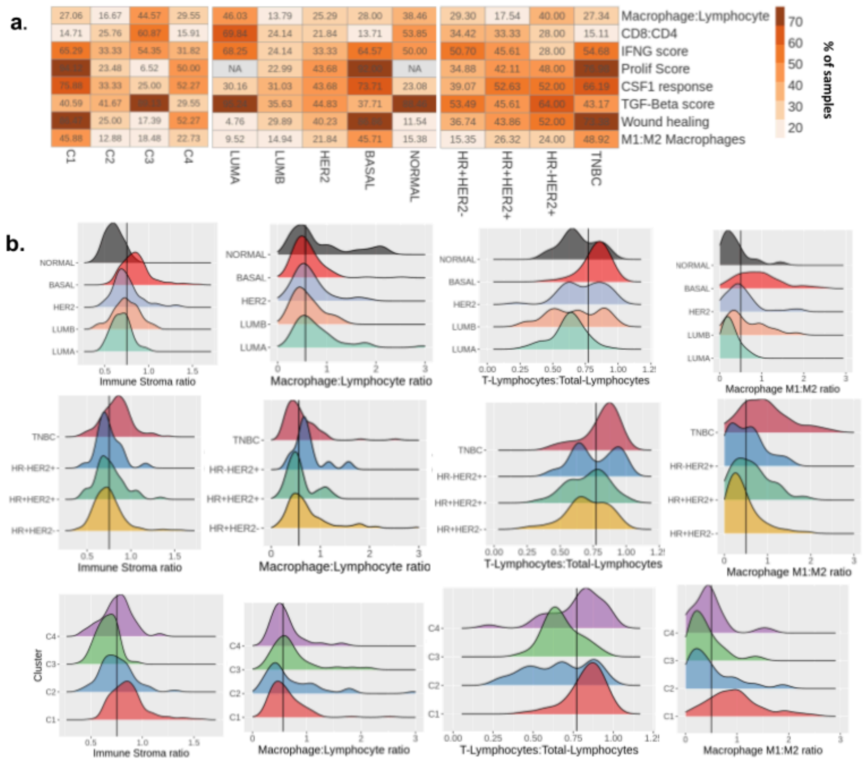


Figure S31: **Density distributions of immune and stromal cell densities across breast cancer sample groups**. (**a**) The heatmap shows percentage of samples having higher scores than median value for the immune estimates that are stratified by consensus clusters (C1-C4) and IHC-defined subtype groupings. (**b**) Densities of immune:stroma, Macrophage:Lymphocyte, T-Lymphocyte:Total-Lymphocytes and Macrophage M1:M2 are compared across four IHC-defined BC subtypes (**top**) and consensus clusters (**bottom**). The density distributions of CIBERSORT immune cell abundances with significant immune cell infiltration ($p < 0.05$, n = 245) and ESTIMATE scores for the total cohort (n = 438).

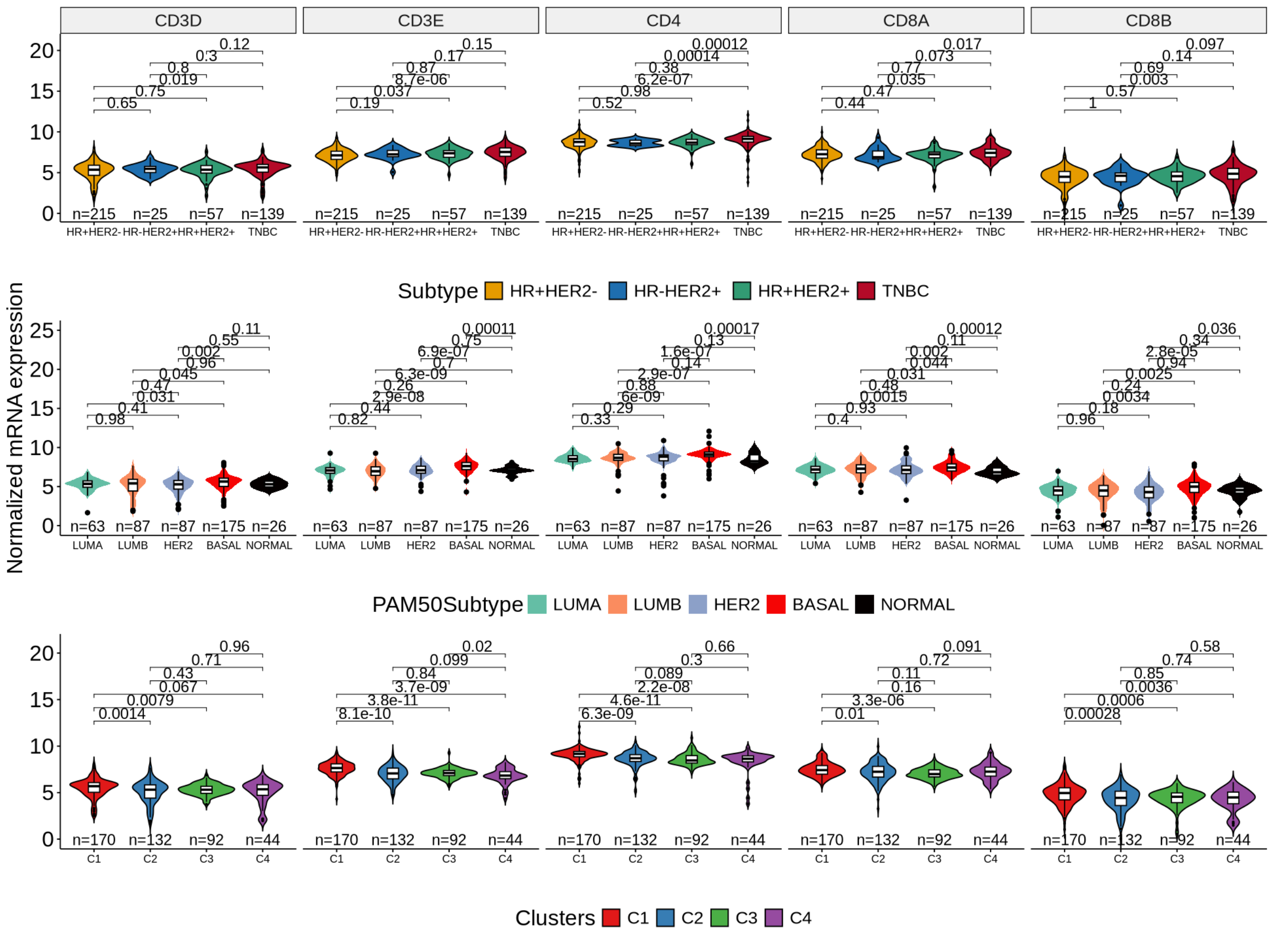


Figure S32: Distribution of mRNA expression of T-cell markers (CD3D, CD3E, CD4, CD8A, CD8B) across IHC-defined BC Subtypes (top), PAM50 subtypes (middle), and consensus clusters (bottom) is shown. Pairwise comparisons were performed using the Wilcoxon rank-sum test. The p-values were adjusted using the FDR method.

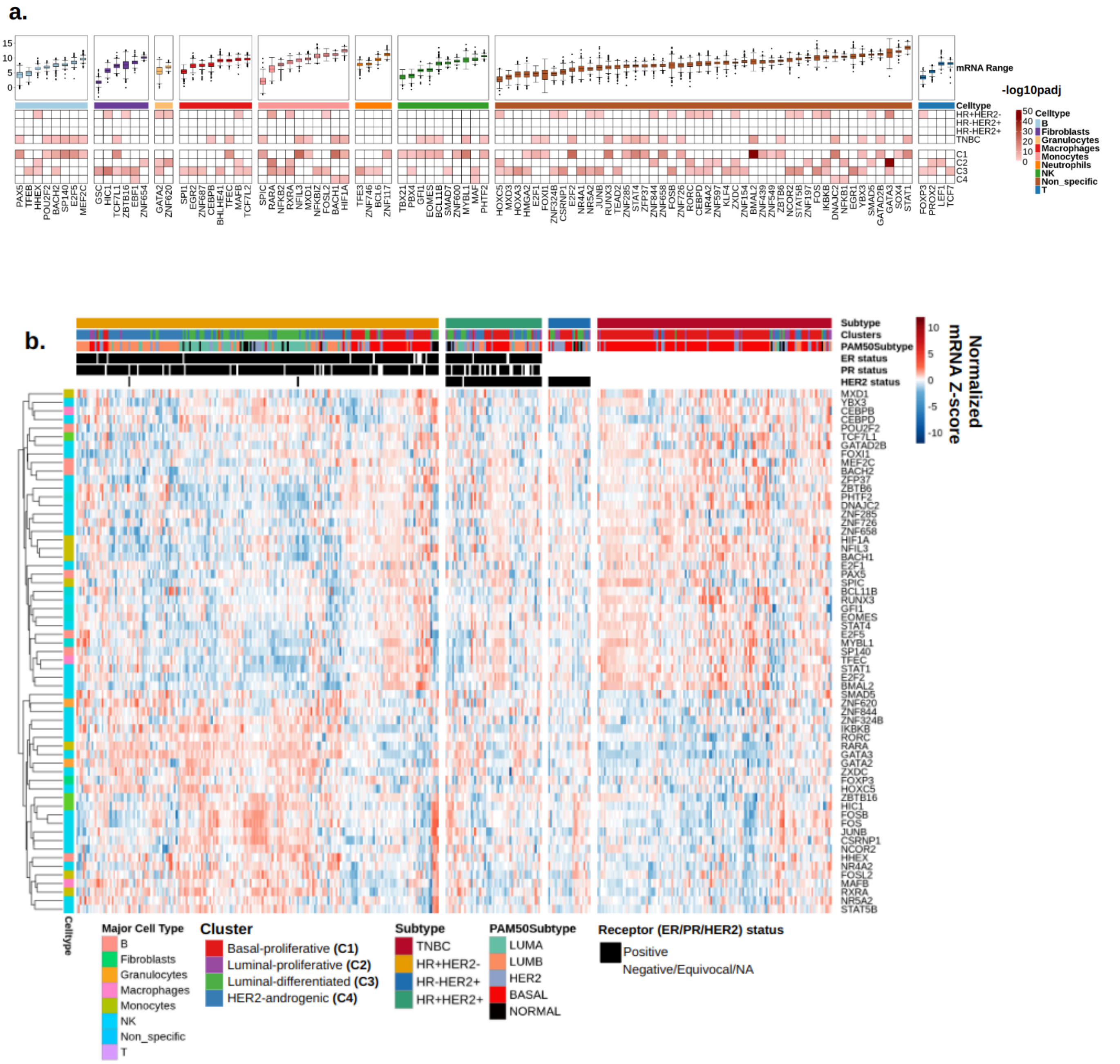


Figure S33: (**a**) Plot depicts the level of significance (-log10pval) of upregulation of immune cell transcriptional factors across each of the four subtypes (HR+/HER2-, HR+/HER2+, HR-/HER2+ and TNBC) and unsupervised sample clusters. The column annotations (top) show the major cell type that expresses the transcriptional factor as specified in the Human Protein Atlas Single Cell Cluster annotations. (**b**) The heatmap depicts the expression profile of differentially expressed transcriptional factors (padj < 0.05) across the IHC-defined BC subtypes. The annotation columns (top) of the heatmap display the subtype, Sample cluster,

PAM50 class & IHC-receptor expression.

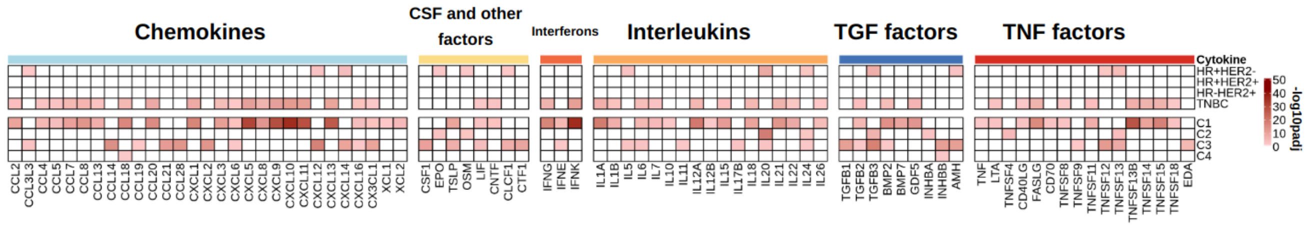


Figure S34: Plot depicts the level of significance (-log10pval) of upregulation of cytokines across each of the four subtypes (HR+/HER2-, HR+/HER2+, HR-/HER2+ and TNBC) and unsupervised sample clusters. The values of -log10 FDR adjusted pvalues for significantly upregulated cytokines across IHC-defined BC subtypes and consensus cluster are depicted ($q < 0.05$). The comparisons were made using a one-tailed Wilcoxon rank-sum test in a one-vs-rest manner for IHC-defined BC subtypes and consensus clusters independently. The annotation column (top) shows the class of cytokines as per the KEGG cytokine and neuropeptide classification (hsa04052).

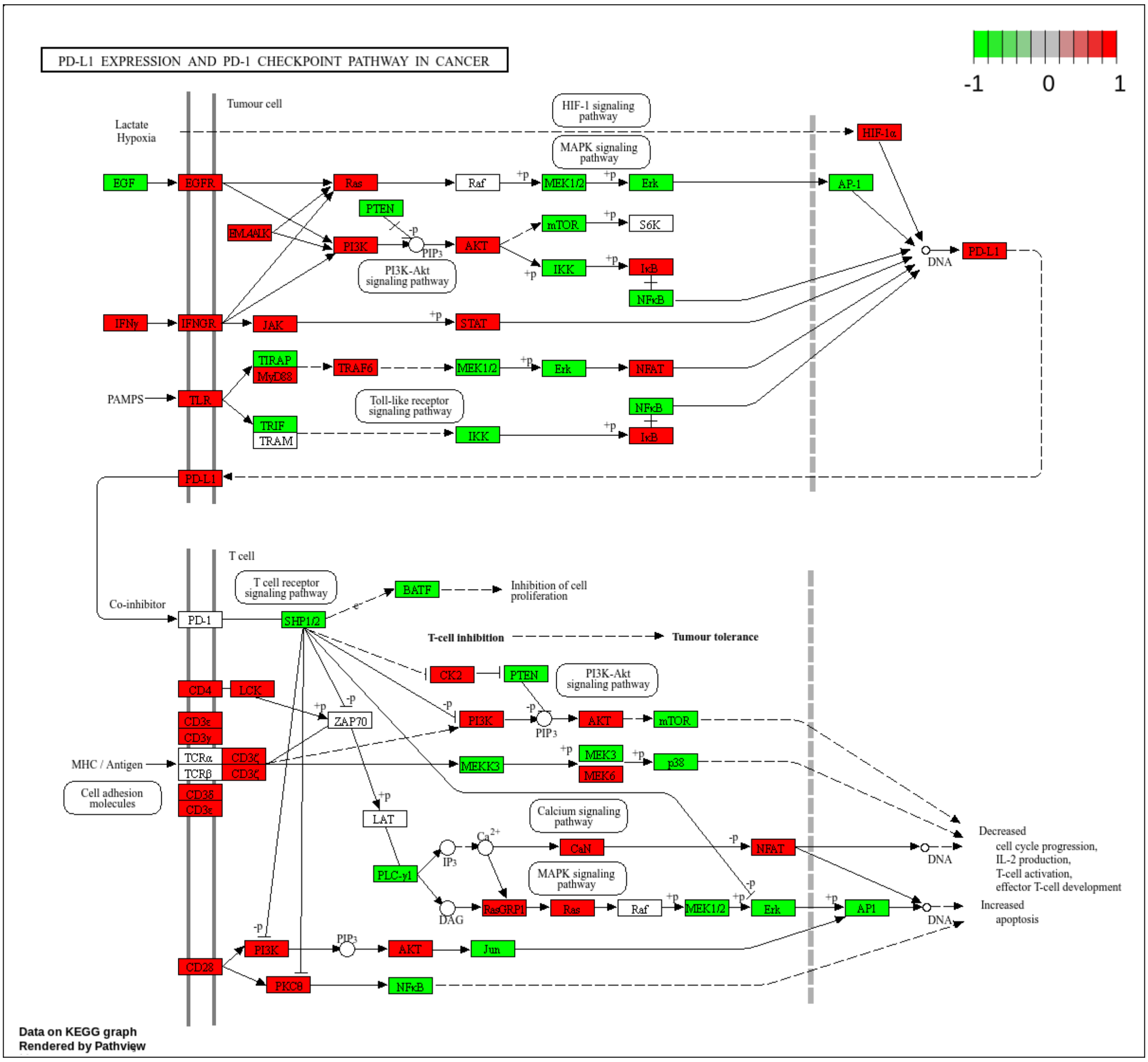

Figure S35: The genes involved in TH1 and Th2 polarization that are differentially regulated in C1 cluster samples when compared to other clusters are depicted in KEGG pathview map ($q$ < 0.05). The scale indicates the level of significance (signed -log10($q$)). The genes highlighted in red indicate overexpression whereas genes colored in green indicated lower expression in TNBC samples as compared to non-TNBC samples ($q$ < 0.05).

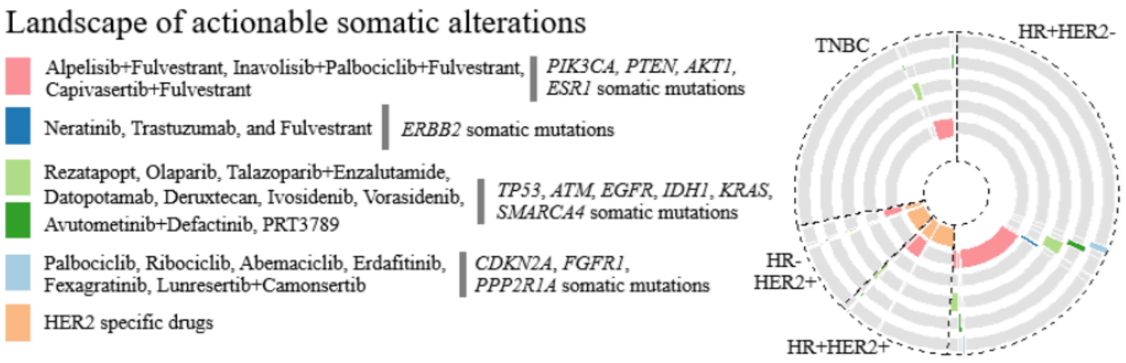


Figure S36: Among the coding somatic mutations, we found 209 that are catalogued in OncoKB (www.oncokb.org). Among these somatic mutations, FDA approved ("support level 1") drugs for breast ductal carcinoma in-situ are available against 42 mutations in 194 (38.72%) patients - comprising 35 targetable PIK3CA mutations (R88Q, I102del, I102_E103delinsK, P104_V105del, P104_V105delinsL, G106R/V, G106_R108del, E110del, K111_N114del, G118D, V344G, N345K/S, D350N, C420R, E453K/Q, E542K, E545A/K, Q546E/K/P/R, E970K, T1025A, M1043V, N1044Y, H1047L/R/Y, G1049R/S, N1068Kfs*5) [available drug combinations: Alpelisib+Fulvestrant, Inavolisib+Palbociclib+Fulvestrant, Capivasertib+Fulvestrant, 5 PTEN mutations (X70_splice, D92Y, R130*, R159S, X212_splice) Capivasertib+Fulvestrant, 1 AKT1 (E17K) (Capivasertib+Fulvestrant) and 1 ESR1 mutation (D538G – Elacestrant). We identified three ERBB2 mutations (R678Q, L755S, and G778_P780dup) in 5 patients, which are targetable through a combination of Neratinib, Trastuzumab, and Fulvestrant ("support level 2": in clinical trial but not FDA approved for breast cancer). Six somatic mutations in 10 patients - TP53 (Mutation: Y220C) [Drug: Rezatapopt], ATM (R337C) [Olaparib, Talazoparib+Enzalutamide], EGFR (A647T) [Datopotamab, Deruxtecan], IDH1 (R132C) [Ivosidenib, Vorasidenib], KRAS (G12D) [Avutometinib+Defactinib], SMARCA4 (E882K) [PRT3789] - showed drug "support level 3A/B" (Supported by clinical studies, but not FDA approved to date or in clinical guideline). We also noted "support level 4" drugs available for 3 mutations - CDKN2A (D74N) [Palbociclib, Ribociclib, Abemaciclib], FGFR1 (N577K) [Erdafitinib, Fexagratinib], and PPP2R1A (R183Q) [Lunresertib+Camonsertib] in 3 patients. In summary, 205 patients (40.92%) patients in the Indian breast cancer cohort [132 (50.77%) HR+HER2-, 33 (51.56%) HR+HER2+, 17 (37.78%) HR-HER2+, 21 (16.41%) TNBC, and 2 (50%) others] were predicted to be responsive against available drugs which are either FDA-approved or in clinical trials.
